# Introducing return of results in the Million Veteran Program: Design and pilot results of the MVP-ROAR Familial Hypercholesterolemia Study

**DOI:** 10.1101/2023.10.07.23295899

**Authors:** Jason L. Vassy, Charles A. Brunette, Thomas Yi, Alicia Harrison, Mark P. Cardellino, Themistocles L. Assimes, Kurt D. Christensen, Poornima Devineni, J. Michael Gaziano, Xin Gong, Qin Hui, Joshua W. Knowles, Sumatra Muralidhar, Pradeep Natarajan, Saiju Pyarajan, Mary Gavin Sears, Yunling Shi, Amy C. Sturm, Stacey B. Whitbourne, Yan V. Sun, Morgan E. Danowski, the Million Veteran Program

**Affiliations:** VA Boston Healthcare System, Boston, MA, USA; Harvard Medical School, Boston, MA, USA; Stanford University School of Medicine, Palo Alto, CA, USA; PRecisiOn Medicine Translational Research Center, Department of Population Medicine, Harvard Pilgrim Health Care Institute; Emory University Rollins School of Public Health, Atlanta, GA; VA Atlanta Healthcare System, Decatur, GA; Veterans Health Administration, Office of Research and Development, Washington, DC; Broad Institute of Harvard and MIT, Cambridge, MA; 23andMe, Inc., Sunnyvale, CA

## Abstract

**Background:** As a mega-biobank linked to a national healthcare system, the Million Veteran Program (MVP) can directly improve the health care and health outcomes of participants. Return of genetic research results at this scale presents challenges and complexities.

**Methods:** To determine the feasibility and outcomes of returning medically actionable genetic results to MVP participants, the program launched the MVP Return Of Actionable Results (MVP-ROAR) Study, with familial hypercholesterolemia (FH) as the exemplar actionable condition. The MVP-ROAR-FH Study consists of a completed pilot phase and an ongoing randomized clinical trial (RCT), in which MVP participants are recontacted and invited to receive clinical confirmatory gene sequencing testing and a telegenetic counseling intervention. The primary outcome of the RCT is 6-month change in low-density lipoprotein cholesterol (LDL-C) between participants receiving results at baseline and those receiving results after 6 months.

**Results:** Nine MVP participants suspected to have a pathogenic variant in *low-density lipoprotein receptor* (*LDLR*) enrolled in the single-arm pilot phase of the study; one was lost to follow-up prior to confirmatory testing. Clinical sequencing confirmed the pathogenic variant for 5 of the remaining 8 participants. Six-month ΔLDL-C among enrollees after the genetic counseling intervention was −37 mg/dL (95% CI: −12 to −61; *p*=0.03).

**Conclusions:** While underscoring the importance of analytic validity and clinical confirmation of research results, the pilot phase of the MVP-ROAR-FH Study demonstrates the feasibility of a protocol to return genetic results to MVP participants and their providers. The ongoing RCT will contribute to understanding of how such a program might improve patient health care and outcomes.

## INTRODUCTION

The Million Veteran Program (MVP) is a mega-biobank that has enrolled nearly 1 million U.S military Veterans with the objective of improving understanding of how genetic characteristics, behaviors, and environmental factors affect health.^1^ While the ultimate goal of MVP is to use these new insights to enhance the health of Veterans, 9 million of whom receive health care at a Veterans Health Administration (VHA) facility, research activities beginning in 2011 initially focused on participant recruitment, biospecimen collection and processing, and data management at unprecedented scale. MVP participants have provided broad consent to the research use of their genomic, survey, and medical record data without the expectation of learning their own genetic results or having those results impact their individual health care.

In the decade since MVP began enrollment, genomic discovery has increased our understanding of the relationship between the human genome and health and disease. The clinical validity and potential translation of this knowledge has been accelerated by consensus-building and standardization of key aspects of genomic interpretation, including how to classify variants as disease-causing and identify the genes with sufficient clinical validity and utility to be considered for reporting to patients if incidentally identified during genomic analysis.^2–5^ Translating these discoveries to improving health care and health outcomes has long been a stated goal of MVP, a research platform built on participant trust and partnership. Healthcare system-linked biobanks like MVP offer the opportunity to advance these goals by identifying genetic risk factors deemed clinically important and reporting them to the clinical setting. Although return of individual genetic results back to participants or their healthcare providers was not a part of the original MVP design or informed consent process, the research program was envisioned from the outset as an enduring research platform that could evolve alongside scientific, ethical, and clinical advances.

The opportunity to use MVP data to improve the health care of individual Veterans is increasingly recognized as potentially beneficial and even life-saving. At the same time, features of MVP present challenges to implementing such return of results at national scale, including the informed consent, non-clinical specimen collection, and the complex clinical and research regulatory environment of a large national organization. To begin to address these challenges, in 2019 MVP launched the MVP Return Of Actionable Results (MVP-ROAR) - Familial Hypercholesterolemia (FH) Study, a trial of the clinical confirmation and return of FH-associated genetic results to MVP participants and their healthcare providers. Here, we describe the rationale, protocol, and pilot phase results of the MVP-ROAR-FH Study, illustrating early challenges and successes of returning results to MVP participants.

## METHODS

### Setting

#### Veterans Health Administration (VHA)

The VHA of the U.S. Department of Veterans Affairs is the largest integrated health care system in the United States, caring for over 9 million military Veterans annually across more than 1,200 health care facilities in all 50 states, the District of Columbia, the US territories, and the Philippines.^6^ Veterans may receive VHA healthcare if they meet certain requirements related to military service, disability, and income. While many VHA patients receive the majority of their care at VHA facilities, about half receive at least some healthcare services outside VHA.^7^

#### Million Veteran Program

Launched in 2011, MVP is a national research program designed to explore how genes, lifestyle, and military exposures affect health among VHA healthcare users.^1^ Upon enrollment, participants complete a baseline survey, provide a blood specimen, and provide consent for researchers to access their electronic health record (EHR) data for research. As of October 2023, over 980,000 Veterans have enrolled across more than 60 MVP primary enrollment locations and through the MVP Online platform.^8^ MVP participants have a mean age of 62 years at enrollment and are primarily male (90%); non-Hispanic white, non-Hispanic Black, and Hispanic race and ethnicity are reported by 75%, 18%, and 8%, respectively. At baseline enrollment, MVP blood samples are collected through research protocols, not under Clinical Laboratory Improvement Amendments (CLIA) regulatory standards. As of April 2018, specimens from the initial tranche of 455,789 MVP participants had undergone genotyping and quality control on the MVP 1.0 custom Axiom array, described in detail previously.^9^ In brief, the array consists of 668,280 genetic markers passing quality control, including standard biobank content and novel content enriched for diverse ancestry populations and for diseases of relevance to the VHA patient population. Genotype data from subsequent tranches of participants, in addition to whole-genome sequence data, will be released over the next two years.

### Familial hypercholesterolemia as an exemplar condition for return of results

Recognizing the potential challenges and benefits to returning genetic results to MVP participants, in December 2018 MVP leadership and the VHA Office of Research and Development convened a planning meeting to develop a pilot project of the return of genetic results. Although dozens of genes are considered to be potentially actionable by various stakeholders,^10,11^ familial hypercholesterolemia (FH) was proposed as an ideal test case to pilot the return of genetic results in the MVP population. With a prevalence between 1:250 and 1:300 in the US,^12,13^ FH is one of the most common monogenic diseases, and yet an estimated 90% of FH cases in the US remain undiagnosed.^14^ It is characterized by markedly increased low-density lipoprotein cholesterol (LDL-C) levels and risk of premature coronary heart disease,^15–17^ such that cholesterol-lowering treatment recommendations are more aggressive for individuals with FH compared with those with common, multifactorial hypercholesterolemia.^18,19^ Moreover, cascade screening among relatives of patients with FH is endorsed by professional organizations and the Centers for Disease Control and Prevention as an efficient, cost-effective method to detect undiagnosed cases.^14,18,20^ Thus, notifying MVP participants that they might carry an undiagnosed FH-associated genetic variant has the potential to improve the lives of both the Veteran and their family. Finally, because primary care providers (PCPs) are the principal clinicians who screen for and manage hypercholesterolemia, FH was considered a monogenic disease with familiar clinical anchoring for PCPs and with straightforward treatment guidelines they could implement.^19^ With this background, MVP launched the MVP-ROAR-FH Study in 2019, with the following objectives: 1) to develop a process to recontact MVP participants for clinical confirmation of FH variants; 2) to promote the effective management of FH by returning results to MVP participants and supporting them and their healthcare providers with informational resources, and 3) to measure the impact of returning actionable genetic variants on medical management, health outcomes, and Veteran quality of life.

### Rationale for a randomized clinical trial design

The MVP-ROAR-FH Study consists of a single-arm pilot trial and a subsequent randomized clinical trial (RCT) of immediate versus delayed confirmation and return of FH-associated genetic results (ClinicalTrials.gov Identifier: NCT04178122). Despite compelling reasons to identify and return actionable genetic results to participants, the state of the science and current practice create equipoise around the question of whether to do so in MVP. First, the American College of Medical Genetics and Genomics (ACMG) recommendation that laboratories identify disease-causing variants in actionable genes applies to patients undergoing exome or genome sequencing for their clinical care, not for participation in research.^10^ Second, MVP samples are not collected under CLIA conditions and thus do not meet regulatory requirements for clinical decision-making. Third, genotyping arrays have variable ability to call rare variants such as those causative for FH, raising the possibility of false positive and negative results in MVP data. Fourth, MVP participants did not explicitly consent to receiving results from their genetic data during study enrollment. In this context, an RCT design was deemed an ethically justifiable method for generating rigorous evidence on the outcomes of genetic return-of-results.

### Selection of genetic variants considered for return

In the MVP-ROAR-FH context, any genetic variant considered for return to participants is assumed to be an opportunistic, not diagnostic, finding. Therefore, the study seeks to minimize false positive results by considering only variants with high analytic and clinical validity for return.

#### Clinical validity

The study considers variants in *low-density lipoprotein receptor* (*LDLR*), *apolipoprotein B* (*APOB*), *low-density lipoprotein receptor adapter protein 1* (*LDLRAP1*), and *proprotein convertase subtilisin/kexin type 9* (*PCSK9*). Only variants classified as likely pathogenic (LP) or pathogenic (P) for FH by ACMG and Association for Molecular Pathology (AMP) standards are eligible for return.^21^ These standards ask laboratories to apply a set of 28 criteria to classify each variant, using population, computational, functional, and segregation data. However, laboratories applying the same ACMG/AMP criteria can arrive at different classifications for a given variant,^5^ and content expertise is required to adjudicate the application of ACMG/AMP criteria to specific disease-gene associations. Therefore, MVP-ROAR-FH Study staff consult with the ClinGen FH Variant Curation Expert Panel to implement a curated list of variants based on FH-specific evidence and FH expert consensus.^3,22,23^ Only variants classified by such expert panels are designated with a 3- or 4-star interpretation in ClinVar.^5^ Study staff additionally consult with the clinical laboratory used for variant confirmation (Invitae Corporation, San Francisco, CA) before considering a variant for potential return.

#### Analytic validity

Using these lists of clinically valid P/LP variants, MVP-ROAR-FH staff assess whether the variants are present and reliably assayed on the MVP genotype array. At present, only variants directly genotyped on the MVP 1.0 array and passing rigorous quality control procedures are considered for return.^9^ As described in the **Results** below, the non-confirmation of several suspected FH-associated variants during the pilot trial prompted examination of the genotype calls, both at the aggregate level for each variant and at the individual level for each participant. To improve the accuracy of rare variant genotypes (minor allele frequency below 0.01%), the Rare Heterozygous Adjustment (RHA) algorithm^24^ was enabled in Release 4 genotype calling for all MVP samples [Release 3 (*n*=455,789) and Release 4 [additional *n*=206,724)]. A quality control (QC) process was developed to validate rare heterozygous genotypes called on the MVP array and to reduce the rate of false positive calls. QC parameters were generated by the RHA algorithm,^24^ which was triggered when fewer than four rare heterozygous genotypes were called per batch of ∼5000 participants. A support vector machine classifier was built to minimize false positive calls,^25^ using genotypes from next-generation sequencing (NGS) as truth. Furthermore, the heterozygous genotypes that passed the QC procedure underwent cluster plot review. NGS research data were available for 7 participants and were used to validate the QC pipeline developed for the MVP-ROAR-FH study. Results were additionally compared to those from MVP-ROAR-FH participants who had undergone commercial laboratory confirmation by gene sequencing.

### Modeling return of genetic research results in VHA

Different biobank studies have used different models of return of genetic results to participants.^26–28^ The clinical and research regulatory environment of MVP and VHA and the national landscape of primary care and genetic consultative services in the VHA shaped the model selected for the MVP-ROAR-FH Study.

#### VHA model of primary care delivery

VHA has adopted a Patient-Aligned Care Team (PACT) model of primary care, a patient-centered team-based approach with the aim of improving primary care quality, efficiency, and accessibility.^29–31^ Although the PACT model is a national VHA initiative, its policies and practices are shaped and implemented at national, regional, and local levels across the more than 1,000 VHA primary care practices nationally,^29^ resulting both in flexibility but also variability in implementation. Although the PACT model is associated with improved provider experience,^32^ VHA primary care remains an time-constrained environment susceptible to provider burnout like other primary care settings nationally.^33^

#### Genetic consultative services at VHA

Referral patterns for VHA patients requiring genetics consultation vary by location.^34^ Medical geneticists or genetic counselors are on staff at a limited number of major VHA medical centers, including those in Boston, MA, Houston, TX, and San Francisco, CA, West Haven, CT and West Los Angeles, CA. Over 80 VHA locations without an onsite genetic consultant have telehealth service agreements with the VA Genomic Medicine Service (GMS) based at the VA Salt Lake City Health Care System.^35^ The service employs 1 medical geneticist and 10 genetic counselors and received about 9,000 consults in fiscal years 2022 and 2023. The local referring provider sends an interfacility consult to GMS, where staff review the consult, make recommendations for genetic testing, and provide pre- and post-test genetic counseling, entirely remotely and often using video telehealth. Veterans receiving care at a VHA location not associated with GMS may pursue genetic consultative services at a non-VHA facility, often at local academic medical centers.

#### Research regulatory environment

As a federated organization of healthcare facilities, VHA is a system in which oversight of research activities is delegated across national, facility, and local levels. The parent MVP protocol is approved by the VA Central Institutional Review Board (IRB), and each MVP enrollment location is additionally overseen by local research regulatory processes. Given that MVP participants carrying an FH-associated variant would be located at dozens of facilities across the country, the MVP-ROAR-FH Study set out to develop a national, centralized protocol for return of results that minimized research administrative burden at the local level.

#### MVP-ROAR-FH model of return of results

Although the ultimate goal of MVP return-of-results efforts is to transition the results reporting and management directly to clinical care, MVP-ROAR-FH was designed as a research protocol that seeks to model how return-of-results might be integrated into the existing VHA clinical workflows described above. Key decisions included whether and how to involve PCPs in the process. Wanting to respect the autonomy of participants and minimize burden on PCPs, the study chose not to assign PCPs the role of informing participants about the research results and consenting them for the additional research protocol and clinical confirmation testing. At the same time, given the centrality of primary care in the PACT model of VHA care, the study reported the clinically confirmed genetic results and outcomes of the associated genetic counseling to the participant’s VA PCP and non-VA PCP, as applicable. Likewise, the study did not use existing, but limited, VHA genetic counseling resources for pre-test and post-test counseling, although the study genetic counselor used similar telehealth workflows to deliver the intervention, as described below.

### Study procedures

#### Recruitment and enrollment

The MVP-ROAR-FH Study has a separate research protocol from the parent MVP study (hereafter termed “MVP Core”). As a result, recruitment occurs through a two-step process (**Figure 1**). **<u>Supplementary File 1</u>** details the full study protocol. Briefly, MVP Core staff query the dataset for living MVP participants with an eligible variant and send eligible participants a letter introducing a new MVP-related research study “on heart disease risk” in which they will have the opportunity to “receive [their] own heart disease risk results” (**<u>Supplementary File 2</u>**). MVP Core staff provide the MVP-ROAR-FH staff with the identities of any participant not opting out of recontact after this introduction. MVP-ROAR-FH staff perform clinical chart review to confirm eligibility. Participants are ineligible if they already have a known molecular diagnosis of FH; there are no inclusion or exclusion criteria related to prior LDL-C values. MVP-ROAR-FH staff mail each participant more detailed information about the study (**Supplementary Files <u>3</u>, <u>4</u>, <u>5</u>**), after which the study genetic counselor calls the participant to review the information and obtain verbal informed consent.

**Figure 1:**
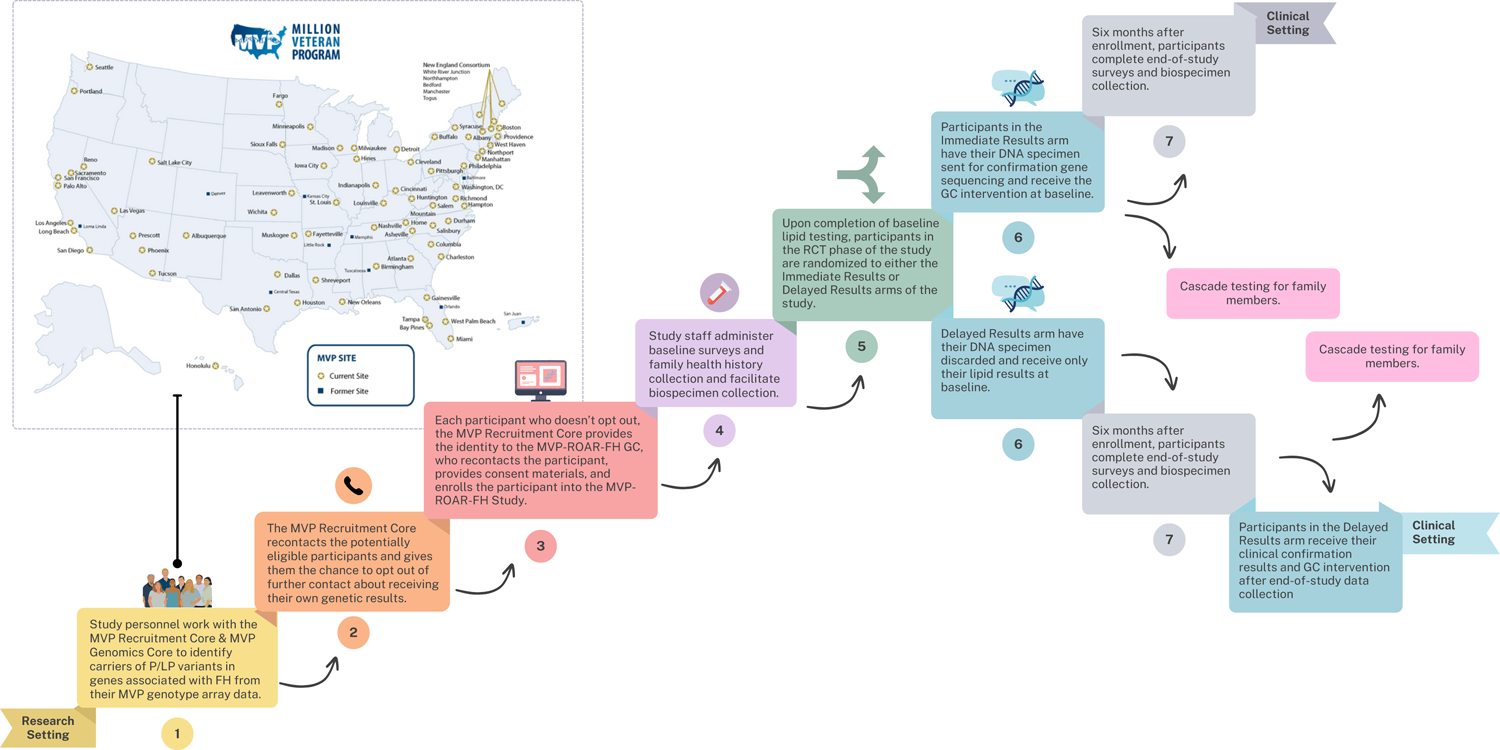
Flow diagram of study processes in the MVP-ROAR-FH Study. The MVP-ROAR-FH Study is a separate research protocol from the core MVP program. 1) Study personnel work with the MVP Recruitment Core and MVP Genomics Core to identify carriers of pathogenic/likely pathogenic variants in genes associated with FH from their MVP genotype array data. 2) The MVP Recruitment Core recontacts the potentially eligible participants and gives them the chance to opt out of further contact about receiving their own genetic results. 3) For each participant not opting out of recontact, the MVP Recruitment Core provides the identity to the MVP-ROAR-FH genetic counselor, who confirms eligibility with chart review, recontacts the participant, provides informed consent materials, and enrolls the participant into the MVP-ROAR-FH Study. 4) Study staff administer baseline surveys and family health history collection and facilitate biospecimen collection. 5) Upon completion of baseline lipid testing, participants in the RCT phase of the study are randomized to either the Immediate Results or Delayed Results arms of the study. 6) Participants in the Immediate Results arm have their DNA specimen sent for confirmation gene sequencing and receive the GC intervention at baseline, while participants in the Delayed Results arm have their DNA specimen discarded and receive only their lipid results at baseline. 7) Six months after enrollment, participants complete end-of-study surveys and biospecimen collection, including a DNA specimen for participants in the Delayed Results arm. Participants in the Delayed Results arm receive their clinical confirmation results and GC intervention after end-of-study data collection. All participants in the pilot phase of the MVP-ROAR-FH Study receive the GC intervention at baseline. Abbreviations: FH, familial hypercholesterolemia; GC, genetic counselor; MVP-ROAR-FH, Million Veteran Program Return of Actionable Results Familial Hypercholesterolemia; PCP, primary care provider; RCT, randomized clinical trial.

#### Biospecimen collection and laboratory analysis

For each enrolled participant, study staff facilitate local CLIA biospecimen collection for gene panel sequencing (either from blood or saliva) and baseline LDL-C testing. All LDL-C biospecimens are shipped to the VA Boston Healthcare System for centralized analysis on the same Abbott Architect ci8200. During the COVID-19 pandemic, procedures were amended to allow for saliva collection for DNA and use of extant clinical LDL-C values within the 6 months prior to enrollment as the study baseline LDL-C value. Each DNA specimen is shipped to a contracted CLIA-certified, College of American Pathologists (CAP)-accredited commercial laboratory (Invitae Corporation) for next-generation gene panel sequencing and deletion/duplication analysis of *LDLR, APOB, LDLRAP1*, and *PCSK9*. Study staff communicate each participant’s suspected variant to the commercial laboratory.

#### Intervention

In the pilot study, all participants were assigned to receive the intervention. In the RCT, once study staff confirm biospecimen collection, participants are randomly assigned to receive the intervention at baseline (*Immediate Results* arm) or after 6 months (*Delayed Results* arm). The return-of-results intervention consists of several components (**Table 1**). The study genetic counselor schedules a telehealth appointment by phone or videoconferencing with each participant to disclose the results of their variant confirmation and study-related LDL-C values. This genetic counseling appointment includes discussion about FH, including management guidelines and information about local and national FH resources. The genetic counselor performs a family history assessment, makes recommendations for family members, and facilitates genetic cascade testing. The genetic counselor subsequently provides by mail or email a letter reiterating information from the discussion (**<u>Supplementary Files 6</u> and <u>7</u>**), a copy of the participant’s clinical variant report (**<u>Supplementary File 8</u>**), and a family letter the participant may share with family members (**<u>Supplementary File 9</u>**). The genetic counselor also sends the participant’s PCP by mail or email the clinical LDL-C and variant reports along with provider-level information about FH treatment guidelines (**<u>Supplementary File 10</u>**). The study staff also work through local processes to facilitate the entry of the clinical variant report, study LDL-C results, and a clinical note into the EHR summarizing the result and documenting the conversation with the participant (**Supplementary Figure 1**). Participants in the Delayed Results arm and their PCP receive only their study LDL-C results by mail or email at baseline and then receive the return-of-results intervention 6 months after randomization, after completion of end-of-study data collection.

**Table 1.** Components of the MVP-ROAR-FH Study intervention. Intervention adapted from Sturm 2018.^20^ Abbreviations: EHR, electronic health record; MVP-ROAR-FH, Million Veteran Program Return of Actionable Results Familial Hypercholesterolemia; PCP, primary care provider

|  |
| --- |
| **Genetic counseling via telehealth** |
| Disclosure of genetic testing result |
| Discussion of mode of inheritance and recurrence risk to family members |
| Facilitation of family cascade testing |
| Discussion of screening, prevention, and medical management options |
| Discussion of reproductive options |
| Provision of written documentation of medical, genetic, and counseling information to patients, including family letters |
| Provision of psychosocial counseling and anticipatory guidance |
| Provision of education and resources from FH Foundation and other national organizations and advocacy groups |
| **Communication with PCP** |
| Genetic testing results and lipid results to PCP via email |
| Genetic counseling physician letter to PCP via email, including a brief explanation of FH and inheritance pattern, recommendations for specialty referral, and additional resources from the FH Foundation |
| Treatment algorithm based on the 2018 Multisociety Guideline on the Management of Blood Cholesterol via email to PCP<sup>19</sup> |
| Availability of genetic counselor as an ongoing resource to PCP as needed |
| **EHR documentation** |
| Genetic counseling note and genetic testing result entered in EHR |
| EHR alert to PCP indicating availability of results and note in EHR |

### Data collection, outcomes, and analysis

In addition to the laboratory analyses described above, study data collection includes brief surveys administered at baseline (**<u>Supplemental File 11</u>**) and 6 months after enrollment (**<u>Supplemental File 12</u>**), structured data abstraction from the VA Corporate Data Warehouse,^36^ and manual chart review of each participant’s medical record. **Table 2** shows the study outcomes and data sources. The primary outcome is the 6-month change in LDL-C, a biomarker surrogate outcome for cardiovascular disease risk reduction.^37^ Secondary outcomes are the proportion of participants meeting clinically significant LDL-C targets (<100 mg/dL for primary prevention and <70 mg/dL for secondary prevention) and the proportion of participants with intensification of lipid-lowering pharmacotherapy, a composite outcome including prescription of new monotherapy, dose escalation of existing pharmacotherapy, and addition of one or more medications to existing pharmacotherapy. Exploratory outcomes include medication adherence; cascade testing of first degree relatives; lifestyle behaviors including smoking, physical activity, and saturated fat intake; healthcare costs; and quality of life, all assessed 6 months after randomization. Enrollment of 10 participants in the non-randomized pilot phase was planned to determine feasibility. Enrollment of 244 into the RCT was planned to enable 80% power to detect a significant between-group difference in 6-month change in LDL-C (6-month LDL-C value minus baseline LDL-C value) at alpha=0.05, assessed using intention-to-treat approach and independent t-test (see **<u>Supplementary File 13</u>** for the full statistical analysis plan).

**Table 2:**
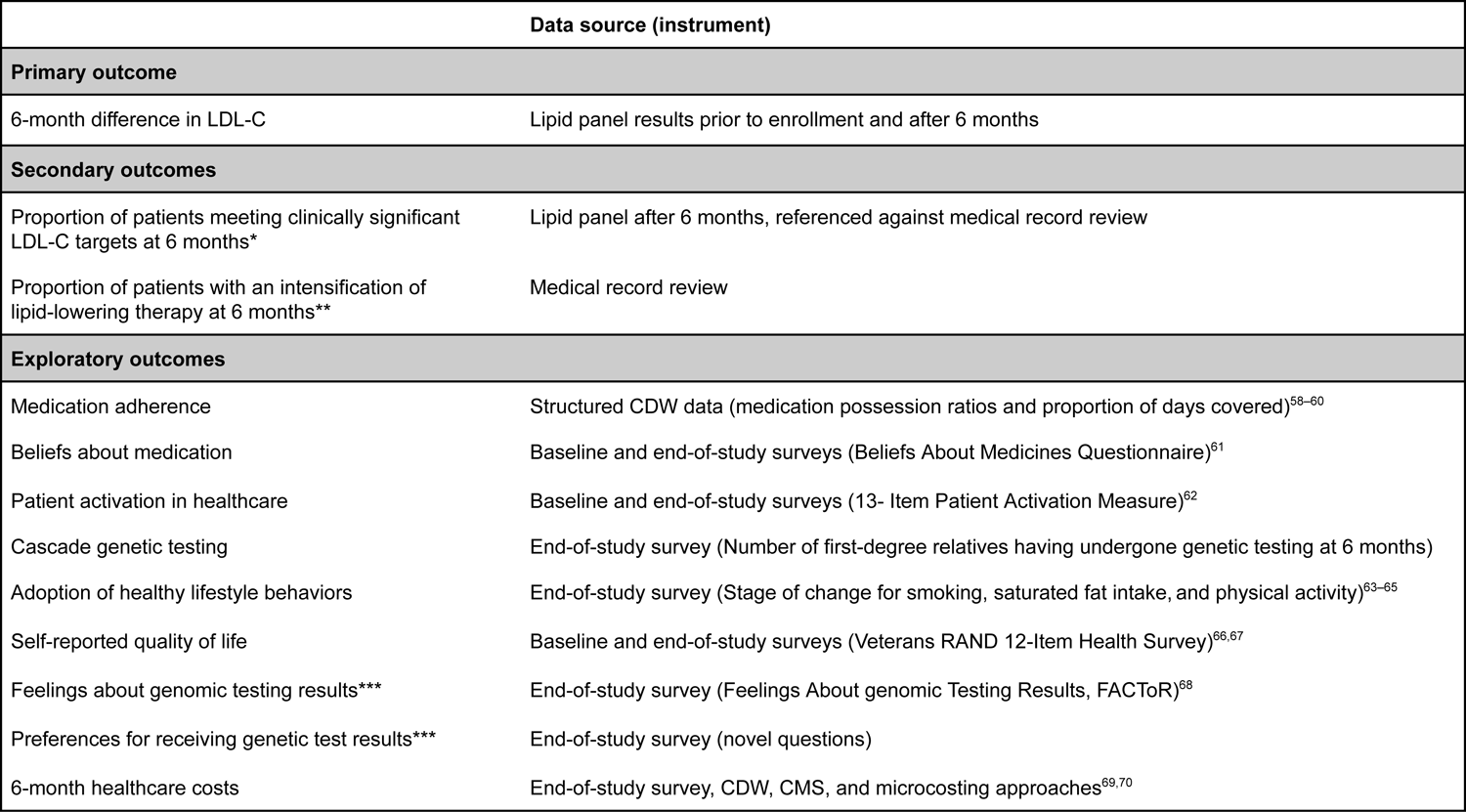
Primary, secondary, and exploratory outcomes of the MVP-ROAR-FH Study. Described in greater detail in the Statistical Analysis Plan (Supplementary File 13). *Defined as LDL-C<100 mg/dL for primary prevention and <70 mg/dL for secondary prevention, the latter being indicated for pre-existing atherosclerotic cardiovascular disease or presence of any atherosclerotic cardiovascular disease risk factors: age ≥65 years; prior percutaneous coronary intervention; prior coronary artery bypass graft; other evidence of coronary artery disease; diabetes mellitus; hypertension; chronic kidney disease; current smoking; congestive heart failure; family history of premature atherosclerotic cardiovascular disease; elevated coronary artery calcium score, lipoprotein(a), or apolipoprotein B; or ankle-brachial index <0.9. **A composite including prescription of new monotherapy, dose escalation of existing pharmacotherapy, or addition of one or more medication to existing pharmacotherapy. ***Asked only of participants in the Immediate Results arm. Abbreviations: CMS, Centers for Medicare & Medicaid Services; CDW, Corporate Data Warehouse;^36^ EHR, electronic health record; LDL-C, low-density lipoprotein cholesterol; MVP-ROAR-FH, Million Veteran Program Return Of Actionable Results Familial Hypercholesterolemia.

## RESULTS

### Variants considered for return

At study outset, we cross-referenced *LDLR* variants previously reported from MVP^38^ and classified as P/LP by at least one submitter in ClinVar, including Invitae Corporation, as of October 2019. Among these, we worked with the ClinGen FH Variant Curation Expert Panel to select an initial 3 likely pathogenic or pathogenic *LDLR* variants on the MVP genotype array for return to pilot participants: rs768563000 [(NM_000527.5(*LDLR*):c.718G>T (p.Glu240Ter), rs145787161 [NM_000527.5(*LDLR*):c.2140+1G>A], and rs377271627 [NM_000527.5(*LDLR*):c.296C>G (p.Ser99Ter)].^22^ The ClinGen Variant Curation Expert Panel has since published its consensus guidelines for *LDLR* variant classification;^23^ we have continued to work with members of this panel to identify additional variants appropriate for return.

### Pilot study recruitment, enrollment, and participant baseline characteristics

We identified 63 MVP participants with one of the 3 *LDLR* variants selected for return in the pilot phase. Of these, we selected 21 participants receiving care at VHA locations with GMS service agreements for recruitment for the pilot study. Of these, 18 did not opt out of further contact, 11 were reached by phone, and 9 enrolled. **Table 3** shows the characteristics of these enrollees, one of whom was lost to follow-up prior to baseline biospecimen collection and survey. Mean (range) age was 75 (68-89) years, 8 (89%) were men, and all reported European ancestry. Five participants (63%) reported a family history of myocardial infarction or stroke; one reported a personal history of myocardial infarction. At baseline, mean LDL-C was 113.5 (SD 70.1) mg/dL; three participants (38%) had an LDL-C>100 mg/dL. Six (67%) had an active statin prescription prior to enrollment. None reported prior genetic testing for familial hypercholesterolemia.

**Table 3:**
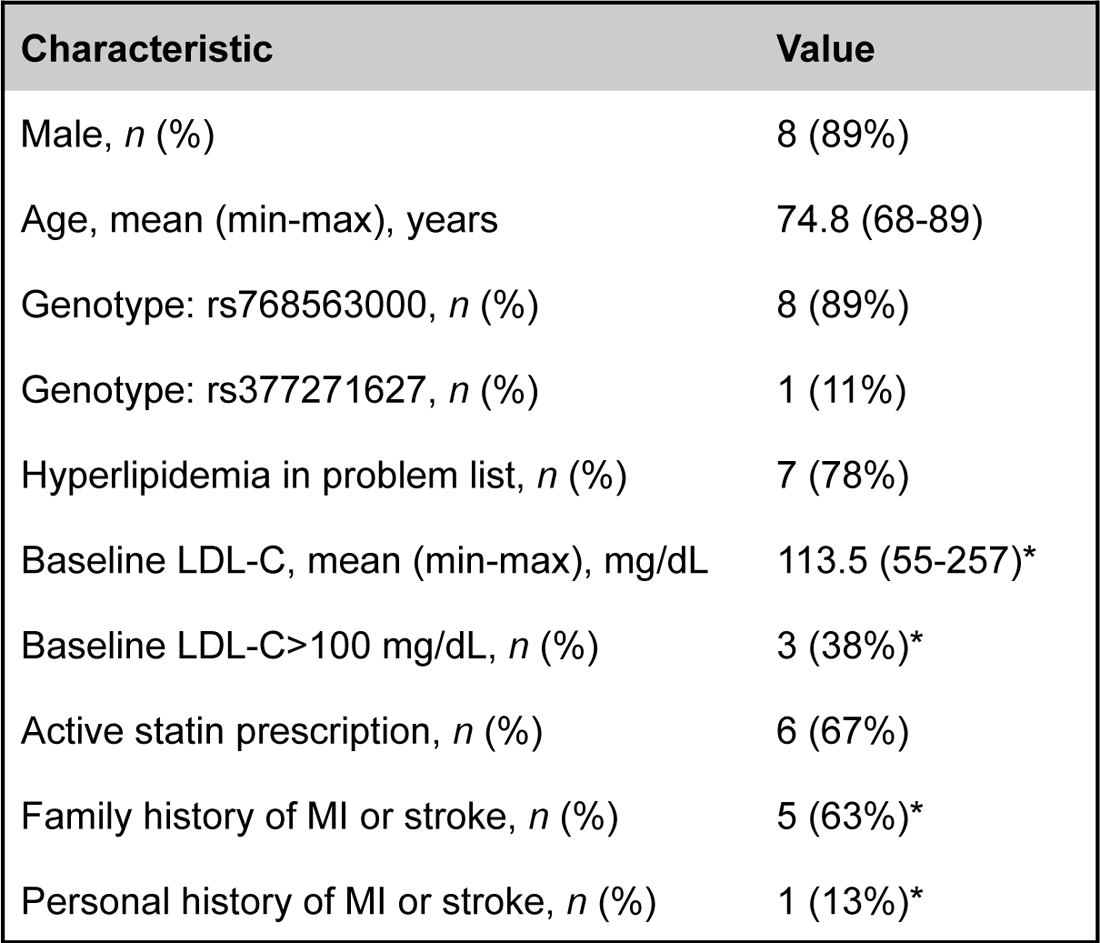
Baseline characteristics of participants in the pilot phase of the MVP-ROAR-FH Study. Data are from 9 enrollees unless otherwise specified. rs768563000 corresponds to NM_000527.5(*LDLR*):c.718G>T (p.Glu240Ter) or NM_000527.5(*LDLR*):c.718G>A (p.Glu240Lys) and rs377271627 corresponds to NM_000527.5(*LDLR*):c.296C>G (p.Ser99Ter). *Data are from 8 enrollees, after one participant was lost to follow prior to baseline data collection. Abbreviations: LDL-C, low-density lipoprotein cholesterol; MI, myocardial infarction; MVP-ROAR-FH, Million Veteran Program Return of Actionable Results Familial Hypercholesterolemia.

### Pilot study return-of-results

One participant was considered lost to follow-up prior to biospecimen collection. As described in the Methods, the MVP research result was clinically confirmed in 5/8 (63%) pilot participants (*LDLR* c.718G>A). All results were disclosed via telephone due to participant preference, and call duration ranged from 10-30 minutes. Results were also returned to 8 VA PCPs, 3 non-VA PCPs, and 2 non-VA cardiologists. Relatives of 3 pilot participants called the study genetic counselor for more information about obtaining familial variant testing.

### Pilot study outcomes

Compared to available reference populations, MVP-ROAR-FH pilot participants endorsed equivalent physical and mental health burdens, held analogous beliefs about medication overuse and harm, and scored within similar ranges on a measure of patient activation, both at baseline and after 6 months (**Table 4**).^39–42^ After 6 months, mean LDL-C was 74.6 (SD 44.0) mg/dL, 1 (13%) participant had LDL-C>100 mg/dL, and 8 (100%) had active statin prescriptions. Mean 6-month ΔLDL-C in the overall sample was −36.8 mg/dL (95% CI −12.1 to −61.4; paired *t*-test *p*=0.028). Among those with positive clinical confirmation (*n*=5), mean 6-month ΔLDL-C was −54.4 mg/dL (95% CI −12.3 to −96.5; *p*=0.023, **Figure 2**). When asked whom the research team should have contacted first about the genetic research results, 5 participants indicated themselves, 0 indicated their PCP, and 2 did not have a preference. 6 indicated that the research genetic counselor should have been the person who told them their clinical confirmation results, 0 indicated their PCP, and 1 did not have a preference. Among a total of 27 living first-degree and 56 second-degree relatives, 4 participants reported they had shared their results with a total of at least 16 and 0 family members respectively.

**Figure 2:**
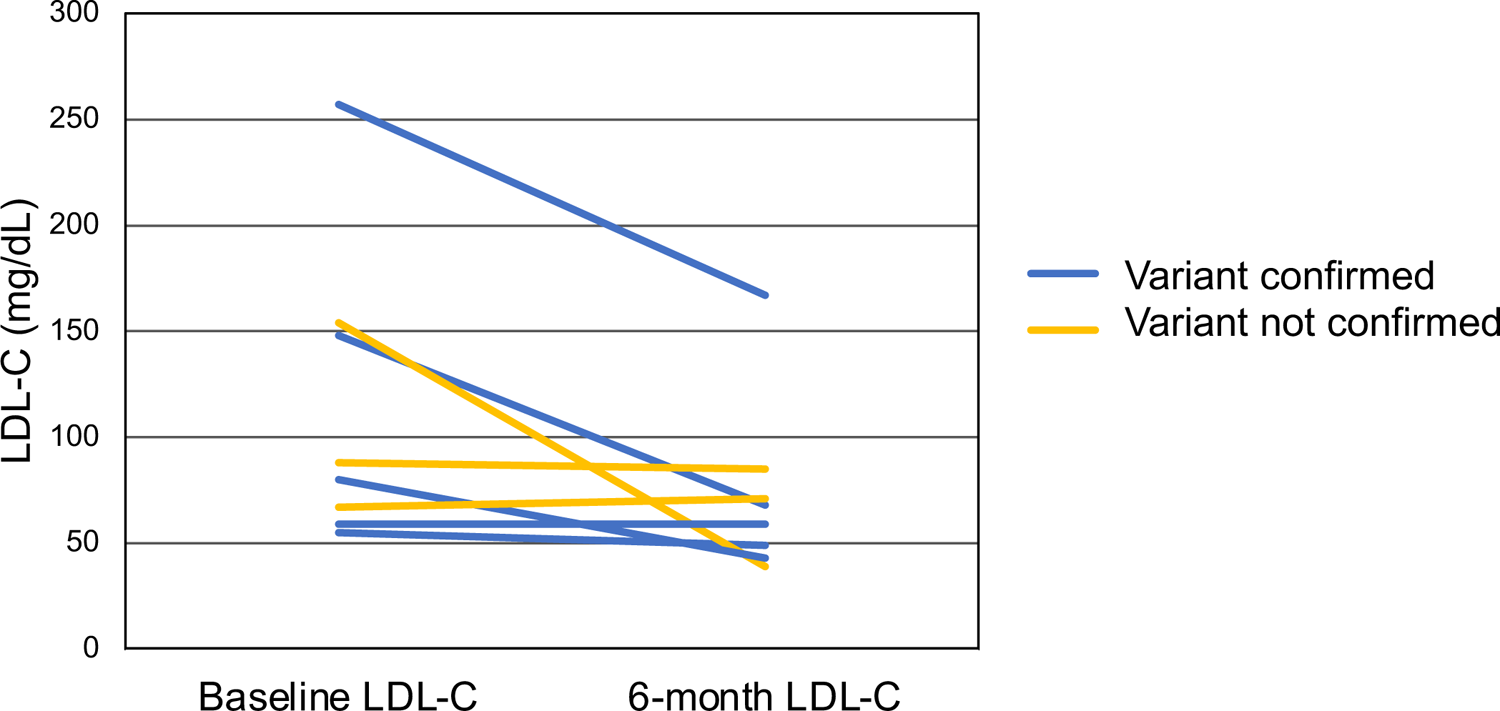
Six-month change in LDL-C among participants in the pilot phase of the MVP-ROAR-FH Study. Presentation of data is stratified by confirmation (blue lines) or non-confirmation (yellow lines) of suspected FH-associated variant from MVP genotype array data by clinical sequencing. One participant had baseline value carried forward to six months due to loss to follow-up. Abbreviations: LDL-C, low-density lipoprotein cholesterol, MVP-ROAR-FH, Million Veteran Program Return of Actionable Results Familial Hypercholesterolemia.

**Table 4:** 6-month outcomes of participants in the pilot phase of the MVP-ROAR-FH Study. Data are presented as means (standard deviation). Data are from 8 enrollees unless otherwise specified. VR-12 Physical Component is a self-reported measure of physical health status, where values <50 indicate increasingly greater disease burden. VR-12 Mental Component is a self-reported measure of psychological health status, where values <50 indicate increasingly greater disease burden. The PAM-13 (range 0-100) assesses patient knowledge, beliefs, skill, and confidence for health self-management, with scores 0-47 indicating individuals are disengaged and overwhelmed, scores 47.1-55.1 indicating individuals are becoming aware but still struggling, scores 55.2-72.4 indicating individuals are taking action and gaining control, and scores 72.5-100 indicating individuals are maintaining behaviors and pushing further. The BMQ General assesses beliefs that medicines are overused by physicians (Overuse Scale, range 4-20) and beliefs that medicines are harmful (Harm Scale, range 4-20), where higher values indicate beliefs that medications are generally overused and harmful, respectively. ^ Difference calculated by subtracting baseline survey value from 6-month survey value. * Data are from 7 enrollees, after one participant was lost to follow prior to 6-month data collection. Abbreviations: BMQ, Beliefs About Medicines Questionnaire; PAM-13, 13-item Patient Activation Measure; VR-12, Veterans RAND 12-Item Health Survey.

|  | Baseline survey | 6-month survey | Difference<sup>^</sup> |
| --- | --- | --- | --- |
| **VR-12 Physical Component** | 42.5 (11.2) | 38.9 (10.4)* | -2.2 (7.7)* |
| **VR-12 Mental Component** | 51.3 (12.7) | 49.6 (13.4)* | -1.5 (11.1)* |
| **PAM-13** | 64.4 (13.9) | 67.5 (11.6)* | 2.2 (8.1)* |
| **BMQ General** | 23.6 (6.3) | 22.0 (4.2)* | 2.1 (3.7)* |
| **BMQ General - Overuse Scale** | 13.9 (3.4) | 12.9 (2.7)* | 1.4 (2.2)* |
| **BMQ General - Harm Scale** | 9.8 (3.6) | 9.1 (2.0)* | 0.7 (2.4)* |

## DISCUSSION

As a mega-biobank linked to the largest healthcare system in the US, MVP can make important contributions to the integration of genetic research results into participants’ health care. Results from the pilot phase of the MVP-ROAR-FH Study demonstrate the feasibility and potential for improved health outcomes of returning FH-associated variants to MVP participants and their PCPs. The ongoing RCT will provide rigorous evidence on the impact of this intervention on patient outcomes.

Biobank participants often state that they would desire and expect actionable medical findings to be returned for their benefit.^43–45^ The ethics and practicalities of such return are complicated, but since the inception of MVP, emerging consensus has better defined the actionability of specific genetic results and favors the return of incidentally identified results that are analytically valid and associated with serious medical conditions.^2,10,11^ Consensus articulated by ACMG guidelines and others is that medically actionable results identified during the course of sequencing for clinical care should be offered for return.^2,10,46^ These guidelines are not intended to apply to genetic results identified incidentally in the research context, but guidelines recommend that researchers establish clear policies and procedures for such return from the outset and include this information in the informed consent process.^47–53^ Given rapid advances in multi-omic and artificial intelligence technologies, matching the content of this consent to the breadth of possible future medical discoveries is a formidable challenge. The participant partnership and trust and that MVP has fostered since its beginning are key to navigating this evolving landscape in good stewardship.

Other initiatives have identified actionable FH-associated variants among biobanks participants and implemented processes to return results to participants, including the Geisinger Health System MyCode Community Health Initiative and 7 sites in the Electronic Medical Records and Genomics network.^26–28^ In contrast to local or regional health system-linked programs, the MVP-ROAR-FH Study is national in scope. At the same time, unlike national initiatives like the All of Us Research Program,^54^ MVP’s position within the single largest US healthcare system creates the opportunity and perhaps expectation that important results will be integrated into participants’ healthcare. This national research-clinical ecosystem created complexities for the clinical confirmation and return of genetic research results. It necessitated a balance between standardizing certain procedures (*e.g.*, a standardized genetic counseling intervention administered centrally) and tailoring some procedures locally across diverse geographic areas (*e.g.*, communicating with local personnel to facilitate phlebotomy and documentation in the medical record). The study encountered other distinct challenges in returning genetic results. The absence of explicit consent at original MVP enrollment to receive individual genetic results necessitated the creation of an additional research protocol under which return-of-results activities could occur. Non-clinical biospecimen collection at enrollment and DNA analysis on the MVP genotyping array necessitated clinical confirmatory testing for any suspected pathogenic results. This decision proved wise, given that research results were not confirmed on clinical sequencing for 3 of 8 pilot phase participants, whose genotype analysis had been completed prior to implementation of an improved calling algorithm for rare variants. As of September 15, 2023, all research results returned to participants after implementation of the improved calling algorithm in the RCT phase of the MVP-ROAR-FH Study have been clinically confirmed.

The MVP-ROAR-FH Study pilot phase experience supports the following recommendations for the future of return of genetic results in MVP and other similar programs. First, consolidation of core biobank and potential return-of-results activities under a single research protocol creates regulatory efficiencies and a more streamlined participant experience. In this case, the informed consent process should describe the potential for and manner of recontact, while acknowledging to participants that scientific and medical advances might produce actionable information not anticipated at the time of consent.^45,55–57^ Second, this pilot study demonstrates the primacy of confirming the analytic validity of any incidental genetic results considered for return in the research setting. Rare variants are by nature difficult to call from genotype array data.^24^ After lower-than-expected rates of variant confirmation from clinical gene sequencing among pilot participants, improving the rare variant calling algorithm was a critical next step before proceeding with the MVP-ROAR-FH Study. Sequencing of research samples would be expected to improve rare variant calls and facilitate the expansion to other genes deemed medically actionable. Third, centralized processes improve the scalability of return-of-results. Delays often resulted from attempts at blood collection from local facilities, each of which may see only one eligible participant during a multiyear initiative. Because LDL-C change is the study’s primary outcome, blood samples are preferred in MVP-ROAR-FH, but future return-of-results from MVP might consider more scalable methods such as at-home saliva collection, a process the study adopted during the COVID-19 pandemic and continues to use for participants having difficulty presenting to a local facility. Fourth, close integration with existing VHA clinical services (*e.g.*, primary care, medical genetics, preventive cardiology) will likely improve the clinical implementation and patient outcomes from return of results. Such integration aligns with the VHA priority of patient-centered care coordination but will need to weigh the competing demands on the clinical VHA workforce. Given the rarity of many results considered for return, regional hub-and-spoke models of consultation and management might prove an efficient care delivery model.

In conclusion, the ongoing MVP-ROAR-FH Study underscores the complexities, feasibility, and potential value of returning genetic results from a large biobank linked to a national healthcare system. The study offers lessons for how future endeavors can optimize patient-participant outcomes as genomic and clinical science advance.

## Data Availability

The data produced in the present study are not currently available. Some of these data will be made available through a data repository after publication of the full clinical trial.

## Acknowledgements

This research is based on data from the Million Veteran Program, Office of Research and Development, Veterans Health Administration, and was supported by award number MVP030. Please see Supplementary File 14 for the full MVP Core Acknowledgements. The authors thank Jonathan Dryjowicz-Burek and the staff of the clinical laboratory at the VA Boston Healthcare System, West Roxbury Division, for processing and analyzing study lipid panels; Martha Paige Greene for her assistance with figure preparation; and Annika Toivonen for assistance in data collection. This publication does not represent the views of the Department of Veterans Affairs or the United States Government.

## Supplementary figures

**Supplementary Figure 1:** Example intervention note in electronic health record

## Supplementary files

**<u>Supplementary File 1</u>:** MVP-ROAR-FH Study protocol

**<u>Supplementary File 2</u>:** MVP Core introduction letter and opt-out postcard

**<u>Supplementary File 3</u>:** MVP-ROAR-FH Study recruitment letter

**<u>Supplementary File 4</u>:** MVP-ROAR-FH Study informed consent form (pilot phase)

**<u>Supplementary File 5:</u>** MVP-ROAR-FH Study informed consent form (RCT phase)

**<u>Supplementary File 6</u>:** MVP-ROAR-FH Study positive results letter

**<u>Supplementary File 7</u>:** MVP-ROAR-FH Study negative results letter

**<u>Supplementary File 8</u>:** Example clinical confirmation results report

**<u>Supplementary File 9</u>:** MVP-ROAR-FH Study family letter

**<u>Supplementary File 10:</u>** MVP-ROAR-FH Study PCP letter and resources

**<u>Supplementary File 11</u>:** MVP-ROAR-FH Study baseline survey

**<u>Supplementary File 12</u>:** MVP-ROAR-FH Study 6-month survey

**<u>Supplementary File 13</u>:** MVP-ROAR-FH Study statistical analysis plan

**<u>Supplementary File 14</u>:** MVP Core Acknowledgement for Publications

## REFERENCES

1. Gaziano JM, Concato J, Brophy M. Million Veteran Program: A mega-biobank to study genetic influences on health and disease. J Clin Epidemiol. 2016;70:214–223.

2. Green RC, Berg JS, Grody WW. ACMG recommendations for reporting of incidental findings in clinical exome and genome sequencing. Genet Med. 2013;15(7):565–574.

3. Rivera-Munoz EA, Milko LV, Harrison SM. ClinGen Variant Curation Expert Panel experiences and standardized processes for disease and gene-level specification of the ACMG/AMP guidelines for sequence variant interpretation. Hum Mutat. 2018;39(11):1614–1622.

4. Hunter JE, Irving SA, Biesecker LG, et al. A standardized, evidence-based protocol to assess clinical actionability of genetic disorders associated with genomic variation. Genet Med. 2016;18(12):1258–1268. doi:10.1038/gim.2016.40

5. Landrum MJ, Lee JM, Benson M, et al. ClinVar: improving access to variant interpretations and supporting evidence. Nucleic Acids Res. 2018;46(D1):D1062–D1067. doi:10.1093/nar/gkx1153

6. U.S. Department of Veterans Affairs. Department of Veterans Affairs Fiscal Years 2022-28 Strategic Plan.; 2022. Accessed August 30, 2023. https://department.va.gov/wp-content/uploads/2022/09/va-strategic-plan-2022-2028.pdf

7. Wang ZJ, Dhanireddy P, Prince C, Larsen M, Schrimpf M, Pearman G. 2021 Survey of Veteran Enrollees’ Health and Use of Health Care.; 2021. Accessed August 30, 2023. https://www.va.gov/VHASTRATEGY/SOE2021/2021_Enrollee_Data_Findings_Report-508_Compliant.pdf

8. Whitbourne SB, Li Y, Brewer JVV, et al. Overview of Efforts to Increase Women Enrollment in the Veterans Affairs Million Veteran Program. Health Equity. 2023;7(1):324–332. doi:10.1089/heq.2023.0006

9. Hunter-Zinck H, Shi Y, Li M, et al. Genotyping Array Design and Data Quality Control in the Million Veteran Program. Am J Hum Genet. 2020;106(4):535–548. doi:10.1016/j.ajhg.2020.03.004

10. Miller DT, Lee K, Abul-Husn NS, et al. ACMG SF v3.2 list for reporting of secondary findings in clinical exome and genome sequencing: A policy statement of the American College of Medical Genetics and Genomics (ACMG). Genet Med Off J Am Coll Med Genet. 2023;25(8):100866. doi:10.1016/j.gim.2023.100866

11. Berg JS, Foreman AK, O’Daniel JM, et al. A semiquantitative metric for evaluating clinical actionability of incidental or secondary findings from genome-scale sequencing. Genet Med. 2016;18(5):467–475. doi:10.1038/gim.2015.104

12. de Ferranti SD, Rodday AM, Mendelson MM, Wong JB, Leslie LK, Sheldrick RC. Prevalence of Familial Hypercholesterolemia in the 1999 to 2012 United States National Health and Nutrition Examination Surveys (NHANES). Circulation. 2016;133(11):1067–1072. doi:10.1161/circulationaha.115.018791

13. Beheshti SO, Madsen CM, Varbo A, Nordestgaard BG. Worldwide Prevalence of Familial Hypercholesterolemia: Meta-Analyses of 11 Million Subjects. J Am Coll Cardiol. 2020;75(20):2553–2566. doi:10.1016/j.jacc.2020.03.057

14. Knowles JW, Rader DJ, Khoury MJ. Cascade Screening for Familial Hypercholesterolemia and the Use of Genetic Testing. JAMA. 2017;318(4):381–382.

15. Youngblom E, Pariani M, Knowles JW. Familial Hypercholesterolemia. In: Adam MP, Ardinger HH, Pagon RA, eds. GeneReviews((R)). University of Washington, Seattle; 1993.

16. Benn M, Watts GF, Tybjaerg-Hansen A, Nordestgaard BG. Familial hypercholesterolemia in the Danish general population: prevalence, coronary artery disease, and cholesterol-lowering medication. J Clin Endocrinol Metab. 2012;97(11):3956–3964.

17. Khera AV, Won HH, Peloso GM. Diagnostic Yield and Clinical Utility of Sequencing Familial Hypercholesterolemia Genes in Patients With Severe Hypercholesterolemia. J Am Coll Cardiol. 2016;67(22):2578–2589.

18. Gidding SS, Champagne MA, de Ferranti SD. The Agenda for Familial Hypercholesterolemia: A Scientific Statement From the American Heart Association. Circulation. 2015;132(22):2167–2192.

19. Grundy SM, Stone NJ, Bailey AL. 2018 AHA/ACC/AACVPR/AAPA/ABC/ACPM/ADA/AGS/APhA/ASPC/NLA/PCNA Guideline on the Management of Blood Cholesterol. Circulation. 2018;0(0):CIR.0000000000000625.

20. Sturm AC, Knowles JW, Gidding SS. Clinical Genetic Testing for Familial Hypercholesterolemia: JACC Scientific Expert Panel. J Am Coll Cardiol. 2018;72(6):662–680.

21. Richards S, Aziz N, Bale S. Standards and guidelines for the interpretation of sequence variants: a joint consensus recommendation of the American College of Medical Genetics and Genomics and the Association for Molecular Pathology. Genet Med. Published online 2015.

22. Iacocca MA, Chora JR, Carrie A. ClinVar database of global familial hypercholesterolemia-associated DNA variants. Hum Mutat. 2018;39(11):1631–1640.

23. Chora JR, Iacocca MA, Tichý L, et al. The Clinical Genome Resource (ClinGen) Familial Hypercholesterolemia Variant Curation Expert Panel consensus guidelines for LDLR variant classification. Genet Med Off J Am Coll Med Genet. 2022;24(2):293–306. doi:10.1016/j.gim.2021.09.012

24. Mizrahi-Man O, Woehrmann MH, Webster TA, et al. Novel genotyping algorithms for rare variants significantly improve the accuracy of Applied Biosystems^TM^ Axiom^TM^ array genotyping calls: Retrospective evaluation of UK Biobank array data. PloS One. 2022;17(11):e0277680. doi:10.1371/journal.pone.0277680

25. Webster T, Mizrahi-Man O, Mittal A, Schmidt J. Improved genotyping of rare variants from AppliedBiosystems^TM^ Axiom^TM^ microarrays, using Support Vector Machine (SVM) prediction models. In:; 2022.

26. Dikilitas O, Sherafati A, Saadatagah S, et al. Familial Hypercholesterolemia in the Electronic Medical Records and Genomics Network: Prevalence, Penetrance, Cardiovascular Risk, and Outcomes After Return of Results. Circ Genomic Precis Med. 2023;16(2):e003816. doi:10.1161/CIRCGEN.122.003816

27. Jones LK, Kulchak Rahm A, Manickam K, et al. Healthcare Utilization and Patients’ Perspectives After Receiving a Positive Genetic Test for Familial Hypercholesterolemia. Circ Genomic Precis Med. 2018;11(8):e002146. doi:10.1161/CIRCGEN.118.002146

28. Jones LK, Chen N, Hassen DA, et al. Impact of a Population Genomic Screening Program on Health Behaviors Related to Familial Hypercholesterolemia Risk Reduction. Circ Genomic Precis Med. 2022;15(5):e003549. doi:10.1161/CIRCGEN.121.003549

29. Veterans Health Administration, Department of Veterans Affairs. VHA Handbook 1101.10(1): Patient-Aligned Care Team (PACT) Handbook.; 2017. Accessed August 28, 2023. https://www.va.gov/vhapublications/ViewPublication.asp?pub_ID=2977

30. Rosland AM, Wong E, Maciejewski M, et al. Patient-Centered Medical Home Implementation and Improved Chronic Disease Quality: A Longitudinal Observational Study. Health Serv Res. 2018;53(4):2503–2522. doi:10.1111/1475-6773.12805

31. Schuttner L, Wong ES, Rosland AM, Nelson K, Reddy A. Association of the Patient-Centered Medical Home Implementation with Chronic Disease Quality in Patients with Multimorbidity. J Gen Intern Med. 2020;35(10):2932–2938. doi:10.1007/s11606-020-06076-7

32. Bidassie B. The Veterans Affairs Patient Aligned Care Team (VA PACT), a New Benchmark for Patient-Centered Medical Home Models: A Review and Discussion. In: Patient Centered Medicine. IntechOpen; 2017. doi:10.5772/66415

33. Sterling R, Rinne ST, Reddy A, et al. Identifying and Prioritizing Workplace Climate Predictors of Burnout Among VHA Primary Care Physicians. J Gen Intern Med. 2022;37(1):87–94. doi:10.1007/s11606-021-07006-x

34. Scheuner MT, Huynh AK, Chanfreau-Coffinier C, et al. Demographic Differences Among US Department of Veterans Affairs Patients Referred for Genetic Consultation to a Centralized VA Telehealth Program, VA Medical Centers, or the Community. JAMA Netw Open. 2022;5(4):e226687. doi:10.1001/jamanetworkopen.2022.6687

35. Genomic Medicine and Genetic Counseling in the Department of Veterans Affairs and Department of Defense. Frontline Medical Communications. Published August 28, 2019. Accessed August 25, 2023. https://www.frontlinemedcom.com/genomic-medicine-genetic-counseling-in-the-va-and-dod/

36. Price LE, Shea K, Gephart S. The Veterans Affairs’s Corporate Data Warehouse: Uses and Implications for Nursing Research and Practice. Nurs Adm Q. 2015;39(4):311–318. doi:10.1097/naq.0000000000000118

37. Collaboration CTT (CTT). Efficacy and safety of more intensive lowering of LDL cholesterol: a meta-analysis of data from 170 000 participants in 26 randomised trials. The Lancet. 2010;376(9753):1670-1681. doi:10.1016/S0140-6736(10)61350-5

38. Sun YV, Damrauer SM, Hui Q, et al. Effects of Genetic Variants Associated with Familial Hypercholesterolemia on Low-Density Lipoprotein-Cholesterol Levels and Cardiovascular Outcomes in the Million Veteran Program. Circ Genomic Precis Med. 2018;11(12):e002192. doi:10.1161/CIRCGEN.118.002192

39. Kazis LE, Ren XS, Lee A, et al. Health status in VA patients: results from the Veterans Health Study. Am J Med Qual. 1999;14(1):28–38.

40. Selim AJ, Rogers W, Fleishman JA. Updated U.S. population standard for the Veterans RAND 12-item Health Survey (VR-12). Qual Life Res. 2009;18(1):43–52.

41. Horne R, Weinman J, Hankins M. The Beliefs about Medicines Questionnaire: the development and evaluation of a new method for assessing the cognitive representation of medication. Psychol Health. 1999;14(1):1–24.

42. Hibbard JH, Mahoney ER, Stockard J, Tusler M. Development and testing of a short form of the patient activation measure. Health Serv Res. 2005;40(6 Pt 1):1918–1930. doi:10.1111/j.1475-6773.2005.00438.x

43. Kaufman D, Murphy J, Erby L, Hudson K, Scott J. Veterans’ attitudes regarding a database for genomic research. Genet Med. 2009;11(5):329–337.

44. Allen NL, Karlson EW, Malspeis S, Lu B, Seidman CE, Lehmann LS. Biobank participants’ preferences for disclosure of genetic research results: perspectives from the OurGenes, OurHealth, OurCommunity project. Mayo Clin Proc. 2014;89(6):738–746.

45. Vears DF, Minion JT, Roberts SJ, et al. Return of individual research results from genomic research: A systematic review of stakeholder perspectives. PloS One. 2021;16(11):e0258646. doi:10.1371/journal.pone.0258646

46. Carrieri D, Howard HC, Benjamin C, et al. Recontacting patients in clinical genetics services: recommendations of the European Society of Human Genetics. Eur J Hum Genet EJHG. 2019;27(2):169–182. doi:10.1038/s41431-018-0285-1

47. Bookman EB, Langehorne AA, Eckfeldt JH, et al. Reporting genetic results in research studies: Summary and recommendations of an NHLBI working group. Am J Med Genet A. 2006;140A(10):1033-1040. doi:10.1002/ajmg.a.31195

48. Bombard Y, Brothers KB, Fitzgerald-Butt S, et al. The Responsibility to Recontact Research Participants after Reinterpretation of Genetic and Genomic Research Results. Am J Hum Genet. 2019;104(4):578–595. doi:10.1016/j.ajhg.2019.02.025

49. Bredenoord AL, Kroes HY, Cuppen E, Parker M, van Delden JJ. Disclosure of individual genetic data to research participants: the debate reconsidered. Trends Genet. 2011;27(2):41–47.

50. Bredenoord AL, Onland-Moret NC, Van Delden JJ. Feedback of individual genetic results to research participants: in favor of a qualified disclosure policy. Hum Mutat. 2011;32(8):861–867.

51. Wolf SM. Return of individual research results and incidental findings: facing the challenges of translational science. Annu Rev Genomics Hum Genet. 2013;14:557–577.

52. Knoppers BM, Zawati MH, Senecal K. Return of genetic testing results in the era of whole-genome sequencing. Nat Rev Genet. 2015;16(9):553–559.

53. Levesque E, Joly Y, Simard J. Return of research results: general principles and international perspectives. J Law Med Ethics. 2011;39(4):583–592.

54. Harrison SM, Austin-Tse CA, Kim S, et al. Harmonizing variant classification for return of results in the All of Us Research Program. Hum Mutat. 2022;43(8):1114–1121. doi:10.1002/humu.24317

55. Blasimme A, Moret C, Hurst SA, Vayena E. Informed Consent and the Disclosure of Clinical Results to Research Participants. Am J Bioeth. 2017;17(7):58–60. doi:10.1080/15265161.2017.1328532

56. Beshir L. A Framework to Ethically Approach Incidental Findings in Genetic Research. EJIFCC. 2020;31(4):302–309.

57. Vears DF, Minion JT, Roberts SJ, Cummings J, Machirori M, Murtagh MJ. Views on genomic research result delivery methods and informed consent: a review. Pers Med. 2021;18(3):295–310. doi:10.2217/pme-2020-0139

58. Steiner JF, Prochazka AV. The assessment of refill compliance using pharmacy records: methods, validity, and applications. J Clin Epidemiol. 1997;50(1):105–116.

59. Zimolzak AJ, Spettell CM, Fernandes J, et al. Early detection of poor adherers to statins: applying individualized surveillance to pay for performance. PloS One. 2013;8(11):e79611. doi:10.1371/journal.pone.0079611

60. Centers for Medicare and Medicaid Services. Medicare 2019 Part C and D Star Ratings Technical Notes.

61. Horne R, Weinman J, Hankins M. The Beliefs about Medicines Questionnaire: the development and evaluation of a new method for assessing the cognitive representation of medication. Psychol Health. 1999;14(1):1–24.

62. Hibbard JH, Mahoney ER, Stockard J, Tusler M. Development and testing of a short form of the patient activation measure. Health Serv Res. 2005;40(6 Pt 1):1918–1930. doi:10.1111/j.1475-6773.2005.00438.x

63. DiClemente CC, Prochaska JO, Fairhurst SK, Velicer WF, Velasquez MM, Rossi JS. The process of smoking cessation: an analysis of precontemplation, contemplation, and preparation stages of change. J Consult Clin Psychol. 1991;59(2):295–304. doi:10.1037//0022-006x.59.2.295

64. Nigg CR, Burbank PM, Padula C, et al. Stages of change across ten health risk behaviors for older adults. The Gerontologist. 1999;39(4):473–482. doi:10.1093/geront/39.4.473

65. Marcus BH, Selby VC, Niaura RS, Rossi JS. Self-efficacy and the stages of exercise behavior change. Res Q Exerc Sport. 1992;63(1):60–66. doi:10.1080/02701367.1992.10607557

66. Kazis LE, Ren XS, Lee A, et al. Health status in VA patients: results from the Veterans Health Study. Am J Med Qual. 1999;14(1):28–38.

67. Selim AJ, Rogers W, Fleishman JA. Updated U.S. population standard for the Veterans RAND 12-item Health Survey (VR-12). Qual Life Res. 2009;18(1):43–52.

68. Li M, Bennette CS, Amendola LM. The Feelings About genomiC Testing Results (FACToR) Questionnaire: Development and Preliminary Validation. J Genet Couns. 2019;28(2):477–490.

69. Vassy JL, Christensen KD, Schonman EF. The impact of whole-genome sequencing on the primary care and outcomes of healthy adult patients: A pilot randomized trial. Ann Intern Med. 2017;167(3):159–169.

70. Sanders GD, Neumann PJ, Basu A. Recommendations for Conduct, Methodological Practices, and Reporting of Cost-effectiveness Analyses: Second Panel on Cost-Effectiveness in Health and Medicine. JAMA. 2016;316(10):1093–1103.

