## Supplementary File 2 for "Introducing return of results in the Million Veteran Program: Design and pilot results of the MVP-ROAR Familial Hypercholesterolemia Study"

Statistical Analysis Plan

Funding Agency: Department of Veterans Affairs Office of Research and Development, Million Veteran Program

Principal Investigator: Jason L. Vassy, MD, MPH, MS
Biostatistician: Charles Brunette, PhD
Version Number 1.0

September 19, 2023

| Role | Name | Signature | Date |
|---|---|---|---|
| Principal Investigator | Jason Vassy, MD | JASON VASSY 619380 (affiliate) Digitally signed by JASON VASSY 619380 (affiliate) Date: 2023.09.21 07:57:49 -04'00' | |
| Biostatistician | Charles Brunette, PhD | CHARLES BRUNETTE Digitally signed by CHARLES BRUNETTE Date: 2023.09.20 19:54:55 -05'00' | |

### List of Abbreviations

| | |
|---|---|
| ADI | Area deprivation index |
| AE | Adverse event |
| apoB | Apolipoprotein (B) |
| ASCVD | Atherosclerotic cardiovascular disease |
| BMQ | Beliefs about Medicines Questionnaire |
| CDW | Corporate data warehouse |
| CIRB | (VA) Central Institutional Review Board |
| CMS | Centers for Medicare and Medicaid Services |
| DNA | Deoxyribonucleic acid |
| EHR | Electronic health record |
| FACToR | Feelings About Genomic Testing Results Questionnaire |
| FH | Familial hypercholesterolemia |
| IQR | Interquartile range |
| ITT | Intention-to-treat |
| LDL-C | Low-density lipoprotein cholesterol |
| LDLR | Low-density lipoprotein receptor gene |
| Lp(a) | Lipoprotein (a) |
| MCS | Mental component score (MCS) |
| MPR | Medication possession ratio |
| MVP | Million Veteran Program |
| MVP-ROAR-FH | Million Veteran Program Return of Actionable Results – Familial Hypercholesterolemia |
| N | Number of observations / participants |
| PAM-13 | Patient Activation Measure - 13 |
| PCS | Physical component score (VR-12) |
| PDC | Proportion of days covered |
| PI | Principal investigator |
| PCP | Primary care provider |
| RCT | Randomized controlled trial |
| SAE | Serious adverse event |
| SD | Standard deviation |
| VA | Veterans Affairs |
| VR-12 | Veterans RAND 12-Item Health Survey |

### TABLE OF CONTENTS


| | | |
|---|---|---|
| 1.0 | Administrative Information | 4 |
| 2.0 | Introduction | 4 |
| 3.0 | Trial Methods | 5 |
| 4.0 | Trial Population | 8 |
| 5.0 | Statistical Considerations | 11 |
| 6.0 | Outcome definitions and timing | 12 |
| 7.0 | Analysis methods | 15 |
| 8.0 | References | 18 |

### 1.0 Administrative Information

#### 1.1 Trial title and registration

Million Veteran Program Return of Actionable Results - Familial Hypercholesterolemia (MVP-ROAR-FH) Study. ClinicalTrials.gov Identifier: NCT04178122

#### 1.2 Revision history

| SAP version | Section changed | Description | Date amended |
|---|---|---|---|
| 1.0 | | Date created | 09/19/2023 |

#### 1.3 Key personnel

##### 1.3.1 Principal investigator

The principal investigator (PI) supervises all aspects of the study. The PI takes responsibility for the scientific development and conduct of the study, including meeting study goals and timelines, monitoring participant safety, and oversight of the dissemination of research findings.

##### 1.3.2 Biostatistician

The biostatistician advises the study team on the appropriate study design and statistical analysis of study outcomes. The biostatistician conducts and/or reviews sample size and power calculations and provides supervision to the data analyst in performing data collection, data cleaning, and statistical analysis of study data.

##### 1.3.3 Data manager/analyst

The data manager creates and maintains the database housing study data. The data analyst ensures the capture of study data and performs requisite merging and cleaning of study data. The data manager prepares summary data tables for study planning, reporting, monitoring, and dissemination of results. The data manager prepares and maintains participant randomization tables and mechanisms for treatment allocation. The data manager does not engage in the enrollment or allocation of participants to study treatments. Under the direction of the biostatistician, the data analyst may perform statistical analysis of study outcomes.

##### 1.3.4 Research genetic counselor / project manager

The research genetic counselor / project manager is responsible for the day-to-day operations of the MVP-ROAR-FH Study. They also obtain participant consent for study participation and genetic confirmation testing and deliver the intervention to enrolled participants.

### 2.0 Introduction

This document details the proposed data analysis, presentation, and reporting of outcomes associated with the MVP-ROAR-FH Study. The results reported in the primary study manuscript(s) will adhere to the strategy outlined here. All amendments to this plan will be documented and reviewed by the relevant key personnel listed within this document. Any deviations to this plan will be justified and detailed in the final manuscript(s). Further analysis, including subset and exploratory analyses not included here, may occur as needed and will be justified and described if reported. This document follows the published guidelines for the content of statistical analysis plans in clinical trials.[1]

#### 2.1 Background

Familial hypercholesterolemia (FH) is a genetic disorder characterized by elevated low-density lipoprotein cholesterol (LDL-C) levels and increased risk for cardiovascular disease. The MVP-ROAR-FH study aims to return potentially actionable genetic results associated with FH to Million Veteran Program (MVP) participants and their primary care providers. Informing participants of this information might lead to earlier interventions and improved health outcomes.

#### 2.2 Study objectives

The primary objective of MVP-ROAR-FH is to evaluate the difference in change in LDL-C (end-of-study LDL-C minus baseline LDL-C) after 6 months between the Immediate Results arm and the Delayed Results arm. Secondary and exploratory objectives include an assessment of the difference in the proportions of participants reaching LDL-C targets between arms, examination of the difference in the intensification of lipid-lowering pharmacotherapy between arms, and evaluation of medication adherence, cascade testing, lifestyle behaviors, healthcare costs, and quality-of-life after a 6-month observation period.

#### 2.3 Primary outcome and research hypothesis

Primary outcome: Change in LDL-C 6 months after randomization

Null hypothesis: 6-month change in LDL-C is not significantly different between groups receiving their genetic results at baseline compared to those receiving their results after 6 months.

Alternative hypothesis: 6-month change in LDL-C is significantly different between groups receiving their genetic results at baseline compared to those receiving their results after 6 months.

### 3.0 Trial Methods

#### 3.1 Trial design

The MVP-ROAR-FH Study is a randomized controlled trial (RCT). MVP participants suspected of carrying a genetic variant associated with familial hypercholesterolemia are recontacted, enrolled, and, upon receipt of a DNA sample, randomly assigned either to an Immediate Results arm, where the return of genetic results intervention occurs at baseline, or a Delayed Results arm, where the intervention is delivered at the end of the study, after 6 months. The intervention consists of variant confirmation testing and reporting, standard genetic counseling, provision of informational resources to the participant and their primary care providers (PCPs), and documentation of the intervention in the medical record.

3.2 Sample size

The MVP-ROAR-FH Study aims to enroll ten participants into a pilot trial and 244 participants into the RCT.

Sample size is based on the primary outcome of change in LDL-C in each arm after 6 months. Assuming a mean LDL-C reduction of 20% in the Immediate Results arm, a mean LDL-C reduction of 0% in the Delayed Results arm, and a common standard deviation of 30%,[2] 72 total participants (36 per arm) are needed to have 80% power to detect a significant between-group difference at alpha=0.05. Enrollment of twice this number (144 total) will account for an absence of therapy escalation in up to 50% of participants in the Immediate Results arm. Enrollment of 180 total participants will account for up to 20% loss to follow-up.

| **Change in LDL-C at 6 months** | | | **Total sample size required** |
|---|---|---|---|
| **Immediate Results** | **Delayed Results** | **Common SD** | |
| -20% | 0% | 30% | 72 |
| -20% | 0% | 40% | 126 |
| -20% | -5% | 30% | 126 |
| -20% | -5% | 40% | 224 |

An important secondary outcome is the proportion of participants in each arm meeting accepted LDL-C targets at 6 months (<100 mg/dL for primary prevention and <70 mg/dL for secondary prevention). In preparatory-to-research analyses, only 175/322 (46%) MVP participants with a potentially pathogenic LDLR variant had a most recent LDL-C <100mg/dL. To have 80% power to detect a between-arm difference of 20% of participants meeting this LDL-C target at alpha=0.05, up to 194 total participants are needed (97 per arm). To account for up to 20% loss to follow-up, a total of 244 participants (122 per arm) are needed.

| **Proportion of participants with LDL-C < 100mg/dL at 6 months** | | **Total sample size required** |
|---|---|---|
| **Immediate Results** | **Delayed Results** | |
| 10% | 30% | 124 |
| 20% | 40% | 162 |
| 30% | 50% | 186 |
| 40% | 60% | 194 |

3.3 Randomization

Study staff use pre-generated randomization tables for 1:1 allocation of participants to each study arm using a permuted block design with a block size of four. Pre-generated randomization tables are created using standard statistical software (*e.g.* computerized random block and sequence generation) by the MVP-ROAR-FH Study data manager, under the direction of the biostatistician, and stored in a secure file share accessible to select study staff. Randomization occurs upon the completion of baseline procedures and confirmed receipt of each participant's DNA specimen and is mechanized through a computerized randomization tool. Study staff enrolling and allocating participants to study treatments are blinded to the pre-generated randomization tables.

#### 3.4 Data sources, collection, and storage

Study outcomes data will be collected from the VA Corporate Data Warehouse (CDW), a repository of administrative and clinical data from the VA's nationally deployed electronic health record (EHR) system;[3] clinical chart review of the EHR, participant baseline and follow-up surveys, clinical confirmation genetic testing, and trial operations data recorded by the study team. All study data will be stored, cleaned, and analyzed within a secure VA computing environment and will be accessible to authorized study staff only.

#### 3.5 Stopping guidance

This study has no stopping rules. Enrolled participants may withdraw from the study at any time.

#### 3.6 Protocol deviations

Protocol deviations are characterized as circumstances that depart from planned study procedures and anticipated events (*e.g.*, participant withdrawal, loss to follow-up). Protocol deviations may include, but are not limited to, the following:

1. Deviation from inclusion or exclusion criteria (*e.g.*, ineligible patient enrolled and/or randomized)
2. Patient receipt of treatment other than treatment as randomized

The number of ineligible patients randomized, patients receiving a treatment other than as randomized, or other yet to be determined protocol deviations, if any, will be characterized and reported in the final manuscript(s). For the purposes of primary and secondary outcomes, data from patients who experience a protocol deviation will be included in the final data sets. Their outcomes data will be analyzed as part of the treatment group to which they were randomly allocated. The inclusion or exclusion of these patients' data in subsequent secondary or subgroup analyses will be detailed in the final manuscript(s) as needed.

#### 3.7 Adverse events

Adverse events (AEs) related to MVP-ROAR-FH procedures do not include anticipated events related to blood draws (e.g., pain, minor bleeding, bruising, fainting, or lightheadedness) and minor feelings of discomfort while answering survey questions. Pre-existing conditions or illnesses which are expected to exacerbate or worsen are also not considered adverse events and will be accounted for in the subject's medical history. An AE may be considered any other unanticipated or unintended medical occurrence or worsening of a sign or symptom (including an abnormal laboratory finding other than the return of genetic information associated with FH) or disease in a study subject, which does not necessarily have a causal relationship with the study condition, procedure(s) or study agent(s), that occurs after participant informed consent is obtained. A serious adverse event (SAE) will be defined as an AE resulting in one of the following outcomes: death during the 6 months after study enrollment, life threatening event (defined as an event that places a participant at immediate risk of death), inpatient hospitalization, and any other condition which, in the judgment of the PI, represents a significant hazard, such as an important medical event that does not result in one of the above outcomes. An event may be considered an SAE when it jeopardizes the participant or requires medical or surgical intervention to prevent one of the outcomes listed above. AEs may be observed by the study staff or volunteered by participants, their family members, their PCPs, or others. All AEs and SAEs will be assessed for relationship to the study research procedures by the study PI, to determine whether study participation was likely to have caused the AE/SAE.

**4.0 Trial Population**

The overall study population includes all participants of the Million Veteran Program (MVP) mega-biobank research study.

4.1 Study inclusion and exclusion criteria

Inclusion criteria: A participant is eligible for enrollment in this study if he/she meets the following criteria:

- Is a living enrollee in MVP.
- Is identified to have a pathogenic or likely pathogenic variant in an FH-associated gene in their MVP genotype data.
- Has not previously undergone genetic testing for familial hypercholesterolemia. Study staff first ascertain this by review of the medical record and then confirm during the informed consent call by asking the participants about any prior genetic testing he/she has undergone.
- Is not incarcerated.
- Is not pregnant.

4.2 Screening, recruitment, and withdraw

MVP Core study staff queries MVP databases for living participants with an eligible FH variant. The MVP Core study team mails eligible participants a letter introducing this new MVP-related study giving participants the opportunity to opt out of further contact by returning a prepaid opt out postcard or by calling the MVP Call Center. To any participant who does not opt out within 2 weeks of this initial mailing, the MVP-ROAR-FH study team mails a letter providing more detail about the study, including all necessary informed consent information. Two weeks after this mailing, the study genetic counselor calls the participant to review the informed consent information, answers any questions about the study, and documents verbal consent or decline. See MVP-ROAR-FH Study protocol VA CIRB 19-11 for additional detail regarding the study recruitment process.

Duration of the study recruitment period, the total number of patients screened, the number of screened patients not recruited and reason for non-recruitment, and other screening and recruitment metrics will be collected and reported for the overall study. In addition to protocol deviations and AEs or SAEs, if any, the number, and reasons (if known) for participant withdrawal and/or loss to follow-up prior to the conclusion of the study's period of enrollment will be reported in the final manuscript(s). Participant flow will be reported in the final manuscript using CONSORT guidelines for the reporting of clinical trials.[4,5]

4.3 Reference start and end dates

Participant study enrollment occurs on the date of consent. Randomization occurs upon completion of baseline procedures (i.e. baseline survey) and confirmed receipt of DNA biospecimen (blood or saliva). Study participation concludes upon completion of the 6-month biospecimen collection and end-of-study survey, conducted approximately 6 months after the date of randomization and end-of-study survey.

4.3.1 Baseline assessment

For the purposes of outcomes assessment, baseline is defined as the most recent measurement of a study-related outcome (see Section 6.0) on or prior to a participant's date of randomization, unless otherwise specified. The total number of participants with baseline measurements obtained post-randomization and/or any statistical analysis including a baseline measurement obtained post-randomization will be reported.

4.3.2 End-of-study follow-up and period of observation

Enrolled participants will be observed for a total of 6 months. For the primary (change in LDL-C) and secondary (proportion meeting LDL-C targets) outcomes related to LDL-C, the end-of-study date will correspond to the date 6 months from the date of randomization. The most recent LDL-C value on or after this date, either associated with a completed study-related blood draw or LDL-C value extracted from CDW data (if unable to obtain an end-of-study specimen), will be used as the 6-month LDL-C value.

Similarly, for the secondary outcome associated with intensification of pharmacotherapy, the end-of-study date will correspond to the date 6 months from the date of randomization. All lipid-lowering related prescriptions, either derived from participant survey responses or extracted from CDW data, which are identified as active prior to or after the date of randomization and considered active either on or before the date 6 months from the date of randomization will be considered for analysis. Prescriptions are defined as active if they fall within a time window of one and a half times the total days supply (equivalent to a medication possession ratio of 0.67) from the prescription start date.[6–8]

4.4 Analysis populations

4.4.1 Intention-to-treat populations

Intention-to-treat (ITT) populations include all patients who undergo randomization and are characterized by the treatment they were randomized to receive (Immediate Results vs. Delayed Results).

4.4.2 Complete case populations

The complete case populations consist of patients who undergo randomization and complete all study assessments, including both the baseline and end-of-study patient surveys and end-of-study LDL-C measurement.

4.4.3 Subgroup populations

Additional subgroups of the study population may include the analyses of patients stratified by demographic (e.g. age), sex, or baseline LDL-C values. Each subgroup population will be described in detail as reported in the final study manuscript(s).

4.5 Baseline patient characteristics

Baseline characteristics will be summarized and presented for participants in the ITT populations. Standard statistical summaries, depending on data type and distribution, will be presented as 1) total numbers of participants with each characteristic (n) and as a proportion (%) of each group stratified by randomization arm or 2) as means and standard deviations (if normally distributed) or medians and interquartile ranges (IQR) (if non normally distributed) stratified by randomization arm. No statistical testing will be carried out for participant baseline characteristics or measures between treatment groups.

At minimum, the below participant baseline characteristics, including the pre-specified baseline measurements of the study outcomes described in Section 6.0, will be derived and reported:

| **Baseline characteristic** | **How derived** | **Presentation** |
|---|---|---|
| Age in years | As calculated using EHR administrative data relative to date of consent and enrollment. | mean (SD) / median (IQR) |
| Gender / Sex | As determined by EHR administrative data. | n (%) |
| Race | As determined by EHR administrative data and/or data collected from baseline survey. | n (%) |
| Ethnicity | As determined by EHR administrative data and/or data collected from baseline survey. | n (%) |
| Socioeconomic Status / Area Deprivation Index | Calculated using income, geographic, and other EHR administrative data.(ADI)[9] | n (%) |
| Self-reported health status and quality of life | As determined by data collected from baseline survey. (VR-12)[10–12] | mean (SD) / median (IQR) |
| Self-reported patient activation | As determined by data collected from baseline survey. (PAM-13)[13] | mean (SD) / median (IQR) / n (%) |
| Beliefs about medications | As determined by data collected from baseline survey. (Beliefs About Medicines Questionnaire)[14] | mean (SD) / median (IQR) |
| Low-density lipoprotein cholesterol | As determined by EHR data. | mean (SD) / median (IQR) |
| Lipid-lowering pharmacotherapy | As determined by data collected from baseline survey and EHR data. | n (%) |

### 5.0 Statistical Considerations

5.1 Statistical framework

The principal analysis uses an intention-to-treat (ITT)[15] approach to compare the Immediate Results and Delayed Results arms. Using an independent t-test, analysis of the primary outcome uses a t-statistic, and two-sided type I error rate of 0.05 to test the null hypothesis of no difference in LDL-C change between arms. To quantify the treatment arm difference for the primary endpoint, a mean difference will be presented with a corresponding confidence interval estimate. Mean change in LDL-C is also reported separately for each arm. Similarly, for secondary and other pre-specified outcomes an ITT approach is used to make outcomes comparisons across treatment groups. Subset analyses, including sensitivity and group analyses by demographic, variant confirmation status, or other study or patient characteristics, are considered exploratory and will be described in detail if reported.

5.2 Interim analyses

No formal interim hypothesis testing is planned.

5.3 Timing of final analyses

Final analyses of the MVP-ROAR-FH Study data are conducted upon the conclusion of the final participant's data collection procedures (*i.e.*, end-of-study survey and biospecimen collection).

5.4 Confidence intervals, *P* values, and multiple testing

All statistical testing is reported with an effect, a two-sided 95% confidence interval, and *P* value, unless otherwise specified. *P* values less than 0.05 will be reported as significant for the primary outcome. *P* values reported as significant for secondary outcomes will undergo Bonferroni correction for multiple hypothesis-testing. Other pre-specified and post-hoc analyses are considered exploratory.

5.5 Missing data

Prior to statistical analysis, outcomes data are reviewed for the amount and pattern of data missingness (*e.g.*, missing at random) using standard statistical software and methods. For outcomes analysis, partially observed outcomes may be imputed using mean or median imputation, multiple imputation, or comparable methods, as appropriate.[16–18] Any necessary imputation will be conducted separately within each treatment arm. Proportions of data missingness, reasons for missingness (if known), and methods used for data imputation if required, including number of imputations and sensitivity analyses performed, will be reported in the final manuscript(s).

5.6 Statistical assumptions and issues

Prior to analysis, statistical assumptions are evaluated for each proposed outcome assessment. The presence of distributional assumptions, influential outliers, and homogeneity of variance, among other common assumptions related to the analyses described here, are assessed. Methods used and results of the assessment of statistical assumptions will be acknowledged in the final study manuscript(s). In addition, issues related to significant differences between withdrawn, lost to follow-up, and remaining cases as well as changes in study methods over time (e.g. change in study procedures that result in materially different patient outcomes) will be considered. A description of unusual outliers, violated assumptions, or other issues that may impact the integrity of the analyses, and any corrective action (e.g. assumptions evaluated, review of outliers/sensitivity analyses, variable transformations, etc.) will be described in the final study manuscript(s) as applicable.

5.7 Clustered data

Given the small number of participants relative to the number of VA PCPs and facilities nationally, there is little potential for clustering effect among patients receiving care from the same PCPs or at the same VA facility. As a result, provider or facility clustering will not be considered in the final analyses.

5.8 Statistical software

Statistical analysis will be conducted using appropriate and validated software, including SAS, STATA, R, or other comparable statistical programs. The applicable software(s), package(s), and version(s) used for the analyses of study data will be reported in the final manuscript(s).

### 6.0 Outcome definitions and timing

6.1 Primary outcome (6-month difference in LDL-C)

Baseline (at enrollment) and end-of-study (at 6 months) LDL-C values are defined as the LDL-C measurements obtained from study-related blood draws as processed by the study's centralized laboratory. For participants for whom it is not feasible to visit their local VA facility for a blood draw at baseline, a documented LDL-C value in their medical record may be used for a study measurement, provided it is not older than 6 months from their date of enrollment and a change in lipid-lowering therapy has not occurred between the clinical LDL-C value and study enrollment. For participants for whom it is not feasible to visit their local VA facility for an end-of-study blood draw, a documented LDL-C value in their medical record may be used for a study measurement. The documented LDL-C value closest to the date 6 months after the date of randomization will be used for the end-of-study value. If no 6-month value is available, the study may use an LDL-C value obtained from the medical record during the 6-month observation period, including carry forward of the baseline value.

6.2 Secondary outcomes

6.2.1. Proportion meeting LDL-C targets

The proportion of participants meeting individualized clinically significant LDL-C targets (<100mg/dL for primary prevention and <70 mg/dL for secondary prevention) will be determined at 6 months, using the LDL-C values as described for the primary outcome in Section 6.1.

Secondary prevention will be defined as any patient with any of the following:

> Pre-existing atherosclerotic disease (ASCVD): acute coronary syndrome in the prior 12 months, history of myocardial infarction, history of ischemic stroke, symptomatic peripheral artery disease including aneurysm, all of atherosclerotic origin.

Presence of any ASCVD risk factors: age ≥65 years; prior percutaneous coronary intervention; prior coronary artery bypass graft; other evidence of coronary artery disease; diabetes mellitus; hypertension; chronic kidney disease; current smoking; congestive heart failure; family history of premature ASCVD; elevated coronary artery calcium score, Lp(a), or apoB; or ankle-brachial index <0.9.

All other participants will be considered eligible for primary prevention.

The primary or secondary prevention status of each participant will be determined by clinician review of participant medical records through the end-of-study date. The clinician reviewer(s) will be blinded to randomization status. For each participant deemed eligible for secondary prevention, the clinician will record the reason(s) for eligibility in a study database.

6.2.2. Intensification of pharmacotherapy

The proportion of participants with an intensification of lipid-lowering pharmacotherapy will be determined from baseline and end-of-study prescription data, obtained from the CDW, medical record review, and patient surveys. Intensification of pharmacotherapy at 6-months will be a composite outcome including prescription of new monotherapy, dose escalation of existing pharmacotherapy, or addition of one or more medications to existing pharmacotherapy compared to baseline pharmacotherapy status.

6.3 Other prespecified outcomes

6.3.1. Medication adherence

Continuous medication adherence is assessed using medication possession ratios (MPRs) or proportion of days covered (PDC) as derived from CDW pharmacy data.[6–8] All lipid-lowering related prescriptions, which are identified as active after the date of randomization and considered active either on or before the date 6 months from the date of randomization will be considered for analysis. PDC is calculated as the percentage of days in which a participant has access to a prescribed medication over the period of observation (assessed as sum of total days supply / total days in observation period). A measure of medication adherence is derived for each participant with a lipid-lowering medication prescription during the study observation period between the date of randomization and the date 6 months after randomization. Participants with a PDC of 80% or greater associated with lipid-lowering medications will be considered adherent. Total numbers of participants with lipid-lowering prescriptions, as well as subsets of participants with PDC measures will be reported in the final manuscript(s).

6.3.2. Cascade testing

The number of first-degree relatives undergoing genetic testing within 6 months will be recorded for each participant, as measured on the end-of-study survey.

6.3.3. Self-reported quality of life

Self-reported quality of life is assessed via the baseline and end-of-study surveys, using the Veterans RAND 12-Item Health Survey (VR-12).[10–12] The VR-12 computes two continuous composite scores, a physical component summary (PCS) and a mental component summary (MCS).

6.3.4. Lifestyle behaviors

Lifestyle behaviors are assessed using items structured on the transtheoretical model of behavior change.[19] Response options seek to assess at what point participants are in the behavior change process at 6 months. Response options assess readiness for change across myriad lifestyle behaviors using response options ranging from engaging in a specific behavior for more than 6 months (maintenance stage) to not at all (precontemplation stage).

a) Self-reported physical activity[20]

Self-reported physical activity is assessed with the baseline and end-of-study surveys, using the single-item question "Do you exercise 3 times a week for at least 20 minutes each time?"

b) Self-reported smoking status[21,22]

Self-reported smoking status is assessed with the baseline and end-of-study surveys, using two items: "Are you currently a smoker?" and, if yes, "Are you seriously thinking of quitting smoking?"

c) Self-reported saturated fat consumption[19]

Self-reported saturated fat consumption is assessed with the baseline and end-of-study surveys, using the single-item question "Do you consistently avoid eating high fat foods?"

6.3.5. Healthcare costs and utilization

A combination of administrative data, survey data, and microcosting approaches are used to estimate costs over the 6 months after randomization. Utilization of healthcare services, including laboratory tests, office visits, time demands, transportation cost, and hospitalization information will be derived from both end-of-study participant surveys and CDW administrative data.[23,24] Estimates of the infrastructure and personnel needed to deliver the intervention are derived empirically from the study. Healthcare costs are abstracted from billing and administrative data from the CDW and CMS data.

6.4 <u>Other exploratory outcomes</u>

6.4.1. Self-reported patient activation

Self-reported understanding, competence, and willingness to participate in health care decisions and processes are assessed via the baseline and end-of-study surveys, using the 13-item short form of the Patient Activation Measure (PAM-13).[13] Each PAM-13 item has four possible response options: "*Strongly disagree*" (1), "*Disagree*" (2), "*Agree*" (3), "*Strongly agree*" (4), as well as "*Does not apply*" (0). Response values are summed, divided by the total number of items responded to (excluding selections of non-applicable items), and multiplied by 13. The raw score is converted using a scoring table to derive both a linear score from 0 (no activation) to 100 (fully activated) and interval patient activation scores (1: activation not important, passive recipient of care; 2: lack of knowledge or confidence to take action; 3: beginning to take action; 4 taking action).

6.4.2. Beliefs about medications

Beliefs about medications is assessed on the baseline and end-of-study surveys, using the 8-item Beliefs About Medicines Questionnaire (BMQ) - General Scale.[14] Each item has five possible response options: (1) *Strongly disagree*, (2) *Disagree*, (3) *Neither agree nor disagree*, (4) *Agree*, (5) *Strongly agree*. A total score reflecting overall beliefs is calculated as a numerical sum across participant responses ranging from 8 to 40. Higher scores represent stronger beliefs about medication overuse and harm. The general use subscale is calculated as a numerical sum using items 1, 3, 4, and 8 ranging from 4 to 20 and represents beliefs about medication overuse. The general harm subscale is calculated as a numerical sum using items 2, 5, 6, and 7 ranging from 4 to 20 and represents beliefs about medication harm. A measure of medication beliefs is derived for each participant at baseline and 6 months after randomization.

6.4.3. Self-reported feelings about genomic testing results

Self-reported feelings about the psychosocial impact of receiving genomic test results will be assessed using the Feelings About genomic Testing Results (FACToR) Questionnaire.[25] The FACToR is only administered to the Immediate Results arm.

6.4.4. Preferences for receiving genetic test results

Preferences for receiving genetic test results include two items, developed specifically for this study, to assess 1) preferred first contact for genetic test results (e.g., participant, participant's primary care provider) and 2) preferred provider to deliver results (e.g., research genetic counselor, participant's primary care provider). These items are only administered to the Immediate Results arm.

### 7.0 Analysis methods

7.1 <u>Covariate adjustment</u>

No prespecified covariate adjustment is planned. If any covariate adjustment is deemed necessary during analysis or used to improve the precision of estimates, rationale and methods will be fully described in the final manuscript(s).

7.2 <u>Primary outcome</u>

The difference in LDL-C reduction after 6 months between the Immediate Results arm and the Delayed Results arm will be assessed. The LDL-C values at baseline and 6 months will first be summarized descriptively for each arm. The change in LDL-C from baseline to 6 months will be calculated for each participant. An independent *t*-test will be employed to compare the mean LDL-C change between the two arms, assuming normally distributed data. If the change scores do not follow a normal distribution, a non-parametric alternative, the Wilcoxon rank-sum test, may be used. The effect size (Cohen's d), confidence intervals, and *P* value will be reported to determine the statistical significance of the observed difference. Additionally, potential confounders may be included in regression modeling to ensure the robustness of findings.

7.3 <u>Secondary outcomes</u>

7.3.1 Proportion meeting LDL-C targets

Proportions of patients meeting individualized cholesterol targets at end of study (yes or no for <100mg/dL for primary prevention and <70 mg/dL for secondary prevention) will be compared between the Immediate Results and Delayed Results arm using either a Z-test of proportions or chi-squared test. In the event of small cell counts, we will employ Fisher's exact test to ensure the validity of the results. We will compute the relative risk and 95% confidence intervals to quantify the strength and direction of the association. Potential confounders may be adjusted for by using logistic regression. Frequencies and proportions of participants meeting LDL-C targets will be reported by randomization arm.

7.3.2 Intensification of pharmacotherapy

Proportion of patients with intensification of pharmacotherapy from baseline to end of study will be compared between the Immediate Results and Delayed Results arm using either a Z-test of proportions or chi-squared test. In the event of small cell counts, we will employ Fisher's exact test to ensure the validity of the results. We will compute the relative risk and 95% confidence intervals to quantify the strength and direction of the association. Potential confounders may be adjusted for by using logistic regression. Frequencies and proportions of the intensification of medications among participants will be reported by randomization arm. Exploratory analyses may compare participant medication trajectories between groups, including intensification, unchanged prescription status, or deintensification status at baseline, 3 months, and 6 months using a generalized linear model fit with generalized estimating equations.[26]

7.4 Other prespecified and exploratory outcomes

7.4.1 Medication adherence

Proportions of patients considered adherent to lipid-lowering medication prescriptions (PDC ≥ 80%) are compared between the Immediate Results and Delayed Results arm using either a Z-test of proportions or chi-squared test. In the event of small cell counts, we will employ Fisher's exact test to ensure the validity of the results. We will compute the relative risk and 95% confidence intervals to quantify the strength and direction of the association.

7.4.2 Cascade testing

The number of first-degree relatives undergoing genetic testing within 6 months will be recorded. Mean numbers and standard deviations of first-degree relatives tested will be calculated for each arm. A Poisson regression or negative binomial regression (in case of overdispersion) will be used to compare the rates of cascade testing between the two study arms. Potential confounders may be included in regression modeling to ensure the robustness of findings.

7.4.3 Self-reported quality of life

Mean change scores (6-month scores minus baseline scores) and standard deviations for both PCS and MCS of the VR-12 will be calculated for each arm. Independent samples t-tests will compare differences in mean change between the two arms for both PCS and MCS. Multivariable linear regression models (e.g. ANCOVA) may be fit to account for potential confounding variables.

7.4.4 Healthcare costs

Healthcare costs will be described using means and standard deviations for both arms. The differences in mean healthcare costs between the two groups will be evaluated using a generalized linear model with appropriate distributional assumptions (e.g., gamma for right skewed cost data). Additional covariates may be included to obtain more precise estimates or to adjust for potential confounding variables. Treatment effect is characterized by treatment versus control arm, presented as mean follow-up estimates with accompanying standard errors, 95% confidence intervals and *P* values. In the presence of substantial missing data, mixed modeling or other repeated measures designs may be implemented.

7.4.5 Categorical and ordinal outcome measures

The following measures are compared between treatment groups among the ITT populations using standard methods for categorical data analysis:[27,28]

- Self-reported physical activity
- Self-reported smoking status
- Self-reported saturated fat consumption
- Self-reported patient activation (categorical score)
- Self-reported feelings about genomic testing results (Immediate Results arm only, items)
- Preferences for receiving genetic test results (Immediate Results arm only)

Frequency of end-of-study self-reported responses to study surveys, including categorical, ordinal, and Likert items, will be reported by treatment group (n, %). Binary logistic regression will be used to compare end-of-study dichotomous outcomes between treatment groups. To assess post- treatment ordered outcomes (e.g., lifestyle behaviors along the transtheoretical model continuum) between treatment arms, we will use ordinal logistic regression (e.g., cumulative logit model). Initial models will include treatment group assignment, and when available, baseline measures as covariates. Treatment effect is characterized by an odds ratio estimate presented with standard error, 95% confidence interval, and *P* value.

7.4.6 Continuous outcome measures

The following continuous measures are compared between treatment groups among the ITT populations using standard linear methods:[29,30]

- Self-reported patient activation (linear score)
- Self-reported beliefs about medications
- Self-reported feelings about genomic testing results (Immediate Results arm only, score)

Mean and standard deviation (or median and IQR, if applicable) of continuous scores associated with end-of-study self-reported responses to study surveys will be reported by treatment group. Analysis of covariance (ANCOVA) is used to compare continuous follow-up measures, including participant baseline measures, when available, and treatment group assignment. Treatment effect is characterized by treatment versus control arm, presented as mean follow-up estimates with accompanying standard errors, 95% confidence intervals and *P* values. In the presence of substantial missing data, mixed modeling or other repeated measures designs may be implemented.

7.5 Additional analyses

Further exploratory analyses using the methods described may be conducted for all study outcomes between treatment arms and across relevant subgroups. Inclusion of additional covariates in the models described or use of alternative statistical methods may be implemented to enhance model precision, to adjust for differences in baseline factors or multilevel characteristics, or to improve the integrity of the analyses (e.g., in the event of substantial missing data), among other reasons. To assess robustness of ITT analyses, analyses may be replicated within the relevant complete case populations. The addition of covariates or use of alternative methods to assess primary, secondary, and other outcomes may be considered and are supplemental to the prespecified analyses.

Additional exploratory analyses may be conducted to further examine study data or address research questions that arise during the conduct of the study. Any exploratory analyses or use of alternative methods will be justified and described in detail if reported.

#### 8.0 References


1. Gamble C, Krishan A, Stocken D, et al. Guidelines for the Content of Statistical Analysis Plans in Clinical Trials. *JAMA*. 2017;318(23):2337-2343. doi:10.1001/jama.2017.18556
2. Lewis SJ, Olufade T, Anzalone DA, Malangone-Monaco E, Evans KA, Johnston S. LDL cholesterol levels after switch from atorvastatin to rosuvastatin. *Curr Med Res Opin*. 2018;34(10):1717-1723. doi:10.1080/03007995.2017.1421147
3. Price LE, Shea K, Gephart S. The Veterans Affairs's Corporate Data Warehouse: Uses and Implications for Nursing Research and Practice. *Nurs Adm Q*. 2015;39(4):311-318. doi:10.1097/NAQ.0000000000000118
4. Schulz KF, Altman DG, Moher D, the CONSORT Group. CONSORT 2010 Statement: updated guidelines for reporting parallel group randomised trials. *Trials*. 2010;11(1):32. doi:10.1186/1745-6215-11-32
5. Butcher NJ, Monsour A, Mew EJ, et al. Guidelines for Reporting Outcomes in Trial Reports: The CONSORT-Outcomes 2022 Extension. *JAMA*. 2022;328(22):2252-2264. doi:10.1001/jama.2022.21022
6. Steiner JF, Prochazka AV. The assessment of refill compliance using pharmacy records: Methods, validity, and applications. *J Clin Epidemiol*. 1997;50(1):105-116. doi:10.1016/S0895-4356(96)00268-5
7. Zimolzak AJ, Spettell CM, Fernandes J, et al. Early Detection of Poor Adherers to Statins: Applying Individualized Surveillance to Pay for Performance. *PLoS One*. 2013;8(11):e79611. doi:10.1371/journal.pone.0079611
8. Centers for Medicare and Medicaid Services. *Medicare 2023 Part C &D Star Ratings Technical Notes*.; 2023. Accessed September 1, 2023. https://www.cms.gov/files/document/2023-star-ratings-technical-notes.pdf
9. Kind AJH, Buckingham WR. Making Neighborhood-Disadvantage Metrics Accessible — The Neighborhood Atlas. *N Engl J Med*. 2018;378(26):2456-2458. doi:10.1056/NEJMp1802313
10. Jones D, Kazis L, Lee A, et al. Health Status Assessments Using the Veterans SF-12 and SF-36: Methods for Evaluating Outcomes in the Veterans Health Administration. *J Ambulatory Care Manage*. 2001;24(3):68-86. doi:10.1097/00004479-200107000-00011
11. Kazis LE, Selim A, Rogers W, Ren XS, Lee A, Miller DR. Dissemination of methods and results from the veterans health study: final comments and implications for future monitoring strategies within and outside the veterans healthcare system. *J Ambulatory Care Manage*. 2006;29(4):310-319. doi:10.1097/00004479-200610000-00007
12. Selim AJ, Rogers W, Fleishman JA, et al. Updated U.S. population standard for the Veterans RAND 12-item Health Survey (VR-12). *Qual Life Res*. 2009;18(1):43-52. doi:10.1007/s11136-008-9418-2
13. Hibbard JH, Mahoney ER, Stockard J, Tusler M. Development and Testing of a Short Form of the Patient Activation Measure. *Health Serv Res*. 2005;40(6p1):1918-1930. doi:10.1111/j.1475-6773.2005.00438.x
14. Horne R, Weinman J, Hankins M. The beliefs about medicines questionnaire: The development and evaluation of a new method for assessing the cognitive representation of medication. *Psychol Health*. 1999;14(1):1-24. doi:10.1080/08870449908407311
15. Lachin JM. Statistical Considerations in the Intent-to-Treat Principle. *Control Clin Trials*. 2000;21(3):167-189. doi:10.1016/S0197-2456(00)00046-5
16. Spiro A, Rogers W, Qian S, Kazis E. Imputing Physical and Mental Summary Scores (PCS and MCS) for the Veterans SF-12 Health Survey in the Context of Missing Data. In: *Technical*

*Report, Scoring Algorithms and Users Guide, Submitted and Approved by the Center for Medicare and Medicaid Services (CMS) and National Committee for Quality Assurance (NCQA)*. ; 2004.
17. Lee KJ, Carlin JB. Multiple Imputation for Missing Data: Fully Conditional Specification Versus Multivariate Normal Imputation. *Am J Epidemiol*. 2010;171(5):624-632. doi:10.1093/aje/kwp425
18. Li P, Stuart EA, Allison DB. Multiple Imputation: A Flexible Tool for Handling Missing Data. *JAMA*. 2015;314(18):1966-1967. doi:10.1001/jama.2015.15281
19. Nigg CR, Burbank PM, Padula C, et al. Stages of change across ten health risk behaviors for older adults. *The Gerontologist*. 1999;39(4):473-482. doi:10.1093/geront/39.4.473
20. Marcus BH, Selby VC, Niaura RS, Rossi JS. Self-efficacy and the stages of exercise behavior change. *Res Q Exerc Sport*. 1992;63(1):60-66. doi:10.1080/02701367.1992.10607557
21. DiClemente CC, Prochaska JO, Fairhurst SK, Velicer WF, Velasquez MM, Rossi JS. The process of smoking cessation: an analysis of precontemplation, contemplation, and preparation stages of change. *J Consult Clin Psychol*. 1991;59(2):295-304. doi:10.1037//0022-006x.59.2.295
22. Velicer WF, Fava JL, Prochaska JO, Abrams DB, Emmons KM, Pierce JP. Distribution of smokers by stage in three representative samples. *Prev Med*. 1995;24(4):401-411. doi:10.1006/pmed.1995.1065
23. Sanders GD, Neumann PJ, Basu A, et al. Recommendations for Conduct, Methodological Practices, and Reporting of Cost-effectiveness Analyses: Second Panel on Cost-Effectiveness in Health and Medicine. *JAMA*. 2016;316(10):1093-1103. doi:10.1001/jama.2016.12195
24. Vassy JL, Christensen KD, Schonman EF, et al. The Impact of Whole-Genome Sequencing on the Primary Care and Outcomes of Healthy Adult Patients: A Pilot Randomized Trial. *Ann Intern Med*. 2017;167(3):159-169. doi:10.7326/M17-0188
25. Li M, Bennette CS, Amendola LM, et al. The Feelings About genomiC Testing Results (FACToR) Questionnaire: Development and Preliminary Validation. *J Genet Couns*. 2019;28(2):477-490. doi:10.1007/s10897-018-0286-9
26. Touloumis A. R Package multgee: A Generalized Estimating Equations Solver for Multinomial Responses. *J Stat Softw*. 2015;64:1-14. doi:10.18637/jss.v064.i08
27. Agresti A. *Categorical Data Analysis*. 3rd ed. Wiley-Interscience; 2013.
28. Harrell FE. *Regression Modeling Strategies With Applications to Linear Models, Logistic and Ordinal Regression, and Survival Analysis*. 2nd ed. Springer International Publishing; 2015.
29. Vickers AJ, Altman DG. Statistics notes: Analysing controlled trials with baseline and follow up measurements. *BMJ*. 2001;323(7321):1123-1124. doi:10.1136/bmj.323.7321.1123
30. O'Connell NS, Dai L, Jiang Y, et al. Methods for Analysis of Pre-Post Data in Clinical Research: A Comparison of Five Common Methods. *J Biom Biostat*. 2017;8(1):1-8. doi:10.4172/2155-6180.1000334
