## Supplementary File 3 for "Introducing return of results in the Million Veteran Program: Design and pilot results of the MVP-ROAR Familial Hypercholesterolemia Study"

VA | U.S. Department of Veterans Affairs

**Million Veteran Program**
Washington, DC 20420

MVP ROAR Return Of Actionable Results

### MVP-ROAR 6-MONTH SURVEY

"Hello, this is [STUDY STAFF NAME] from the MVP-ROAR study. May I please speak to [PARTICIPANT NAME]?"

"Good morning/afternoon/evening. You recall that you enrolled in the MVP-ROAR study about 6 months ago. I'm now calling to see if this would be a good time for you to complete your follow-up survey with me over the phone. This will take about 20 minutes. Is this a good time for us to complete the survey?"

[IF NO] "When would be another day and time when it would be more convenient for us to call you back for the survey?"

"Thank you. This survey will ask you a few questions about yourself and your health care. You answered some of these questions 6 months ago, and we want to see if your responses to those questions have changed. You may choose to skip any of these questions that you do not wish to answer."

#### 1. MEDICATIONS

"Before administering the survey, I'd like to ask you about the medications you take. I am looking at a list of your VA prescriptions. Can you confirm if you are currently taking the following medications?"

[Study staff reads the name of each medication with an active VA prescription one at a time, noting each medication as *Taking*, *Not Taking*, or *I'm Not Sure/I Don't Know*.]

"Are there any other prescription medications you take, either from a VA provider or from a provider outside of the VA?"

[Study staff notes name, dose, and frequency of any medication the participant identifies. If participant does not know or have all of this information, study staff may ask the participant to call back with the information at a later time.]

"Are there any over-the-counter medications, vitamins, or other supplements that you take?"

[Study staff notes name, dose, and frequency of any medication the participant identifies. If participant does not know or have all of this information, study staff may ask the participant to call back with the information at a later time. Once collected, study staff may complete the remaining portions of the survey by phone. The below survey may also be administered in paper form by participant request.]

**< SURVEY BEGINS ON NEXT PAGE >**

**MVP-ROAR 6-MONTH SURVEY**

**PARTICIPANT ID:_______________________**

#### 1. HEALTH BEHAVIORS

The following statements are about your health behaviors. Please state whether you have been doing the following health behaviors consistently in the last 6 months.

| | YES, I have been for MORE than 6 months | YES, I have been, but for less than 6 months | NO, but I intend to in the next 30 days | NO, but I intend to in the next 6 months | NO, and I do NOT intend to in the next 6 months |
|---|---|---|---|---|---|
| Have you been trying to lose weight? | ❑ | ❑ | ❑ | ❑ | ❑ |
| Do you limit the calories you eat each day? | ❑ | ❑ | ❑ | ❑ | ❑ |
| Do you limit your sugar intake? | ❑ | ❑ | ❑ | ❑ | ❑ |
| Do you consistently avoid eating high fat foods? (e.g., butter, cheese, processed meats) | ❑ | ❑ | ❑ | ❑ | ❑ |
| Have you been eating a diet high in fiber? (e.g., fruits, vegetables, grains) | ❑ | ❑ | ❑ | ❑ | ❑ |
| Do you exercise 3 times a week for at least 20 minutes each time? | ❑ | ❑ | ❑ | ❑ | ❑ |
| Do you take vitamins, minerals, herbs or other supplements? | ❑ | ❑ | ❑ | ❑ | ❑ |
| Do you limit the amount of alcohol you drink? | ❑ | ❑ | ❑ | ❑ | ❑ |
| Have you been trying to reduce the stress you experience? | ❑ | ❑ | ❑ | ❑ | ❑ |

**Are you currently a smoker?**
- ❑ YES, I currently smoke.
- ❑ NO, I quit within the last 6 months.
- ❑ NO, I quit more than 6 months ago.
- ❑ NO, I have never smoked.

**[IF CURRENTLY SMOKING]: Are you seriously thinking of quitting smoking?**
- ❑ YES, within the next 30 days.
- ❑ YES, within the next 6 months.
- ❑ NO, not thinking of quitting.

**2. SELF-RELATED HEALTH AND QUALITY OF LIFE**

The following questions ask about how you feel and how well you are able to do your usual activities. If you are unsure how to answer a question, please give the best answer you can.

**1. In general, would you say your health is...?**

[] Excellent [] Very Good [] Good [] Fair [] Poor

**2. The following questions are about activities you might do during a <u>typical day</u>. Does your health now limit you in these activities? If so, how much?**

**a. Moderate activities, such as moving a table, pushing a vacuum cleaner, bowling, or playing golf?**

[] Yes, limited a lot [] Yes, limited a little [] No, not limited at all

**b. Climbing several flights of stairs?**

[] Yes, limited a lot [] Yes, limited a little [] No, not limited at all

**3. During the <u>past 4 weeks</u>, have you had any of the following problems with your work or other regular daily activities as a result of <u>your physical health</u>?**

**a. Accomplished less than you would like?**

[] No, none of the time
[] Yes, a little of the time
[] Yes, some of the time
[] Yes, most of the time
[] Yes, all of the time

**b. Were limited in the kind of work or other activities?**

[] No, none of the time
[] Yes, a little of the time
[] Yes, some of the time
[] Yes, most of the time
[] Yes, all of the time

**4. During the <u>past 4 weeks</u>, have you had any of the following problems with your work or other regular daily activities as a result of <u>any emotional problems</u> (such as feeling depressed or anxious)?**

**a. Accomplished less than you would like?**

[] No, none of the time
[] Yes, a little of the time
[] Yes, some of the time
[] Yes, most of the time
[] Yes, all of the time

**b. Didn't do work or other activities as carefully as usual?**

[] No, none of the time
[] Yes, a little of the time
[] Yes, some of the time
[] Yes, most of the time
[] Yes, all of the time

**5. During the <u>past 4 weeks</u>, how much did pain interfere with your normal work (including both work outside the home and house work)?**

[] Not at all [] A little bit [] Moderately [] Quite a bit [] Extremely

These questions are about how you feel and how things have been with you during the past 4 weeks. For each question, please give the one answer that comes closest to the way you have been feeling.

**6. How much of the time during the past 4 weeks:**

**a. Have you felt calm and peaceful?**

[] All of the time
[] Most of the time
[] A good bit of the time
[] Some of the time
[] A little of the time
[] None of the time

**b. Did you have a lot of energy?**

[] All of the time
[] Most of the time
[] A good bit of the time
[] Some of the time
[] A little of the time
[] None of the time

**c. Have you felt downhearted and blue?**

[] All of the time
[] Most of the time
[] A good bit of the time
[] Some of the time
[] A little of the time
[] None of the time

**7. During the past 4 weeks, how much of the time has your physical health or emotional problems interfered with your social activities (like visiting with friends, relatives, etc.)?**

[] All of the time
[] Most of the time
[] Some of the time
[] A little of the time
[] None of the time

Now, we'd like to ask you some questions about how your health may have changed.

**8. <u>Compared to one year ago</u>, how would you rate your physical health in general now?**

[] Much better
[] Slightly better
[] About the same
[] Slightly worse
[] Much worse

**9. <u>Compared to one year ago</u>, how would you rate your emotional problems (such as feeling anxious, depressed, or irritable) now?**

[] Much better
[] Slightly better
[] About the same
[] Slightly worse
[] Much worse"

**3. PATIENT ACTIVATION**

The following are statements that people sometimes make when they talk about their health. Please indicate how much you disagree or agree with each statement as it applies to you personally. Your answers should be what is true for you and not just what you think others expect of you. Your choices are *Strongly Disagree, Disagree, Agree,* or *Strongly Agree*. If the statement does not apply to you, please say *Does Not Apply*.

**1. When all is said and done, I am the person who is responsible for managing my health.**

[] Strongly Disagree [] Disagree [] Agree [] Strongly Agree [] Does Not Apply

**2. Taking an active role in my own health care is the most important factor in determining my health and ability to function.**

[] Strongly Disagree [] Disagree [] Agree [] Strongly Agree [] Does Not Apply

**3. I am confident that I can take actions that will help prevent or minimize some symptoms and problems associated with my health.**

[] Strongly Disagree [] Disagree [] Agree [] Strongly Agree [] Does Not Apply

**4. I know what each of my prescribed medications does.**

[] Strongly Disagree [] Disagree [] Agree [] Strongly Agree [] Does Not Apply

**5. I am confident that I can tell when I need to go get medical care and when I can handle a health problem myself.**

[] Strongly Disagree [] Disagree [] Agree [] Strongly Agree [] Does Not Apply

**6. I am confident I can tell a doctor concerns I have even when he or she does not ask.**

[] Strongly Disagree [] Disagree [] Agree [] Strongly Agree [] Does Not Apply

**7. I am confident that I can follow through on medical treatments I need to do at home.**

[] Strongly Disagree [] Disagree [] Agree [] Strongly Agree [] Does Not Apply

**8. I understand the nature and causes of my health problems.**

[] Strongly Disagree [] Disagree [] Agree [] Strongly Agree [] Does Not Apply

**9. I know the different medical treatment options available for my health condition.**

[] Strongly Disagree [] Disagree [] Agree [] Strongly Agree [] Does Not Apply

**10. I have been able to maintain the lifestyle changes for my health that I have made.**

[] Strongly Disagree [] Disagree [] Agree [] Strongly Agree [] Does Not Apply

**11. I know how to prevent further problems with my health.**

[] Strongly Disagree [] Disagree [] Agree [] Strongly Agree [] Does Not Apply

**12. I am confident I can figure out solutions when new situations or problems arise with my health.**

[] Strongly Disagree [] Disagree [] Agree [] Strongly Agree [] Does Not Apply

**13. I am confident I can maintain lifestyle changes, like diet and exercise, even during times of stress.**

[] Strongly Disagree [] Disagree [] Agree [] Strongly Agree [] Does Not Apply”

**4. GENETIC TESTING**

The following questions ask about genetic testing in you and your family members.

You received a genetic test result related to cholesterol and heart disease as a part of this study. Thinking about that result, have you shared your results with any of the following family members? You may indicate *Yes, No,* or *I don't currently have this family member*.

| | Don't currently have this family member(s) | | | If yes, number of family members you have shared with | If yes, has this person had genetic testing for cholesterol or heart disease risk him/herself? | | |
|---|---|---|---|---|---|---|---|
| | | Yes | No | | Yes | No | I don't know |
| a. Your mother | ☐ | ☐ | ☐ | NA | ☐ | ☐ | ☐ |
| b. Your father | ☐ | ☐ | ☐ | NA | ☐ | ☐ | ☐ |
| c. Your sisters | ☐ | ☐ | ☐ | __ | ☐ | ☐ | ☐ |
| d. Your brothers | ☐ | ☐ | ☐ | __ | ☐ | ☐ | ☐ |
| e. Your children | ☐ | ☐ | ☐ | __ | ☐ | ☐ | ☐ |
| f. Your grandchildren | ☐ | ☐ | ☐ | __ | ☐ | ☐ | ☐ |

#### 5. PREFERENCES FOR RECEIVING GENETIC TEST RESULTS

In this study, the genetic counselor on the research team contacted you first about a genetic result identified in your Million Veteran Program blood sample. He/she then helped you get clinical genetic testing and then shared those results with you and your primary care provider. The following questions ask about your preferences for receiving results like these.

**1. Who should the research team have contacted first about your genetic research results?**

[] You
[] Your primary care provider
[] Unsure / I don't know
[] Other:_______________________________________________

**2. After you got the clinical genetic testing to confirm the research result, who should have told you those results?**

[] The genetic counselor on the research team
[] Your primary care provider
[] Unsure / I don't know
[] Other:_______________________________________________

#### 6. FEELINGS ABOUT GENETIC TESTING

The following questions ask about how you felt after receiving your genetic test results. Please indicate how much you had each specific feeling in the past week for each question. The response options for each question are: *not at all, a little, somewhat, a good deal, or a great deal.*

**1. How upset did you feel about your genetic test result?**

[] Not at all [] A little [] Somewhat [] A good deal [] A great deal

**2. How anxious or nervous did you feel about your genetic test result?**

[] Not at all [] A little [] Somewhat [] A good deal [] A great deal

**3. How sad did you feel about your genetic test result?**

[] Not at all [] A little [] Somewhat [] A good deal [] A great deal

4. **How happy did you feel about your genetic test result?**

   [] Not at all [] A little [] Somewhat [] A good deal [] A great deal

5. **How relieved did you feel about your genetic test result?**

   [] Not at all [] A little [] Somewhat [] A good deal [] A great deal

6. **How much did you feel that you understood clearly your choices for disease prevention or early detection?**

   [] Not at all [] A little [] Somewhat [] A good deal [] A great deal

7. **How helpful was the information you received from your genetic test result in planning for the future?**

   [] Not at all [] A little [] Somewhat [] A good deal [] A great deal

8. **How frustrated did you feel that there are no definite disease prevention guidelines for you?**

   [] Not at all [] A little [] Somewhat [] A good deal [] A great deal

9. **How uncertain did you feel about what your genetic test result means for you?**

   [] Not at all [] A little [] Somewhat [] A good deal [] A great deal

10. **How uncertain did you feel about what your genetic test result means for your child(ren) and/or family's risk of disease?**

    [] Not at all [] A little [] Somewhat [] A good deal [] A great deal

11. **How concerned did you feel that your genetic test result would affect your health insurance status?**

    [] Not at all [] A little [] Somewhat [] A good deal [] A great deal

12. **How concerned did you feel that your genetic test result would affect your employment status?**

    [] Not at all [] A little [] Somewhat [] A good deal [] A great deal

#### 7. HEALTH CARE AND HEALTHCARE UTILIZATION

The following statements are about the medical care you've received since enrolling in this study. The statements include some medical tests and procedures. Please indicate if you've had any of these tests or procedures in the last 6 months. You may respond *Yes, No,* or *I'm not sure/I don't know*."

| | Yes | No | I'm not sure/I don't know | If yes, how many of these did you have in the last 6 months? | Notes |
|---|---|---|---|---|---|
| a. Blood pressure check | ☐ | ☐ | ☐ | | |
| b. Cholesterol test | ☐ | ☐ | ☐ | | |
| c. Another lab test for heart health (example C-reactive protein, CK, troponin) | ☐ | ☐ | ☐ | | |
| d. Electrocardiogram (EKG or ECG) | ☐ | ☐ | ☐ | | |
| e. Echocardiogram (Echo or heart ultrasound) | ☐ | ☐ | ☐ | | |
| f. Stress test | ☐ | ☐ | ☐ | | |
| g. Another imaging test of your heart (cardiac calcium score test, coronary angiogram, CT scan of your heart or chest) | ☐ | ☐ | ☐ | | |
| h. Any cardiac procedure, such as coronary stenting, coronary angiography, or coronary bypass surgery | ☐ | ☐ | ☐ | | |

The following statements include some types of healthcare providers. Please indicate if you've seen any of the following providers in the last 6 months. You may respond *Yes, No,* or *I'm not sure/I don't know*."

| | Yes | No | I'm not sure/I don't know | If yes, how many times did you see this kind of provider in the last 6 months? | Notes |
|---|---|---|---|---|---|
| a. VA primary care provider | ☐ | ☐ | ☐ | | |
| b. Non-VA primary care provider | ☐ | ☐ | ☐ | | |
| c. Cardiologist | ☐ | ☐ | ☐ | | |
| d. Geneticist or genetic counselor other than the research genetic counselor | ☐ | ☐ | ☐ | | |
| e. Any other cholesterol specialist | ☐ | ☐ | ☐ | | |
| f. Dietician or nutritionist | ☐ | ☐ | ☐ | | |

"In the last 6 months, have you been admitted to the hospital?
☐ No
☐ Yes

[If Yes] How many times have you been admitted in the last 6 months?
☐ 1 [answer A]
☐ 2 [answer A & B]
☐ 3 or more [answer A, B, & C]

A. Admission 1
Please provide us with the reason for this hospital admission:
How many days did you spend in the hospital?
Did you spend any time in the ICU?
☐ No
☐ Yes
If yes how may days?______

B. Admission 2
Please provide us with the reason for this hospital admission:
How many days did you spend in the hospital?
Did you spend any time in the ICU?
☐ No
☐ Yes
If yes how may days?______

C. Admission 3
Please provide us with the reason for this hospital admission:
How many days did you spend in the hospital?
Did you spend any time in the ICU?
☐ No
☐ Yes
If yes how may days?______

**8. BELIEFS ABOUT MEDICINES**

The following statements are related to your thoughts about medications in general. For each of the following statements, please indicate whether you *Strongly Disagree*, *Disagree*, *Neither Agree Nor Disagree*, *Agree*, or *Strongly Agree*.

1. **Doctors use too many medicines.**

   [] Strongly Disagree [] Disagree [] Neither Agree Nor Disagree []Agree [] Strongly Agree

2. **People who take medicines should stop their treatment every now and then.**

   [] Strongly Disagree [] Disagree [] Neither Agree Nor Disagree []Agree [] Strongly Agree

3. **If doctors have more time with patients they would prescribe fewer medicines.**

   [] Strongly Disagree [] Disagree [] Neither Agree Nor Disagree []Agree [] Strongly Agree

4. **Doctors place too much trust in medicines.**

   [] Strongly Disagree [] Disagree [] Neither Agree Nor Disagree []Agree [] Strongly Agree

5. **Medicines do more harm than good.**

   [] Strongly Disagree [] Disagree [] Neither Agree Nor Disagree []Agree [] Strongly Agree

6. **Most medicines are addictive.**

   [] Strongly Disagree [] Disagree [] Neither Agree Nor Disagree []Agree [] Strongly Agree

7. **All medicines are poisons.**

   [] Strongly Disagree [] Disagree [] Neither Agree Nor Disagree []Agree [] Strongly Agree

8. **Natural remedies are safer than medicines.**

   [] Strongly Disagree [] Disagree [] Neither Agree Nor Disagree []Agree [] Strongly Agree”
