## Supplementary File 4 for "Introducing return of results in the Million Veteran Program: Design and pilot results of the MVP-ROAR Familial Hypercholesterolemia Study"

VA | U.S. Department of Veterans Affairs

MVP ROAR Return Of Actionable Results

**Million Veteran Program**
Washington, DC 20420

### MVP-ROAR BASELINE SURVEY

**PARTICIPANT ID:** ______________________

The following questions are intended to collect information about you and your health care. You may choose to skip any question that you do not wish to answer.

#### 1. SELF-RELATED HEALTH AND QUALITY OF LIFE

This information will help keep track of how you feel and how well you are able to do your usual activities. If you are unsure how to answer a question, please give the best answer you can.

**9. <u>Compared to one year ago</u>, how would you rate your emotional problems (such as feeling anxious, depressed, or irritable) now?**

[] Much better
[x] Slightly better
[] About the same
[] Slightly worse
[] Much worse"

**2. PATIENT ACTIVATION**

The following are statements that people sometimes make when they talk about their health. Please indicate how much you disagree or agree with each statement as it applies to you personally. Your answers should be what is true for you and not just what you think others expect of you. Your choices are *Strongly Disagree, Disagree, Agree,* or *Strongly Agree*. If the statement does not apply to you, please say *Does Not Apply*.

**1. When all is said and done, I am the person who is responsible for managing my health.**

[] Strongly Disagree [] Disagree [x] Agree [] Strongly Agree [] Does Not Apply

**2. Taking an active role in my own health care is the most important factor in determining my health and ability to function.**

[] Strongly Disagree [] Disagree [] Agree [x] Strongly Agree [] Does Not Apply

**3. I am confident that I can take actions that will help prevent or minimize some symptoms and problems associated with my health.**

[] Strongly Disagree [] Disagree [] Agree [x] Strongly Agree [] Does Not Apply

**4. I know what each of my prescribed medications does.**

[] Strongly Disagree [] Disagree [] Agree [x] Strongly Agree [] Does Not Apply

**5. I am confident that I can tell when I need to go get medical care and when I can handle a health problem myself.**

[] Strongly Disagree [] Disagree [] Agree [x] Strongly Agree [] Does Not Apply

**6. I am confident I can tell a doctor concerns I have even when he or she does not ask.**

[] Strongly Disagree [] Disagree [] Agree [x] Strongly Agree [] Does Not Apply

**7. I am confident that I can follow through on medical treatments I need to do at home.**

[] Strongly Disagree [] Disagree [x] Agree [x] Strongly Agree [] Does Not Apply

**8. I understand the nature and causes of my health problems.**

[] Strongly Disagree [] Disagree [x] Agree [] Strongly Agree [] Does Not Apply

**9. I know the different medical treatment options available for my health condition.**

[] Strongly Disagree [] Disagree [] Agree []x Strongly Agree [] Does Not Apply

**10. I have been able to maintain the lifestyle changes for my health that I have made.**

[] Strongly Disagree [] Disagree [] Agree []x Strongly Agree [] Does Not Apply

**11. I know how to prevent further problems with my health.**

[] Strongly Disagree [] Disagree x[] Agree [] Strongly Agree [] Does Not Apply

**12. I am confident I can figure out solutions when new situations or problems arise with my health.**

[] Strongly Disagree [] Disagree [] Agree [x] Strongly Agree [] Does Not Apply

**13. I am confident I can maintain lifestyle changes, like diet and exercise, even during times of stress.**

[] Strongly Disagree [] Disagree [] Agree []x Strongly Agree [] Does Not Apply”

**3. BELIEFS ABOUT MEDICINES**

The following statements are related to your thoughts about medications in general. For each of the following statements, please indicate whether you *Strongly Disagree, Disagree, Neither Agree Nor Disagree, Agree*, or *Strongly Agree*.

1. **Doctors use too many medicines.**

[] Strongly Disagree [] Disagree [x] Neither Agree Nor Disagree []Agree [] Strongly Agree

2. **People who take medicines should stop their treatment every now and then.**

[] Strongly Disagree [] Disagree [x] Neither Agree Nor Disagree []Agree [] Strongly Agree

3. **If doctors have more time with patients they would prescribe fewer medicines.**

[] Strongly Disagree [] Disagree x[] Neither Agree Nor Disagree []Agree [] Strongly Agree

4. **Doctors place too much trust in medicines.**

[] Strongly Disagree [] Disagree [x] Neither Agree Nor Disagree x[]Agree [] Strongly Agree

5. **Medicines do more harm than good.**

[] Strongly Disagree [x] Disagree [] Neither Agree Nor Disagree []Agree [] Strongly Agree

6. **Most medicines are addictive.**

[x] Strongly Disagree [] Disagree [] Neither Agree Nor Disagree []Agree [] Strongly Agree

7. **All medicines are poisons.**

[x] Strongly Disagree [] Disagree [] Neither Agree Nor Disagree []Agree [] Strongly Agree

8. **Natural remedies are safer than medicines.**

[x] Strongly Disagree [] Disagree [] Neither Agree Nor Disagree []Agree [] Strongly Agree

#### 4. DEMOGRAPHIC INFORMATION

1. **What is your race? (*Please mark all that apply*)**

[] White
[x] Black / African-American
[] American Indian / Alaska Native
[] Chinese
[] Japanese
[] Asian Indian
[] Other Asian
[] Filipino
[] Pacific Islander
[] Other

2. **Are you Spanish, Hispanic, or Latino?**

[x] No, not Spanish, Hispanic, or Latino
[] Yes, Mexican, Mexican American, Chicano
[] Yes, Puerto Rican
[] Yes, Cuban
[] Yes, other Spanish, Hispanic, Latino
