## Supplementary File 5 for "Introducing return of results in the Million Veteran Program: Design and pilot results of the MVP-ROAR Familial Hypercholesterolemia Study"

**Million Veteran Program**
Washington, DC 20420

September 1, 2023

Dear Dr. Mickey Mouse,

We are writing to let you know that one of your patients, John Doe (DOB 01/01/1900), was recently identified to have a genetic variant associated with **familial hypercholesterolemia (FH)**. This variant was first found through his participation in the Million Veteran Program (MVP) research study and was then confirmed in a clinical laboratory as a part of the MVP-ROAR (Return of Actionable Results) study. A copy of the clinical variant report is enclosed. The results of a fasting lipid profile are also enclosed.

Your patient has received genetic counseling about this result from Morgan Danowski, MS, CGC, but he will probably have questions for you, too.

Patients will FH have a markedly elevated risk of atherosclerotic cardiovascular disease and require aggressive lowering of their LDL cholesterol. We have enclosed a treatment algorithm you might find helpful, based on the 2018 multisociety Guideline on the Management of Blood Cholesterol (Grundy SM, *et al. Circulation* 2018). You might also consider referring this patient to a preventive cardiologist, lipidologist, or other specialist with FH expertise. Because FH is inherited in an autosomal dominant manner, guidelines recommend screening of first-degree relatives (parents, siblings, children) for FH. The FH Foundation website (https://thefhfoundation.org/) includes many helpful resources for patients and providers about FH, including providers in your area who treat patients with FH.

We also invite you to contact Morgan Danowski, MS, CGC at 617-595-5635 for any additional questions you might have about these results and recommendations for next steps in clinical management. She would be happy to discuss the care of this Veteran with you further.

Sincerely,

Jason L. Vassy, M.D., M.P.H
*MVP-ROAR Principal Investigator*

### Familial hypercholesterolemia clinical management guidelines

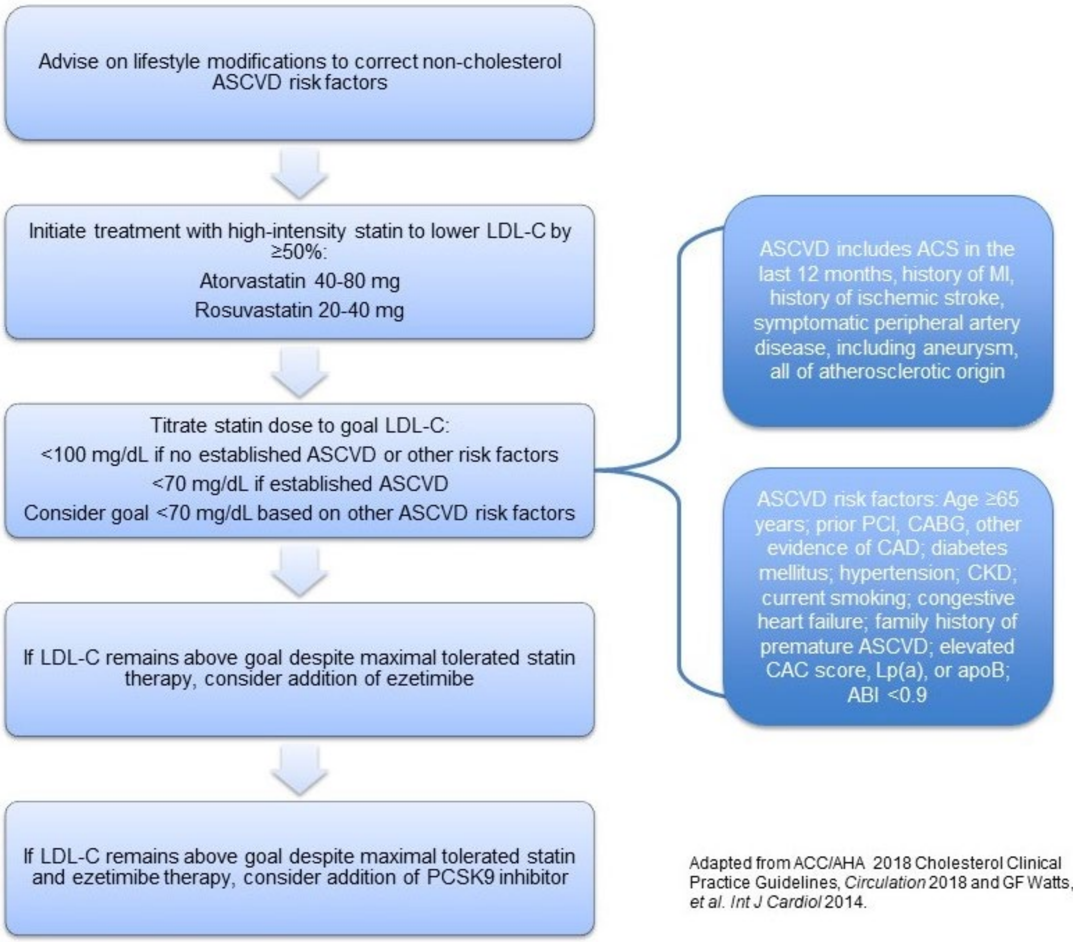


Adapted from ACC/AHA 2018 Cholesterol Clinical Practice Guidelines, *Circulation* 2018 and GF Watts, *et al. Int J Cardiol* 2014.
