## Supplementary File 6 for "Introducing return of results in the Million Veteran Program: Design and pilot results of the MVP-ROAR Familial Hypercholesterolemia Study"

U.S. Department of Veterans Affairs

**Million Veteran Program**
Washington, DC 20420

MVP ROAR Return Of Actionable Results

<DATE>

To the family member of <FIRST LAST NAME>,

You are receiving this letter because <FIRST LAST NAME> has been diagnosed with a genetic condition called **familial hypercholesterolemia (FH)**. The purpose of this letter is to provide you with a summary of what this condition might mean for you and other members of your family.

FH is a genetic disorder that causes high cholesterol levels, resulting in a 20-times greater risk of premature heart disease (including heart attack) if left untreated. A person can have FH without any physical signs or symptoms and may feel and look healthy. However, FH needs to be treated with medications and lifestyle changes (not smoking, regular exercise and a healthy diet). The good news is that these actions can prevent the heart disease due to FH.

**Mr./Ms. <LAST NAME> was found to have an inherited change in the *LDLR* gene, associated with FH. This genetic change is called the <VARIANT NAME> variant**. The test was performed at the Invitae laboratory. The lab accession number for Mr./Ms. <LAST NAME> is <ACCESSION NUMBER>, and a copy of the test result is included with this letter.

While this finding has implications for Mr./Ms. <LAST NAME>, it also has meaning for his/her family members. First-degree relatives (parents, children, siblings) have a 50% chance of also having this same inherited change. **You are receiving this letter because you fall into that category. We recommend that you talk to your healthcare provider and consider genetic counseling to learn more about testing opportunities for yourself.** A blood test is available to see whether you carry the same genetic change as your family member.

To find a genetic counselor, you may visit www.nsgc.org and click on "Find a Genetic Counselor" to search for genetic counselors in your area. You may also contact me, Morgan Danowski, MS, CGC, at 857-364-3182. I am the genetic counselor who spoke to Mr./Ms. <LAST NAME> about his/her results. Invitae has offered to provide genetic testing to first-degree family members of Mr./Ms. <LAST NAME> free of charge. Please contact me if you are interested in pursuing this option. Otherwise, your own healthcare provider can work with you and your insurance company to find an alternative way to get this genetic testing.

I hope that this information has been helpful for you. If you have any questions or concerns, please do not hesitate to contact me at the number above.

Sincerely,

Morgan Danowski, MS, CGC
