## Supplementary File 7 for "Introducing return of results in the Million Veteran Program: Design and pilot results of the MVP-ROAR Familial Hypercholesterolemia Study"

INVITAE

### INVITAE DIAGNOSTIC TESTING RESULTS

| | | | | | |
|---|---|---|---|---|---|
| **Patient name:** | John Doe | **Sample type:** | Blood | **Report date:** | 05/15/2023 |
| **DOB:** | 01/01/1900 | **Sample collection date:** | 05/01/2023 | **Invitae #:** | RQ1234567 |
| **Sex assigned at birth:** | Male | **Sample accession date:** | 05/02/2023 | **Clinical team:** | Morgan Danowski<br>Jason Vassy |
| **Gender:** | | | | | |
| **Patient ID (MRN):** | 1234 | | | | |

**Reason for testing**

Diagnostic test for a personal history of disease

**Test performed**

Sequence analysis and deletion/duplication testing of the 4 genes listed in the Genes Analyzed section.

- Invitae Familial Hypercholesterolemia Panel

#### RESULT: POSITIVE

**One Pathogenic variant identified in LDLR. LDLR is associated with autosomal dominant and recessive hypercholesterolemia.**

| GENE | VARIANT | ZYGOSITY | VARIANT CLASSIFICATION |
|---|---|---|---|
| LDLR | c.1238C>T (p.Thr413Met) | heterozygous | PATHOGENIC |

**About this test**

This diagnostic test evaluates 4 gene(s) for variants (genetic changes) that are associated with genetic disorders. Diagnostic genetic testing, when combined with family history and other medical results, may provide information to clarify individual risk, support a clinical diagnosis, and assist with the development of a personalized treatment and management strategy.

#### Clinical comments

- When a single Variant of Uncertain Significance is found in a requisitioned gene that is only associated with autosomal recessive condition(s), it may not be included in the report.


© 2023 Invitae Corporation

#### Next steps

- This is a medically important result that should be discussed with a healthcare provider, such as a genetic counselor, to learn more about this result and the appropriate next steps for further evaluation, treatment and/or management. This result should be interpreted within the context of additional laboratory results, family history and clinical findings.

- Please see PMID: 21600525, 30071997, 21600528, 30423391, 21600530, 30798614, and 28225426 for management guidelines regarding LDLR-related condition(s).

- Consider sharing this result with relatives as they may also be at risk. Details on our Family Variant Testing program can be found at **www.invitae.com/family**.

- Register your test at **www.invitae.com/patients** to download a digital copy of your results. You can also access educational resources about how your results can help inform your health.


© 2023 Invitae Corporation

#### Clinical summary

A Pathogenic variant, c.1238C>T (p.Thr413Met), was identified in LDLR.

- The LDLR gene is associated with familial hypercholesterolemia (FH) (MedGen UID: 5688). Heterozygous FH (HeFH) results from one pathogenic variant, while homozygous FH (HoFH) is a more severe presentation caused by the presence of homozygous or compound heterozygous pathogenic variants.
- This result is consistent with a predisposition to, or diagnosis of, autosomal dominant familial hypercholesterolemia.
- FH is characterized by an increase of low-density lipoprotein cholesterol (LDL-C) that can cause atherosclerotic plaque deposits and lead to premature coronary artery disease. Untreated individuals have a significantly increased risk of developing coronary heart disease (CHD) (PMID: 21600525, 15177124). Untreated individuals who are homozygous or compound heterozygous for two pathogenic variants (HoFH) typically have marked hypercholesterolemia and are at risk of developing severe CHD, often in childhood or adolescence (PMID: 21986285). For more information about diagnosis and management of FH, please visit Invitae's website at www.invitae.com/management-guidelines.
- Biological relatives have a chance of being at risk for autosomal dominant familial hypercholesterolemia and have a chance of being carriers for autosomal recessive familial hypercholesterolemia. Those at risk should consider testing.

#### Variant details

LDLR, Exon 9, c.1238C>T (p.Thr413Met), heterozygous, PATHOGENIC

- This sequence change replaces threonine, which is neutral and polar, with methionine, which is neutral and non-polar, at codon 413 of the LDLR protein (p.Thr413Met).
- This variant is present in population databases (rs368562025, gnomAD 0.01%), including at least one homozygous and/or hemizygous individual.
- This missense change has been observed in individual(s) with familial hypercholesterolemia (PMID: 11857755, 20236128, 25682026; Invitae). In at least one individual the data is consistent with being in trans (on the opposite chromosome) from a pathogenic variant.
- ClinVar contains an entry for this variant (Variation ID: 161276).
- Advanced modeling of protein sequence and biophysical properties (such as structural, functional, and spatial information, amino acid conservation, physicochemical variation, residue mobility, and thermodynamic stability) performed at Invitae indicates that this missense variant is expected to disrupt LDLR protein function.
- For these reasons, this variant has been classified as Pathogenic.


© 2023 Invitae Corporation

#### Genes analyzed

This table represents a complete list of genes analyzed for this individual, including the relevant gene transcript(s). If more than one transcript is listed for a single gene, variants were reported using the first transcript listed unless otherwise indicated in the report. An asterisk (*) indicates that this gene has a limitation. Please see the Limitations section for details. Results are negative unless otherwise indicated in the report. Benign and Likely Benign variants are not included in this report and in specific scenarios variants of uncertain significance in the requisitioned gene(s) may not be included in this report. These variants are available upon request.

| GENE | TRANSCRIPT |
|---|---|
| APOB | NM_000384.2 |
| LDLR | NM_000527.4 |

| GENE | TRANSCRIPT |
|---|---|
| LDLRAP1 | NM_015627.2 |
| PCSK9 | NM_174936.3 |


© 2023 Invitae Corporation

#### Methods

- Genomic DNA obtained from the submitted sample is enriched for targeted regions using a hybridization-based protocol, and sequenced using Illumina technology. Unless otherwise indicated, all targeted regions are sequenced with ≥50x depth or are supplemented with additional analysis. Reads are aligned to a reference sequence (GRCh37), and sequence changes are identified and interpreted in the context of a single clinically relevant transcript, indicated in the Genes Analyzed table. Enrichment and analysis focus on the coding sequence of the indicated transcripts, 20bp of flanking intronic sequence, and other specific genomic regions demonstrated to be causative of disease at the time of assay design. Promoters, untranslated regions, and other non-coding regions are not otherwise interrogated. For some genes only targeted loci are analyzed. Exonic deletions and duplications are called using an in-house algorithm that determines copy number at each target by comparing the read depth for each target in the proband sequence with both mean read-depth and read-depth distribution, obtained from a set of clinical samples. Markers across the X and Y chromosomes are analyzed for quality control purposes and may detect deviations from the expected sex chromosome complement. Such deviations may be included in the report in accordance with internal guidelines. Variants are reported according to the Human Genome Variation Society (HGVS) guidelines. Confirmation of the presence and location of reportable variants is performed as needed based on stringent criteria using one of several validated orthogonal approaches (PubMed ID 30610921). Sequencing is performed by Invitae Corporation (1400 16th Street, San Francisco, CA 94103, #05D2040778). Confirmatory sequencing is performed by Invitae Corporation (1400 16th Street, San Francisco, CA 94103, #05D2040778). RNA sequencing is performed by Invitae Corporation (1400 16th Street, San Francisco, CA 94103, #05D2040778).

  The following additional analyses are performed if relevant to the requisition. For PMS2 exons 12-15, the reference genome has been modified to force all sequence reads derived from PMS2 and the PMS2CL pseudogene to align to PMS2, and variant calling algorithms are modified to support an expectation of 4 alleles. If a rare SNP or indel variant is identified by this method, both PMS2 and the PMS2CL pseudogene are amplified by long-range PCR and the location of the variant is determined by Pacific Biosciences (PacBio) SMRT sequencing of the relevant exon in both long-range amplicons. If a CNV is identified, MLPA or MLPA-seq is run to confirm the variant. If confirmed, both PMS2 and PMS2CL are amplified by long-range PCR, and the identity of the fixed differences between PMS2 and PMS2CL are sequenced by PacBio from the long-range amplicon to disambiguate the location of the CNV. For C9orf72 repeat expansion testing, hexanucleotide repeat units are detected by repeat-primed PCR (RP-PCR) with fluorescently labeled primers followed by capillary electrophoresis. Interpretation Reference Ranges: Benign (Normal Range): <25 repeat units, Uncertain: 25-30 repeat units, Pathogenic (Full Mutation): >=31 repeat units (PMID: 21944779, 22406228, 23111906, 28689190, 31315673, 33168078, 33575483). A second round of RP-PCR utilizing a non-overlapping set of primers is used to confirm the initial call in the case of suspected allele sizes of 22 or more repeats. For RNA analysis of the genes indicated in the Genes Analyzed table, complementary DNA is synthesized by reverse transcription from RNA derived from a blood specimen and enriched for specific gene sequences using capture hybridization. After high-throughput sequencing using Illumina technology, the output reads are aligned to a reference sequence (genome build GRCh37; custom derivative of the RefSeq transcriptome) to identify the locations of exon junctions through the detection of split reads. The relative usage of exon junctions in a test specimen is assessed quantitatively and compared to the usage seen in control specimens. Abnormal exon junction usage is evaluated as evidence in the Sherloc variant interpretation framework. If an abnormal splicing pattern is predicted based on a DNA variant outside the typical reportable range, as described above, the presence of the variant is confirmed by targeted DNA sequencing.
- A PMID is a unique identifier referring to a published, scientific paper. Search by PMID at **http://www.ncbi.nlm.nih.gov/pubmed**.
- An rsID is a unique identifier referring to a single genomic position, and is used to associate population frequency information with sequence changes at that position. Reported population frequencies are derived from a number of public sites that aggregate data from large-scale population sequencing projects, including ExAC (**http://exac.broadinstitute.org**), gnomAD (**http://gnomad.broadinstitute.org**), and dbSNP (**http://ncbi.nlm.nih.gov/SNP**).
- A MedGen ID is a unique identifier referring to an article in MedGen, NCBI's centralized database of information about genetic disorders and phenotypes. Search by MedGen ID at **http://www.ncbi.nlm.nih.gov/medgen**. An OMIM number is a unique identifier referring to a comprehensive entry in Online Mendelian Inheritance in Man (OMIM). Search by OMIM number at **http://omim.org/**.
- Invitae uses information from individuals undergoing testing to inform variant interpretation. If "Invitae" is cited as a reference in the variant details this may refer to the individual in this requisition and/or historical internal observations.


© 2023 Invitae Corporation

#### Limitations

Based on validation study results, this assay achieves >99% analytical sensitivity and specificity for single nucleotide variants, insertions and deletions <15bp in length, and exon-level deletions and duplications. Invitae's methods also detect insertions and deletions larger than 15bp but smaller than a full exon but sensitivity for these may be marginally reduced. Invitae's deletion/duplication analysis determines copy number at a single exon resolution at virtually all targeted exons. However, in rare situations, single-exon copy number events may not be analyzed due to inherent sequence properties or isolated reduction in data quality. Certain types of variants, such as structural rearrangements (e.g. inversions, gene conversion events, translocations, etc.) or variants embedded in sequence with complex architecture (e.g. short tandem repeats or segmental duplications), may not be detected. Additionally, it may not be possible to fully resolve certain details about variants, such as mosaicism, phasing, or mapping ambiguity. Unless explicitly guaranteed, sequence changes in the promoter, non-coding exons, and other non-coding regions are not covered by this assay. Please consult the test definition on our website for details regarding regions or types of variants that are covered or excluded for this test. This report reflects the analysis of an extracted genomic DNA sample. While this test is intended to reflect the analysis of extracted genomic DNA from a referred patient, in very rare cases the analyzed DNA may not represent that individual's constitutional genome, such as in the case of a circulating hematolymphoid neoplasm, bone marrow transplant, blood transfusion, chimerism, culture artifact or maternal cell contamination. Interpretations are made on the assumption that any clinical information provided, including specimen identity, is accurate. Invitae's RNA analysis is not designed for use as a stand-alone diagnostic method and cannot determine absolute RNA levels. Results from the RNA analysis may not be informative for interpreting copy number gains.

#### Disclaimer

DNA studies do not constitute a definitive test for the selected condition(s) in all individuals. It should be realized that there are possible sources of error. Errors can result from trace contamination, rare technical errors, rare genetic variants that interfere with analysis, recent scientific developments, and alternative classification systems. This test should be one of many aspects used by the healthcare provider to help with a diagnosis and treatment plan, but it is not a diagnosis itself. This test was developed and its performance characteristics determined by Invitae. It has not been cleared or approved by the FDA. The laboratory is regulated under the Clinical Laboratory Improvement Act (CLIA) as qualified to perform high-complexity clinical tests (CLIA ID: 05D2040778). This test is used for clinical purposes. It should not be regarded as investigational or for research.

#### This report has been reviewed and approved by:

Jennifer Sanmann, Ph.D., FACMG
Clinical Molecular Geneticist & Clinical Cytogeneticist


© 2023 Invitae Corporation

This document is not part of Invitae's clinical report and does not represent medical advice. These are general guidelines that are not specific to your result and may not represent all relevant international recommendations. You can use this guide to talk to your healthcare provider about your test results, clinical history, and the most current guidelines. This guide may not be appropriate for results that are suspected to be blood-limited, possibly mosaic, or suggestive of a larger imbalance of genetic material. Invitae recognizes that individuals have diverse gender and sexual identities. In this guide, the terms female, male, women, and men refer to sex assigned at birth.

#### What is a positive LDLR result?

A positive test result means that a genetic change (variant) was found in the LDLR gene. A LDLR variant is considered "pathogenic" or "likely pathogenic" because it is associated with familial hypercholesterolemia (FH).

#### What does this mean?

Familial hypercholesterolemia (FH) is characterized by elevated LDL cholesterol and increased risk of early onset coronary heart disease (CHD). Some individuals will have yellow patches under the skin (xanthomas) or around the eyes (xanthelasmas) caused by the build up of cholesterol. Another physical finding in some individuals with FH is cholesterol deposits around the cornea of the eye (corneal arcus). However, many individuals with FH will have no obvious physical findings despite having elevated cholesterol that predisposes to high risk of CHD. CHD causes the buildup of plaque within the blood vessels, potentially leading to heart attacks and/or strokes. However, symptoms, severity, and age of onset can vary. Treatment of FH may also affect onset of symptoms and disease progression. Individuals may have different conditions or symptoms depending on whether they inherit one or two variants in LDLR. Some people inherit two LDLR variants, which may cause homozygous FH. See the table later in this guide for more information and possible next steps.

#### What does this mean for family members?

Relatives should be informed about these results. It is recommended that family members talk with their own healthcare provider about a plan for genetic testing and/or health screening. Genetic testing is a personal choice and some individuals may choose not to have genetic testing. Laws protecting employment and health insurance may apply to individuals undergoing genetic testing (for example, the Genetic Information Nondiscrimination Act in the United States).

**Will family members have the same variant(s)?**
The image shows where a LDLR variant may have come from. Any individual can inherit and pass on a LDLR variant, regardless of sex.

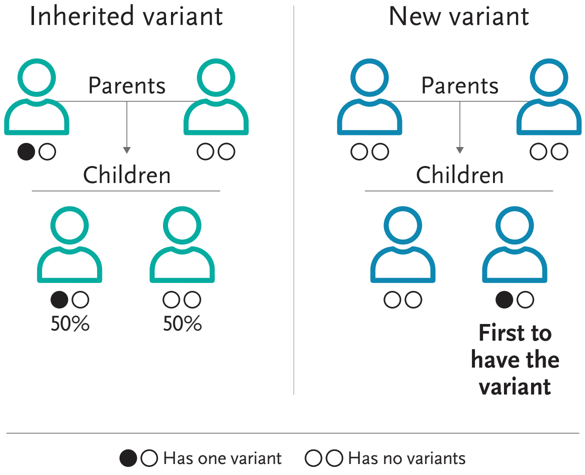


LDLR variants can be inherited from a parent or an individual may be the first person in the family to have a new LDLR variant. Siblings and other relatives may also have this LDLR variant. Individuals with a LDLR variant can pass it on to children.

For individuals who are planning a family, reproductive options may be available to help lower the chance of passing on a variant to children.

#### Create a plan with a healthcare provider


© 2020 Invitae Corporation. All Rights Reserved. 06/06/2023

These options are a guide for an individual and their healthcare provider. They are meant to be used along with an individual's genetic test results and other health information as part of a discussion to make a personalized care plan. Each option may or may not be right for an individual. A positive test result on its own cannot predict how a condition may affect an individual. This guide may not be appropriate for results that are suspected to be blood-limited, possibly mosaic, or suggestive of a larger imbalance of genetic material.

**Options to consider**

| TOPIC | OPTION | MORE INFORMATION |
|---|---|---|
| FH | • Evaluate for signs and symptoms of FH. | • A clinical diagnosis of FH can be made based on multiple tools including Dutch Lipid Clinic Network (DLCN), Make Early Diagnosis Prevent Early Death (MEDPED), or Simon-Broome Registry. (1)<br>◦ The finding of a positive LDLR variant is considered the gold standard in establishing a diagnosis of FH. (2)<br>• Additional evaluations following a diagnosis of FH may include cholesterol screening, physical examination, and/or cardiovascular tests that may assess for coronary heart disease (CHD). (1,2) |
| | • Management of FH may involve establishing a healthcare team, such as primary care providers, dietitians, nutritionists, cardiologists, endocrinologists, and others. (1,3)<br>• For some individuals with FH, referral to a lipid specialist may be appropriate. (1) | • Screening, diagnostic, and management consensus guidelines are available for FH. (1,3,4,5)<br>• Recommendations for dietary changes (Therapeutic Lifestyle Changes diet) following a diagnosis of FH are available. (1,3)<br>• It is recommended that, at minimum, the following populations with FH be referred to a lipid specialist: (1)<br>◦ Individuals with past difficulties with medical management or control of cholesterol.<br>◦ Individuals who are candidates for more intensive cholesterol-lowering therapy.<br>◦ Individuals who have a family history of early onset CHD.<br>◦ Individuals under age 18. |
| | • For individuals with FH and elevated cholesterol levels, discuss treatment options. | • Treatment for FH involves lowering cholesterol levels. First line treatment for FH is typically with statins (1).<br>◦ Additional treatment options such as ezetimibe, PCSK9 inhibitors, LDL apheresis, or others may be considered depending on clinical history, age, and other factors. (3,4,6)<br>• Early treatment for high cholesterol is highly beneficial. Long-term drug therapy for individuals with FH can substantially reduce or remove the excess lifetime risk of CHD. (1)<br>◦ Individuals with FH and LDL cholesterol greater than or equal to 190 mg/dL after lifestyle changes should be treated with drug therapy. (1) |
| | • Avoid and/or treat risk factors for CHD. (1,6) | • Recommendations for lifestyle changes to reduce risk of CHD include: (1,3)<br>◦ Cessation of smoking.<br>◦ Regular physical activity, healthy diet, and weight control.<br>◦ Blood pressure control.<br>◦ Limited alcohol consumption. |
| LDLR-related research | • Consider options for clinical trials and/or research. | • There are many factors when considering participation in research studies, including time, location, and whether or |


© 2020 Invitae Corporation. All Rights Reserved. 06/06/2023

| TOPIC | OPTION | MORE INFORMATION |
|---|---|---|
| | • To find up-to-date information about available clinical trials, visit ClinicalTrials.gov | not an individual meets the specific requirements to be in a study. |
| Family planning | • Discuss reproductive risks. (7)<br>• Individuals with a LDLR variant have a 50% chance to pass on the variant to a child. | • Preconception and prenatal reproductive options are available and could be discussed in more detail with a reproductive specialist. |
| | • Individuals with a LDLR variant may also have an increased chance to have a child with homozygous FH, if their reproductive partner also has a positive LDLR (or other FH-causing gene) variant. | • Homozygous FH is characterized by extremely high cholesterol (LDL often over 400mg/dL) and, without treatment, very early onset of CHD. |
| | • An individual's reproductive partner can consider genetic testing to help determine the risk of a child inheriting two positive variants and having homozygous FH. (7) | • If an individual's reproductive partner also has a positive LDLR (or other FH-causing gene) variant, there would be a 25% chance to have a child with homozygous FH. |

These options include recommendations from PMID: 21600525 (1), PMID: 30071997 (2), PMID: 21600528 (3), PMID: 30423391 (4), PMID: 21600530 (5), PMID: 30798614 (6), and PMID: 28225426 (7). More information about genetics and disease continues to be available, so please always refer to the current guidelines and recommendations when considering surveillance and treatment options. Information on this document may not include all relevant international recommendations and acts as a supplement to the Invitae result report. This information is not meant to replace a discussion with an individual's healthcare provider and should not be considered or interpreted as medical advice. Additional resources provided within this document do not indicate or imply any endorsement by Invitae with respect to any third party or any website or the products or services offered by any third party.

#### Resources

Genetic counseling can help individuals understand their genetic test results and options for next steps. Reviewing test results with a genetic counselor or other healthcare provider is recommended. Local or telehealth genetic counselors can be identified using the Find a Genetic Counselor search tool at nsgc.org (US and Canada). Individuals who had genetic testing through Invitae can also log in to their patient portal (invitae.com) to view their results, contact a genetic counselor, or join Invitae's Patient Insights Network (PIN), an online platform where individuals can share information about their health and experiences to help advance research and drug development.

**Connect with advocacy groups and other resources**

- Family Heart Foundation: www.familyheart.org

#### Notes for personalized assessment



© 2020 Invitae Corporation. All Rights Reserved. 06/06/2023
