## Supplementary File 8 for "Introducing return of results in the Million Veteran Program: Design and pilot results of the MVP-ROAR Familial Hypercholesterolemia Study"

**Million Veteran Program**
Washington, DC 20420

FULL NAME
ADDRESS LINE 1
ADDRESS LINE 2
CITY, STATE ZIP

DATE

Dear FULL NAME,

This letter is in follow-up to our phone conversation about your genetic test results. As we discussed, you tested **NEGATIVE** for genetic changes associated with **familial hypercholesterolemia (FH)**, a genetic disorder characterized by high cholesterol levels and a 20-times greater risk of premature heart disease.

A negative result means we found no changes or mutations in the genes (*LDLR, APOB, LDLRAP1, PCSK9)* we tested for. Although very unlikely, a negative result does not rule out all hereditary components of FH, or include genes not yet discovered or described in the literature.

The analysis was performed at Invitae, and the accession number for your specimen is [ACCESSION NUMBER] to help locate your result in the future. A copy of the result has been included with this letter for your personal medical record.

In addition, a routine fasting cholesterol test was collected. Your LDL level was [XX] mg/dL. A copy of the result has also been included with this letter.

[INSERT SPECIFIC FOLLOW-UP INFORMATION FROM GENETIC COUNSELING SESSION, SUCH AS: "As we discussed, it is important to eat a healthy diet low in saturated or trans fats, exercise regularly, and continue to take medications as prescribed. You mentioned that you are already taking atorvastatin for your cholesterol. As we discussed, your results will be entered into your medical record and shared with your primary care provider, Dr. [NAME], who can continue to manage your clinical care."]

Heart disease and high cholesterol can be caused by changes in your genes, your health habits (diet and exercise), or a combination of both. You tested negative for mutations in genes associated with FH. While your result does not indicate a known familial mutation, your relatives may consider pursuing genetic testing based on their individual medical and family history. If your relatives would like to discuss this result, they are welcome to contact me with any additional questions.

It was a pleasure speaking with you. Please know I am available at 857-364-3182 if you, your family, or your healthcare providers have any questions.

Sincerely,

Morgan Danowski, MS, CGC
