## Supplementary File 9 for "Introducing return of results in the Million Veteran Program: Design and pilot results of the MVP-ROAR Familial Hypercholesterolemia Study"

VA | U.S. Department of Veterans Affairs

**Million Veteran Program**
Washington, DC 20420

MVP ROAR
Return Of Actionable Results

FULL NAME
ADDRESS LINE 1
ADDRESS LINE 2
CITY, STATE ZIP

DATE

Dear FULL NAME,

This letter is in follow-up to our phone conversation about your genetic test results. As we discussed, you tested POSITIVE for a genetic change in the *LDLR* gene, associated with **familial hypercholesterolemia (FH)**.

FH is a genetic disorder that causes high cholesterol levels, resulting in a 20-times greater risk of premature heart disease (including heart attack) if left untreated. A person can have FH without any physical signs or symptoms and may feel and look healthy. However, FH needs to be treated with medications and lifestyle changes (not smoking, regular exercise and a healthy diet). The good news is that these actions can prevent the heart disease due to FH.

You were found to have an inherited change in the *LDLR* gene, associated with FH. This genetic change is called the [VARIANT NAME] variant. The analysis was performed at Invitae, and the accession number for your specimen is [ACCESSION NUMBER] to help locate your result in the future. A copy of the result has also been included with this letter for your personal medical record.

Below is some general information about FH.

**What is familial hypercholesterolemia (FH)?**
FH is an inherited condition caused by mutations in one of three major genes. Your FH is caused by a mutation is the *LDLR* gene. FH is a relatively common condition; it affects 1 in 250 people. In people with FH, the body does not remove cholesterol (specifically low-density lipoprotein or LDL) as effectively as it should. This causes elevated cholesterol at a young age which leads to premature cardiovascular disease. If untreated, men with FH have a 50% chance of having a heart attack by age 50.

**How is FH inherited?**
We each have two copies of every gene; one we inherit from our mother and the other from our father. FH is typically inherited in an autosomal dominant manner. This means the genetic mutation is found in males and females equally and that only one of the two gene copies needs to have a mutation in order for a person to have the condition.

We know you have a genetic change or mutation in the *LDLR* gene. [TEXT ON IMPLICATIONS FOR FAMILY MEMBERS, SUCH AS "That means you had a 50% chance of passing the *LDLR* gene with the genetic change or mutation to each son and a 50% chance you passed on the "normal" gene. Your sisters also have a 50%
risk of having the same genetic mutation you have. As we discussed, the *LDLR* mutation causing FH in your family likely came from your mother who also had a heart attack at a young age."]

**Can my family members be tested?**
Your family members can be tested for the same genetic mutation found in you if they are interested. They can take a copy of your genetic test result to their providers, who may want to refer them to a genetic professional. I have enclosed a few copies of a family letter you can share with family members who might want genetic testing, plus extra copies of your test result.

**What should I do next?**
In general, people with FH should not smoke, eat a healthy diet that is low is saturated or trans fats, exercise regularly, and make sure to take medications as prescribed. [INSERT SPECIFIC FOLLOW-UP INFORMATION FROM GENETIC COUNSELING SESSION, SUCH AS: "You mentioned that you are already taking atorvastatin for your cholesterol. As we discussed, you plan on sharing these results with your primary care provider, Dr. [NAME], to ask whether she has any new recommendations for your health care. We will also send Dr. [NAME] a copy of these results, along with information about the treatment of FH."]

**Is there support for people with FH?**
The FH Foundation (https://thefhfoundation.org/) is a good resource for general information about FH, treatment plans and ongoing research.

It was a pleasure speaking with you. Please know I am available at 857-364-3182 if you, your family, or your healthcare providers have any questions.

Sincerely,

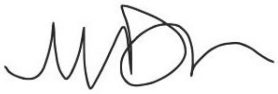

Morgan Danowski, MS, CGC
