## Supplementary File 10 for "Introducing return of results in the Million Veteran Program: Design and pilot results of the MVP-ROAR Familial Hypercholesterolemia Study"

VA | U.S. Department of Veterans Affairs

**Million Veteran Program**
Washington, DC 20420

MVP ROAR
Return Of Actionable Results

### INFORMATION SHEET FOR THE MVP-ROAR STUDY

You are being invited to take part in a research study called MVP-ROAR (Return of Actionable Results), funded by the Department of Veterans Affairs (VA). The person in charge of the study is Dr. Jason Vassy at the VA Boston Healthcare System. Shortly after you receive this information, a staff member from the MVP-ROAR study will call you to review this information, answer your questions, and ask if you are interested in participating in the study.

You recall that you are a participant in the VA Million Veteran Program (MVP) study. Researchers using your MVP blood sample have found a genetic result that might give you, your healthcare providers, and your family members important information about your risk of cardiovascular disease (including heart attack and stroke) due to blood cholesterol. By doing this study, we hope to learn whether confirming these results and reporting them to you and your healthcare providers will help you make better decisions about your health care.

Participation in this research will last about 7 months and will include the following:

- A survey at the beginning of the study, lasting about 30 minutes
- A blood draw or saliva collection at the beginning of the study
- A telephone or video meeting with a genetic counselor to review your genetic results, either at the beginning of the study or at the end of the study (after 6 months)
- A blood draw at the end of the study (after 6 months)
- A survey at the end of the study (after 6 months), lasting about 30 minutes

Before you decide to take part, it is important for you to know why the research is being done and what it will involve. This includes any potential risks and benefits to you. Your participation is completely voluntary. You will not lose any services, benefits, or rights you would normally have if you choose not to volunteer.

**What are key reasons you might choose to volunteer OR NOT TO VOLUNTEER for this study?**

You may want to volunteer to participate in this study if you are interested in learning more about your genetic result, which might give information about you and your family's risk of cardiovascular disease. You may not want to volunteer for this study if you might find information about your risks for certain diseases emotionally distressing.

#### WHY IS THIS STUDY BEING DONE?

Genes are made of DNA that we inherit from each of our parents, making us who we are. Our risks for some diseases are determined by our DNA, by our lifestyle factors (like the food we eat or the things in the environment we are exposed to), or by a combination of DNA and lifestyle factors. In some cases, knowing about your risk for certain diseases might help you and your family take steps to lower your risk for some diseases.

Cardiovascular disease, including stroke, heart attack, and other kinds of heart disease, is common among Veterans. Cardiovascular disease can be caused by your genes, your health habits, or a combination of both. Some common risk factors for cardiovascular disease include smoking, obesity, high cholesterol, high blood pressure, and family history, among other factors. Healthcare providers often look at a patient's cardiovascular disease risk factors and help him or her lower that risk through dietary changes, exercise, quitting smoking, medications, or other steps. Addressing these risk factors can help patients prevent some cases of cardiovascular disease, like heart attacks and strokes.

Researchers using your MVP blood sample have made a genetic discovery that may be important to the risk of cardiovascular disease for you and your family members, related to blood cholesterol. A team of experts has reviewed the findings and decided that genetic results from your sample might help you and your healthcare providers make decisions about your health care. Even if you don't think your cholesterol is particularly high or if you are already receiving treatment for high cholesterol, this genetic result might provide you and your healthcare provider additional information to help take care of your risk of cardiovascular disease. Because genes are shared among blood relatives, this information might also be important for your family members.

However, research findings are not the same as clinical tests done as a part of medical care. For this reason, it would be necessary to repeat this research test in a clinical laboratory to be certain it is accurate. By conducting this research study, we hope to learn if retesting these results and reporting them to you and your healthcare providers helps you make better decisions about your health care and take steps to lower your risk of cardiovascular disease, if appropriate.

#### HOW LONG WILL I BE IN THE STUDY?

This research study is expected to take approximately 3 years. Your individual participation in the study will take approximately 7 months.

#### WHAT WILL HAPPEN IF I TAKE PART IN THE STUDY?

If you agree to participate, you will complete the following steps:

- Over the phone, you will complete a survey lasting approximately 30 minutes. You may skip any questions you prefer not to answer.

- You will go to your local VA laboratory for a fasting blood draw. The lab will draw 2 tubes of blood. One tube will be used to measure your blood cholesterol.
- Some participants may be able to provide a saliva sample instead of a blood draw. The genetic counselor will discuss this option with you over the phone if you choose to participate.
- After your blood or saliva collection, you will be randomly assigned (like the flip of a coin) to receive your genetics results from the MVP-ROAR genetic counselor right away (Immediate Results) or at the end of the study (after 6 months, Delayed Results).

    - If you are assigned to the Immediate Results group, your blood or saliva specimen will be shipped to a clinical genetics lab outside VA to confirm the MVP research result. The lab will send the results to the MVP-ROAR genetic counselor, who will arrange a time to go over your results with you by phone or video. The genetic counselor will explain the results and what importance it might have for your own risk of heart disease and your blood relatives' risk. He/she will also provide additional written information about your risk and what you and your healthcare providers can do to lower that risk. He/she will provide you with a letter you may choose to share with your family members. Your family members may then want to have genetic testing themselves, and the genetic counselor can give them more information about that process. The genetic counselor will also send a copy of your most recent cholesterol and genetic results to your primary care provider(s), along with written materials that might help your provider(s) take care of your heart disease risk. You or your healthcare provider(s) may contact the genetic counselor any time during the study with additional questions or concerns. Your provider may also want to refer you to a heart or cholesterol specialist. The genetic counselor can help your provider find a specialist.

    - If you are assigned to the Delayed Results group, your second tube of blood will be discarded or your saliva specimen placed in temporary storage. You and your primary care provider(s) will receive a letter with your cholesterol results as soon as they are ready, but you will wait 6 months to have your genetic research results confirmed in a clinical laboratory. After you complete your end-of-study survey and blood draw (after 6 months), the genetic counselor will contact you to go over your genetic results by telephone or video, in the same way as for the Immediate Results group. You and your primary care provider(s) will receive the same information the Immediate Results group did at the beginning of the study. Your biospecimen will not be saved after your participation in the study is complete.

- At the end of the study (after 6 months), you will complete a survey lasting about 30 minutes. You may skip any questions you prefer not to answer.
- After this survey, you will go to your local VA laboratory to have either 1 tube of blood drawn for cholesterol testing only (if you are in the Immediate Results group) or 2

tubes of blood drawn for cholesterol testing and clinical genetic testing (if you are in the Delayed Results group).
- A stored saliva sample may be used to obtain genetic test results for participants in the Delayed Results group if a blood draw cannot be arranged.
- During this study, no matter which group you are in, you will continue to receive medical care as usual from your primary care and other providers.
- The decision to use this genetic information in your medical care will be up to you and your healthcare providers. This study does not require you to take a new medication or have any other medical tests or procedures, except the blood draw or saliva collection at the beginning and end of the study

**DO I HAVE TO TAKE PART IN THE STUDY?**

Participation is voluntary. Refusal to take part in the study will involve no penalty or loss of benefits to which you are otherwise entitled. If you are a VA employee, refusal to take part in the study will in no way influence your employment or ratings.

You may discontinue taking part at any time without any penalty or loss of benefits. You will still receive the same medical care you would have received otherwise.

If you withdraw from the study, the research team may continue to review the data already collected for the study but cannot collect further information after you withdraw, except from public records, such as survival data. Specimens already used cannot be withdrawn.

**WHAT IS EXPECTED OF ME IF I TAKE PART IN THIS STUDY?**

As a study participant, you are expected to:
- Keep your study appointments, including telephone or video appointments and lab draws. If you have to miss an appointment, please contact the research staff to reschedule as soon possible.
- Promptly return a saliva kit using a prepaid shipping label, if you are asked to submit a saliva sample
- Complete study surveys as instructed.

**WHAT POSSIBLE RISKS OR DISCOMFORTS MIGHT I HAVE IF I TAKE PART IN THIS STUDY?**

There is always a chance that any procedure can harm you. The procedures in this study are no different. In addition to the risks described below, you may experience a previously unknown risk or side effect.

The blood draws in this study carry the possibility of minor risk and discomfort, including lightheadedness, bleeding, bruising, or infection.

The two surveys in this study may result in emotional distress or discomfort as some questions are related to your personal demographic and medical information.

Learning about genetic risk of a disease in you and your family members might cause you emotional distress.

It is possible that you may feel anxious or distressed about being enrolled into the Delayed Results group as described above.

There is a risk that genetic information obtained as a result of your participation in this study could be used to discriminate against you with regard to your health insurance or your job. There are state, federal, and VA protections that prevent health insurance companies, group health plans, and most employers from discriminating against you based on your genetic information. However, please be aware that these measures do not protect you against genetic discrimination by companies that sell life insurance, disability insurance, or long-term care insurance.

There is a slight risk of breach of confidentiality, in which your genetic information or other medical information becomes known by others. The VA will make every effort to protect your confidentiality and will take steps to secure your personal and medical information.

You are not required to take medication as a part of this study. However, if your healthcare provider chooses to change your medications based on the information you receive as a part of this study, he or she should discuss with you the potential benefits and harms of those medications, which can include side effects.

Risks of the usual care you receive are not risks of this study. Those risks are not included in this document. You should talk with your health care providers if you have any questions about the risks of usual care.

**WHAT ARE THE POSSIBLE BENEFITS OF THIS STUDY?**

We do not know if you will get any benefits from taking part in this research study. However, the results you receive about the risk of cardiovascular disease in you and your family members might help your healthcare providers make medical decisions to lower your risk.

**WHAT OTHER CHOICES DO I HAVE IF I DO NOT WANT TO JOIN THIS STUDY?**

The only alternative to participating in this study is not to participate. If you choose not to participate, we will not return your research result. In either case, you will continue receiving your usual health care from your providers, including any care you might already be receiving to lower your cholesterol and your risk of cardiovascular disease.

#### WHO WILL SEE MY INFORMATION AND HOW WILL IT BE PROTECTED?

There are rules to protect your private information. Federal and state laws and the federal medical Privacy Rule also protect your privacy. To further protect your privacy, the MVP-ROAR Study has obtained a Certificate of Confidentiality, issued by the National Institutes of Health (NIH). This means that the researchers conducting this study cannot be forced to share information that could identify you, even if required by a court order.

The research team working on the study will collect information about you. This includes things learned from the procedures described in this informational sheet. The study team may also collect other information including your name, address, date of birth, and information from your medical records such as HIV status, drug, alcohol or STD treatment, genetic test results, or mental health treatment.

Your study information will be kept confidential. All data will be stored on secure, password-protected computers, accessible only to authorized research personnel.

We will include some information about your study participation in your medical record.

There are times when we might have to show your records to other people. For example, someone from the Office of Human Research Protections (OHRP), the Government Accountability Office (GAO), the Office of the Inspector General, the VA Office of Research Oversight, the VA Central Institutional Review Board, the local Research and Development Committee, and other study monitors may look at or copy portions of records that identify you. Your information, including your name, date of birth, and gender will be shared with an external, VA-approved clinical laboratory for the purposes of genetic testing to confirm your research result.

A description of this clinical trial will be available on http://www.ClinicalTrials.gov as required by U.S. Law. This website will not include information that can identify you. At most, the website will include a summary of the results. You can search this website at any time.

Your study data, including data that can be used to identify you, will be entered into the VA MVP Central Database and may be used for future research studies approved by an Institutional Review Board (IRB).

#### WHAT ARE THE COSTS TO ME IF I TAKE PART IN THIS STUDY?

You will incur the costs of your time and travel to study appointments. You will not be charged for any treatments or procedures that are part of this study. If you usually pay co-payments for VA care and medications, you will still pay these co-payments for VA care and medications that are not part of this study.

#### WILL I RECEIVE ANY PAYMENT IF I PARTICIPATE IN THIS STUDY?

To compensate you for your time and travel expenses, you will receive $50 upon completion of the survey and blood draw at the end of the study.

#### WHO COULD PROFIT FROM THE STUDY RESULTS?

Your study specimens will not be used for commercial profit.

#### DOES THIS STUDY INVOLVE GENETIC RESEARCH AND HOW WILL MY GENETIC INFORMATION BE PROTECTED?

Yes, this study involves retesting you for a genetic result that was identified in your MVP blood sample. This study will take a new sample of blood or saliva from you and only test it to see if you have the genetic result identified in your MVP results. This research does not involve testing any genes not related to heart health.

You and your primary care provider(s) will receive the results of this genetic test, and it will be entered in your medical record.

There are state, federal, and VA protections that prevent health insurance companies, group health plans, and most employers from discriminating against you based on your genetic information. However, please be aware that these measures do not protect you against genetic discrimination by companies that sell life insurance, disability insurance, or long-term care insurance.

#### RE-CONTACT

You may be contacted in the future by the MVP-ROAR Study team to determine your interest in participating in additional research. Your participation in additional research would be voluntary and would require additional consent at that time.

#### WHO DO I CONTACT ABOUT THIS STUDY IF I HAVE QUESTIONS?

In the event of a research related injury, the VA will provide necessary medical treatment at no cost to you unless the injury is due to non-compliance by a study participant with study procedures or if the research is conducted for VA under contract with an individual or non-VA institution. If you should have a medical concern or get hurt or sick as a result of taking part in this study, you should call your primary care or other healthcare provider.

If you have any questions, comments, or concerns about this research or you want to withdraw from the study, please contact the MVP-ROAR Study at 857-364-3267, Dr. Jason Vassy at 857-364-2561, or your local Patient Advocate.

If you have questions about your rights as a study participant, or you want to make sure this is a valid VA study, you may contact the VA Central Institutional Review Board (IRB). This is the Board that is responsible for overseeing the safety of human participants in this study. You may call the VA Central IRB toll free at 1-877-254-3130 if you have questions, complaints or concerns about the study or if you would like to obtain information or offer input.
