## Supplementary File 12 for "Introducing return of results in the Million Veteran Program: Design and pilot results of the MVP-ROAR Familial Hypercholesterolemia Study"

VA U.S. Department of Veterans Affairs

**Million Veteran Program**
Washington, DC 20420

MVP ROAR Return Of Actionable Results

NAME
ADDRESS 1
ADDRESS 2
CITY, STATE, ZIP

DATE

Dear FULL NAME,

You recently received a letter from the Million Veteran Program (MVP) informing you about a new research study, called **MVP-ROAR (Return Of Actionable Results).** We are sending this letter to give you more information about this opportunity.

Researchers using your MVP blood sample have made a discovery that may be important to the cardiovascular health of you and your family members, including the risk of heart attack and stroke. A team of experts has reviewed the findings and decided that genetic results from your sample might help you, your family members, and your healthcare providers make decisions about your health care. However, research tests are not the same as clinical tests done as a part of medical care. For this reason, it would be necessary to have your research finding repeated in a clinical laboratory. The MVP-ROAR research study offers MVP participants the opportunity to have these research results confirmed in a clinical laboratory and then reported back to you and your healthcare providers.

If you are interested in learning more about the MVP-ROAR study, please read the enclosed consent form closely. A member of our study team will contact you shortly to review the information with you, answer questions, and ask whether you are interested in participating in the study. In the meantime, if you have any questions about the study, please call the MVP-ROAR Study at 857-364-3267.

Participation in the MVP-ROAR study is entirely voluntary and will not affect your health care or any VA benefits you currently receive or are eligible to receive. It will also not affect your enrollment in MVP. Thank you for your consideration as we continue working to improve the health care of Veterans.

Sincerely,

Jason L. Vassy, M.D., M.P.H
*MVP-ROAR Principal Investigator*
