## Supplementary File 13 for "Introducing return of results in the Million Veteran Program: Design and pilot results of the MVP-ROAR Familial Hypercholesterolemia Study"

**Million Veteran Program**
Washington, DC 20420

Dear <Veteran Full Name>,

Greetings from the Million Veteran Program (MVP)!

When you joined MVP, you agreed to be contacted about additional research opportunities. We are contacting you today to provide information about a **new research project** sponsored by the Department of Veterans Affairs (VA) to determine your interest in being contacted by the project team.

The new research project, called **MVP-ROAR (Return Of Actionable Results)** is a new project on heart disease risk. MVP participants in MVP-ROAR, such as yourself, will have the opportunity not only to make additional contributions to our understanding of the risk factors for heart disease but also to receive your own heart disease risk results.

**If you are interested in learning more about MVP-ROAR,** you do not need to respond to this letter. A member of the project team may contact you in the next few weeks to answer any questions you have and to ask whether you might be interested in participating. Please note that being contacted does not obligate you to participate in this new project.

**If you do not wish to be contacted about MVP-ROAR**, we ask you to please do one of the following within the next 2 weeks:

1. Return the enclosed stamped, pre-addressed postcard; or
2. Contact the MVP Info Center at 866-441-6075

If we receive confirmation that you do not wish to be contacted, we will not attempt to contact you again about this new project.

As noted above, participation in the MVP-ROAR is entirely voluntary and will not affect your healthcare or any VA benefits you currently receive or are eligible to receive. It will also not affect your status in MVP.

Thank you for your consideration and continued partnership with MVP.

Sincerely,

J. Michael Gaziano, M.D., M.P.H
MVP Principal Investigator

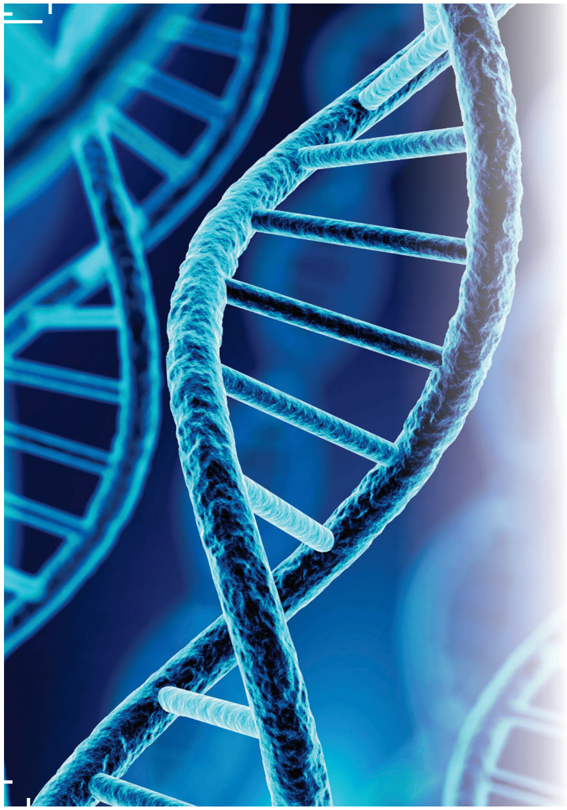

06-28-2019_55

### MILLION VETERAN PROGRAM

The **Million Veteran Program (MVP)** would like to tell you about a new research opportunity, MVP-ROAR. If you do not wish to be contacted, please let us know by returning this card.

- [ ] I do not wish to be contacted to learn more about MVP-ROAR.

Please contact the MVP Info Center with any questions: 866-441-6075.

VA | U.S. Department of Veterans Affairs

___________________
