## Supplementary Figure 1 for "Introducing return of results in the Million Veteran Program: Design and pilot results of the MVP-ROAR Familial Hypercholesterolemia Study"

Funding Agency: VA Office of Research and Development

Principal Investigator: Jason L. Vassy, MD, MPH, SM

Version Number 6

August 26, 2022

VA Central IRB
Effective Date: October 4, 2022

### Abstract

#### Background

In the last 5 years, the genomics research and clinical communities have developed some consensus about which disease-associated genetic results are "actionable" for patients and their healthcare providers. At the same time, professional variant interpretation standards and increased data sharing have increased the validity of genetic variant interpretation. Familial hypercholesterolemia (FH) is an example of an actionable monogenic disease, and validated FH variants are now being identified among participants of the Million Veteran Program (MVP). However, it remains uncertain whether patients and their healthcare providers will use such genetic results to change clinical management.

#### Objectives

The purpose of the MVP-ROAR-FH (Return Of Actionable Results) study is to develop a process to return medically actionable genetic results to living MVP participants and to determine the impact of doing so on medical management and outcomes and Veteran quality of life.

#### Methods

This a randomized controlled trial of immediate vs. delayed (after 6 months) return of FH variant results. After first being given the opportunity to opt out of participating from the MVP Core study team, living MVP participants with an actionable FH variant are contacted by the MVP-ROAR-FH genetic counselor (GC). The GC discusses the fact that MVP researchers have identified a genetic result that might give the participant and his/her healthcare providers information about his/her risk of heart disease from cholesterol but that this is a research result and would have to be confirmed in a clinical laboratory. After consenting to participate in the study, the participant completes a baseline survey and presents to his/her local VA facility for a blood draw or provides a saliva sample using a self-collection kit. After confirmed biospecimen collection by study staff, participants are randomly allocated to the Immediate Results or Delayed Results arm. Baseline cholesterol values from all participants are measured from blood specimens or, in the absence of a blood draw, are obtained from the medical record. Biospecimens (blood or saliva) from participants in the Immediate Results arm are sent for FH variant confirmation. Upon receiving the variant confirmation results, the GC calls participants in the Immediate Results arm to deliver the study intervention at baseline. Briefly, the GC lets the participant know the results of the FH variant confirmation and delivers standard post-test genetic counseling, including the provision of FH-related resources and other information and facilitation of cascade genetic testing of family members, if appropriate. The GC also sends the results and physician-level materials about FH to the participant's primary care provider (PCP). Participants in the Delayed Results group receive only their cholesterol results at baseline. During the 6 months after enrollment, participants in both arms continue receiving usual care from their PCPs and other healthcare providers as usual. After 6 months, participants in both arms complete a follow-up survey and undergo an end of study blood draw for repeat cholesterol panel testing and, for participants in the Delayed Results arm, FH variant confirmation testing using either blood or saliva. After their end-of-study data collection is complete, the GC contacts participants in the Delayed Results arm to deliver the same study intervention the Immediate Results arm received. A subset of PCPs whose patients received results from MVP-ROAR will be invited to participate in qualitative interviews about their experience with the project and to identify




VA Central IRB
Effective Date: October 4, 2022

facilitators and barriers to effective genetic return of results in Veteran healthcare.  The primary hypothesis is that the reduction in low-density lipoprotein cholesterol after 6 months will be greater in the Immediate Results arm compared to the Delayed Results arm. Secondary outcomes include changes in medications, cascade genetic testing among family members, quality of life, and healthcare costs.

**Anticipated Impact on Veteran Healthcare**

This project has the potential to improve the health care and health outcomes for MVP participants with FH variants while also generating generalizable knowledge about the processes and outcomes of returning genetic results to research participants. The processes developed and studied in this project could inform best practices for the return of genetic results in MVP participants in the VA for other conditions.



VA Central IRB
Effective Date: October 4, 2022

### List of Abbreviations

| | |
|---|---|
| ACC | American College of Cardiology |
| ACMG | American College of Medical Genetics and Genomics |
| AE | Adverse event |
| AHA | American Heart Association |
| AMP | Association of Molecular Pathology |
| *APOB* | *Apolipoprotein B* gene |
| B | Benign |
| CHD | Coronary heart disease |
| CLIA | Clinical Laboratory Improvement Amendments |
| CVD | Cardiovascular disease |
| FDA | Food & Drug Administration |
| FH | Familial hypercholesterolemia |
| GC | Genetic counselor |
| GMS | Genomic Medicine Service |
| HERC | Health Economics Resource Center |
| III | Individually identifiable information |
| LB | Likely benign |
| LDL-C | Low-density lipoprotein cholesterol |
| *LDLR* | *Low-density lipoprotein receptor* gene |
| LP | Likely pathogenic |
| MCA | Managerial Cost Accounting |
| MOU | Memorandum of Understanding |
| MVP | Million Veteran Program |
| P | Pathogenic |
| PCP | Primary care provider |
| *PCSK9* | *Proprotein convertase subtilisin/kexin type 9* gene |
| PHI | Protected health information |
| RCT | Randomized controlled trial |
| ROAR | Return of actionable results |
| ROR | Return-of-results |
| SAE | Serious adverse event |
| *SLCO1B1* | *Solute carrier organic anion transporter family member 1B1* gene |
| SOP | Standard operating procedure |
| UP | Unanticipated problem |
| VA | Veterans Affairs |
| VUS | Variant of uncertain significance |



VA Central IRB
Effective Date: October 4, 2022

### Table of Contents


1.0 Study Personnel 7

2.0 Introduction 8

3.0 Objectives 10

4.0 Resources and Personnel 12

5.0 Study Procedures 14

5.1 Study Design 14

5.2. Recruitment Methods 21

5.3 Informed Consent Procedures 22

5.4 Inclusion/Exclusion Criteria 23

5.5 Study Evaluations 23

5.6 Data Analysis 26

5.7 Withdrawal of Subjects 27

6.0 Reporting 27

7.0 Privacy and Confidentiality 29

8.0 Communication Plan 31

9.0 References 32





VA Central IRB
Effective Date: October 4, 2022

### Appendices

Living MVP Participants with FH Variants (LDLR)

MVP-ROAR-FH Informed Consent Letter

MVP-ROAR-FH Informational Sheet

MVP-ROAR-FH Informational Sheet (Pilot)

MVP-ROAR-FH GC IC Phone Script

MVP-ROAR-FH Instructions for Saliva Collection

MVP-ROAR-FH Baseline Survey

MVP-ROAR-FH Survey Cover Letter

MVP-ROAR-FH Loss to Follow-Up Letter (General / Result version)

MVP-ROAR-FH Baseline Letter to Delayed Results Arm

MVP-ROAR-FH Physician Letter (Delayed Results)

MVP-ROAR-FH 6-Month Survey (Immediate Results version)

MVP-ROAR-FH 6-Month Survey (Delayed Results version)

MVP-ROAR-FH Immediate Results 6-Month Letter

MVP-ROAR-FH Physician Immediate Results 6-Month Letter

Sample Lipid Report

MVP-ROAR-FH GC Result Delivery Process

MVP-ROAR-FH Patient Results Letter (Positive / Negative version)

Clinical Variant Report

MVP-ROAR-FH Family Letter

MVP-ROAR-FH Physician Letter (Immediate Results) (Positive / Negative version)

MVP-ROAR-FH Sample Post-counseling CPRS Note

MVP-ROAR-FH Study Withdrawal Letter

MVP-ROAR-FH PCP Email Invitation

MVP-ROAT-FH Draft Interview Guide



VA Central IRB
Effective Date: October 4, 2022

**Protocol Title:** Million Veteran Program Return Of Actionable Results (MVP-ROAR-FH)

### 1.0 Study Personnel

1.1 Principal Investigator:

- Jason Vassy, MD, MPH, SM
  Clinician-Investigator, Section of General Internal Medicine,
  VA Boston Healthcare System
  Associate Professor of Medicine, Harvard Medical School

1.2 Executive committee for the study:

The executive committee will conduct ongoing scientific and operational review of study activities. This committee includes an interdisciplinary collection of clinicians and researchers with expertise in FH, clinical cardiology, genomic medicine, epidemiology, biostatistics, genetic return-of-results, genetic counseling, and econometrics. This committee will meet virtually biweekly, chaired by Dr. Vassy. Members of the executive committee:

- Themistocles (Tim) L. Assimes, MD, PhD
  Associate Director, Epidemiology Research and Information Center for Genomics,
  VA Palo Alto Health Care System
  Associate Professor of Medicine, Stanford University

- Charles A. Brunette, PhD
  Health Science Specialist
  VA Boston Healthcare System

- Kurt D. Christensen, MPH, PhD
  Instructor of Medicine, Harvard Medical School and Brigham and Women's Hospital

- Morgan Danowski, MS, LCGC
  Genetic Counselor
  VA Boston Healthcare System

- Qin Hui, MS
  Genetic Data Analyst, Rollins School of Public Health, Emory University

- Joshua W. Knowles, MD, PhD
  Chief Research Advisor, FH Foundation
  Assistant Professor of Medicine, Stanford University



VA Central IRB
Effective Date: October 4, 2022

- Pradeep Natarajan, MD, MMSc

  Director of Preventive Cardiology at Massachusetts General Hospital
  Assistant Professor of Medicine, Harvard Medical School

- Amy Sturm, MS, LCGC

  Professor and Director, Cardiovascular Genomic Counseling, Geisinger Health

- Yan Sun, PhD

  Associate Professor of Epidemiology, Emory University

- Virginia Morrison, MS, LCGC

  Genetic Counselor, Genomic Medicine Service,
  VA Salt Lake City Health Care System

- Peter Wilson W.F. Wilson, MD

  Director of Epidemiology and Genomic Medicine, Atlanta VA Medical Center
  Professor of Medicine, Division of Cardiology, Emory University

<u>1.3 Potential participating sites</u>

Because eligible participants for this study include Million Veteran Program (MVP) participants found to have potentially actionable genetic variants, participants and their PCPs may be drawn from more than 50 VA locations, primarily but not limited to those with high MVP enrollment (see Living MVP Participants with FH Variants (*LDLR*)). Study procedures, including recruitment, consent, delivery of the intervention, and data collection will be conducted centrally by the MVP-ROAR study team. MVP local site investigators will be notified each time a participant is enrolled from their site. Recruitment start-up will be graduated. A non-randomized pilot trial will be conducted among 10 eligible MVP participants from the VA Boston Healthcare System and 1-2 other VA locations before initiation of the larger randomized controlled trial (RCT) across the entire MVP cohort.

### 2.0 Introduction

<u>2.1 Return of results to research biobank participants</u>

Since 2011, MVP has enrolled participants who receive clinical care in the VA healthcare system to understand role of genetics in health[1]. Participants provide broad consent to use their samples and survey and healthcare data for research and consent to be contacted in the future about additional research opportunities. One of the opportunities afforded by biobanks linked to integrated healthcare systems is the ability to screen for genetic risk factors that, if reported to participants and their healthcare providers, may inform their health care and improve health outcomes. Biobank participants routinely state that they would desire and expect actionable medical findings to be returned for their benefit[2,3]. The ethics and practicalities of such return are



VA Central IRB
Effective Date: October 4, 2022

complicated, but there is an emerging consensus that biobanks should consider reporting clinically validated results in genes deemed medically actionable[4-8].

2.2 Familial hypercholesterolemia: A medically actionable genetic diagnosis

A common monogenic condition, familial hypercholesterolemia (FH) is an ideal test case for piloting genetic return of results in MVP. Cardiovascular disease, including coronary heart disease (CHD) is the leading cause of death in Veterans. Elevated low-density lipoprotein cholesterol (LDL-C) level is an important, prevalent, and modifiable CHD risk factor[9]. Characterized by extreme LDL-C elevation, FH occurs in approximately 1 in 250 in the US[10-12] and markedly raises premature CHD risk, independent of cross-sectional LDL-C values[10,13,14]. Familial hypercholesterolemia can be caused by variants in the *low-density lipoprotein receptor* (*LDLR*), *apolipoprotein B* (*APOB*), *proprotein convertase subtilisin/kexin type 9* (*PCSK9*) and *low-density lipoprotein receptor adaptor protein 1* (*LDLRAP1)* genes. In current practice, FH may be identified through routine lipid testing, but only 1 in 50 individuals with severe hypercholesterolemia have FH mutations and some FH mutation carriers may not have severely elevated LDL-C. However, across LDL-C values, even when not markedly elevated, FH mutation status portends increased risk of premature CHD. Cholesterol values seen in common hypercholesterolemia can overlap considerably with those seen in FH, particularly in middle-aged and older adults[15,16]. As a result, an estimated 90% of FH cases in the US remain undiagnosed[17].

Compared to common hypercholesterolemia, an FH diagnosis changes prognosis and management, as FH individuals have much greater CHD risk than would be predicted by usual risk models and require earlier and more aggressive therapy and surveillance. Individuals with FH have a 10 to 20-fold higher lifetime CHD risk than those without. Even compared to individuals with equivalent LDL-C levels at a single measurement, FH heterozygotes have a 3-fold higher of CHD compared to matched patients without an FH variant[14,18,19]. Contemporary observational analyses indicate a greater relative and absolute clinical benefit of LDL-C-lowering with statins among those with FH mutations[20]. Thus, the American Heart Association and American College of Cardiology recommend intensive LDL-C lowering (<100 mg/dL or <70 mg/dL, depending on other risk factors) when FH is present, which might necessitate statin dose escalation or the addition of ezetimibe or a PCSK9 inhibitor to high-dose statin therapy[21,22].

Diagnosing FH among MVP participants and distinguishing it from common hypercholesterolemia thus has the potential to improve the healthcare of the Veteran. Considerable data suggest that many patients with FH are undiagnosed and undertreated, resulting in many potentially preventable myocardial infarctions and deaths[23-28]. Indeed, in preparatory-to-research analyses for this proposal, only 240/642 (37%) MVP participants with a potential FH variant in one of the 3 FH genes have a most recent LDL-C <100mg/dL, and only 397/642 (62%) have been prescribed a statin of any dose in the last year. This is consistent with observations in other healthcare settings[29,30]. Moreover, some healthcare providers remain skeptical that a genetic diagnosis of FH should change clinical management over and above non-genetic approaches to risk prediction and management.

Identifying an FH variant in an individual also carries family implications. National registry projects demonstrated the benefits of systematic FH case-finding with genetic screening and cascade testing in family members and show that such an approach may lead to more complete capture, earlier treatment of undiagnosed FH, and cost-effective CHD risk reduction[15,17,31-34]. Thus, several professional organizations and the Centers for Disease Control and Prevention endorse cascade screening among first-degree relatives of patients with an FH variant as an



VA Central IRB
Effective Date: October 4, 2022

effective genomic medicine intervention with high evidence of clinical utility[17,21]. Because of its actionability, FH is on the American College of Medical Genetics and Genomics (ACMG) list of reportable monogenic conditions[35]. Thus, identification of MVP participants has the potential to improve the lives not only of the Veteran but also their children, siblings and other family members.

2.3 Emerging consensus around FH variant interpretation and management

Clinical laboratories currently use American College of Medical Genetics and Genomics (ACMG) - Association for Molecular Pathology (AMP) standards and guidelines to classify individual genetic variants into one of five pathogenicity categories for a given disease: benign (B), likely benign (LB), variant of uncertain significance (VUS), likely pathogenic (LP), or pathogenic (P)[36]. In clinical testing, it is standard of care to report only P and LP variants back to ordering providers and patients when unexpected actionable genetic variants are identified[35,37]. The ACMG-AMP standards ask laboratories to apply a set of 28 criteria to classify each variant, using evidence such as population data, computational data, functional data, and segregation data. Once a laboratory interprets a given variant, it is encouraged to share that interpretation in a publicly accessible database such as ClinVar[38], along with any criteria it used to make its determination of pathogenicity. Because there is some subjectivity in how laboratories apply the ACMG-AMP criteria[39], some variants have conflicting interpretations in ClinVar. ClinVar uses a 4-star rating system for the variant interpretations in its database, corresponding to the level of evidence behind each variant classification. Recognizing that content expertise is required to adjudicate the application of ACMG-AMP criteria to specific disease-gene associations, the NIH-funded Clinical Genome Resource (ClinGen) was created in part to implement evidence-based disease expert consensus for curating genes and variants[40]. ClinGen organizes expert curation groups for individual diseases, such as FH. These expert curation groups develop consensus for applying ACMG-AMP criteria in classifying variant pathogenicity for specific diseases and their associated genes. Only variants that are classified by such expert panels can achieve a 3- or 4-star interpretation in ClinVar.

This project will use ClinVar data and the ongoing curation work of the ClinGen Cardiovascular Familial Hypercholesterolemia Variant Curation Expert Panel[41,42] (co-led by Dr. Knowles) to ensure that only variants with high-quality LP or P classifications are returned to participants. As of May 2019, the panel's FH-specific variant criteria have been finalized and are awaiting ClinGen approval, after which the panel will begin to systematically review the >2000 *LDLR* variants listed in ClinVar. We will only consider returning FH variants classified as LP or P according to ACMG-AMP for this project. Thus, it is important to note that, although these are research-derived results, their interpretation will meet clinical standards. Indeed, in December 2018 the US Food and Drug Administration (FDA) formally recognized genetic variant information from the ClinGen Expert Curated Human Genetic Database as a source of valid scientific evidence that can be used to support clinical validity in premarket submissions for diagnostics tests, the first database to receive such recognition.

Although FH may go undiagnosed in routine care, the risk of hypercholesterolemia for CHD in general is widely appreciated in the medical community and among patients. As a result, gaps in genetic literacy may pose less of a barrier for FH return-of-results versus other monogenic conditions. Professional FH guidelines and educational resources exist to help patients and providers manage an FH diagnosis, and these resources will be used in this project to support the responsible and clinically meaningful return of genetic results to MVP participants.



VA Central IRB
Effective Date: October 4, 2022

For all the reasons above, FH is the ideal test case for piloting genetic return-of-results in MVP. Although the ACMG lists FH among its actionable conditions and there is emerging expert consensus about how to interpret and manage FH variants, it is unknown whether participants in a biobank study like MVP and their healthcare providers will be receptive to the information and change clinical management accordingly. Moreover, the return-of-results process may bring unanticipated harms, such as distress or anxiety, that would be important to quantify and address. Thus, there is equipoise in whether actionable genetic results should be returned to biobank participants. This study will use a randomized design to test the hypothesis that returning FH variants to MVP participants and their providers will result in lower LDL cholesterol levels. If the return-of-results process described in this protocol does not change clinical management or results in undue participant distress, further study will be needed to improve the process and achieve net benefit to patient-participants.

### 3.0 Objectives

#### <u>3.1 Study purpose/aims</u>

The purpose of this study is to develop a process to return medically actionable genetic results to living MVP participants and determine the impact of doing so on medical outcomes, Veteran quality of life, and healthcare costs. We will determine this impact by using an RCT of reporting Immediate Results vs. Delayed Results (after 6 months) to test the hypotheses in Section 3.2.

#### <u>3.2 Study outcomes and hypotheses</u>

##### 3.2.1. Primary outcome

The primary outcome will be the change in LDL-C from study baseline to the end of study (6 months after enrollment). We will test the hypothesis that the LDL-C reduction after 6 months will be greater in the Immediate Results arm compared to the Delayed Results arm.

##### 3.2.2. Secondary outcomes

We will test the following secondary hypotheses:

1. The proportion of participants meeting clinically significant LDL-C targets (< 100mg/dL for primary prevention and <70 mg/dL for secondary prevention) at 6 months will be greater in the Immediate Results arm than in the Delayed Results arm.
2. The proportion of participants with an intensification of lipid-lowering pharmacotherapy will be greater in the Immediate Results arm than in the Delayed Results arm. This composite outcome will include prescription of new monotherapy, dose escalation of existing pharmacotherapy, and addition of one or more medications to existing pharmacotherapy.

##### 3.2.3. Exploratory outcomes

We will test the following exploratory hypotheses:

1. Medication adherence at 6 months will be higher in the Immediate Results arm than in the Delayed Results arm.
2. Participants in the Immediate Results arm will report a greater number of first-degree relatives having undergone genetic testing at 6 months than in the Delayed Results arm.



VA Central IRB
Effective Date: October 4, 2022

3. A greater proportion of participants in the Immediate Arm will have healthy lifestyle behaviors (smoking, physical activity, and saturated fat intake) at 6 months than in the Delayed Results arm.

3.2.4. Economic outcomes

We will perform a budget impact analysis of the intervention alongside the randomized trial, as described in Section 5.6 Data Analysis below.

3.2.5 Provider outcomes

We will describe VHA primary care providers' perceptions of the facilitators and barriers of return of actionable genetic results to MVP participants nationwide.

3.3 Relevance to Veterans and VA

This project has the potential to improve the health care and health outcomes for MVP participants with FH variants while also generating generalizable knowledge about the processes and outcomes of returning genetic results to research participants.

The reporting of potentially pathogenic FH variants to MVP participants and their healthcare providers might impact participants' clinical outcomes in the following ways. First, for Veterans without prior cholesterol testing, receipt of such a variant could prompt them and their providers to initiate routinely recommended healthcare screening. Given the high rates of cholesterol screening in the VA, a small proportion of participants are likely to fall into this category[43]. Second, for Veterans with prior elevated cholesterol results not already on therapy, disclosure of an FH variant result might prompt initiation of statin therapy after discussion with their healthcare provider. Third, an FH variant result will identify participants who are already on lipid-lowering therapy but are undertreated for their level of CHD risk per current guidelines. These participants and their providers might intensify current lipid-lowering regimens, by maximizing statin intensity (medication and dose) or adding additional agents such as ezetimibe or a PCSK9 inhibitor as necessary. The end result for all of these pathways would be an overall reduction in LDL-C values among participants receiving FH variant results, mediated by a change in therapy. Since LDL-C is a well-recognized causal mediator of CHD, LDL-C reduction is an established surrogate endpoint for CHD risk reduction. Beyond the MVP participants themselves, an important additional benefit of reporting FH variants may be a greater uptake of family-based screening among these Veterans' family members.

Moreover, the processes developed and studied in this project could inform best practices for the return of genetic results in MVP participants in the VA for other conditions.

### 4.0 Resources and Personnel

4.1 Study personnel

All study personnel listed below will have access to protected health information.

4.1.1 Principal investigator



VA Central IRB
Effective Date: October 4, 2022

The principal investigator (PI) supervises all aspects of the study. The PI takes ultimate responsibility for the conduct of the study, including meeting study goals, monitoring of participant safety, and dissemination of findings.

4.1.2 Project manager

The project manager (PM) oversees the day-to-day operations of the study, under the supervision of the PI. The PM, in conjunction with the PI and relevant study personnel, develops and maintains study-related standard operating procedures (SOPs). The PM is the primary point of contact between the study personnel and the IRB and works with the IRB to maintain approval for the study protocol and associated documents, including relevant correspondence for protocol modifications, continuing reviews, regulatory audits, and event reporting. The PM coordinates and leads regular meetings among the study personnel and collaborators, including preparing meeting agendas and minutes. The PM oversees the study budget and is the primary point of contact between the study and vendors. The PM oversees records management for all study-related documents and materials. The PM may delegate some of these tasks to the research assistant as appropriate. The PM escalates study-related problems to the PI, including any participant safety concerns identified.

4.1.3 Genetic counselor

The genetic counselor (GC) designs and delivers the study intervention to participants and their PCPs. The GC works with the PI and PM to develop the genetic counseling-related materials for review by the IRB. The GC works with the PM to develop the SOP for each step in the return-of-results process, including the ordering of variant confirmation testing, sample acquisition and shipping for variant confirmation testing, receipt of variant confirmation results, reporting of results and associated supportive information to participants and their PCPs, and entry of genetic results into the participants' medical record. The GC describes the study to participants and obtains informed consent. The GC may administer the baseline survey, if conducted at the same time as informed consent. The GC delivers the post-test genetic counseling intervention and works with participants to facilitate cascade genetic testing of family members, as appropriate. The GC is available for follow-up questions by participants and their healthcare providers. The GC is the primary point of contact between the study and the VA-approved, CLIA-certified laboratory used for FH variant confirmation. The GC escalates clinical questions/concerns to the PI, including any participant safety concerns, and escalates research-related questions to the PM.

4.1.4 Senior genetic counselor

The senior GC is a practicing genetic counselor in the VA healthcare system. As applicable, the senior GC guides the use of existing clinical genomic medicine services to facilitate the clinical variant confirmation testing and reporting.

4.1.5. Research assistant

The research assistant (RA) performs study tasks under the supervision of the PM. These include literature reviews; drafting and formatting of IRB and other regulatory documents; preparation of materials for study meetings; formatting of data tables and figures; and drafting of research posters and presentations. The RA performs initial



VA Central IRB
Effective Date: October 4, 2022

participant eligibility screening by chart review, escalating any questions to the GC. Under the direction of the PM, the RA sends participant mailings at appropriate times. In collaboration with the GC, the RA communicates with local sites to make arrangements for participant blood draws, to facilitate entry of genetic results into the medical record, and to prepare letters and accompanying materials for patients, their families, and PCPs. The RA administers the baseline and end-of-study participant surveys. The RA facilitates the distribution of participation incentives to participants. The RA escalates clinical questions to the GC and research questions to the PM.

4.1.6. Statistician

The statistician advises the study team on the appropriate study design and statistical analyses for the study outcomes. The statistician conducts sample size and power calculations and supervises the data analyst in performing statistical analysis of study data.

4.1.7 Data analyst

The data analyst (DA) creates and maintains the database housing the study data. The DA creates a data capture system to collect and merge data from participant surveys, medical records, and other VA databases. The DA ensures that data capture and management systems comply with required data security standards. The DA prepares summary data tables for study planning, reporting, monitoring, and dissemination of results. Under the direction of the statistician, the DA performs statistical analyses of study outcomes.

4.2 Services

The study staff will partner with VA Genomic Medicine Service (GMS) to facilitate clinical genetic confirmation testing and report processing for the MVP-ROAR-FH Study. The GMS will provide services and support for FH variant confirmation testing as arranged for MVP-ROAR-FH participants by study staff. GMS uses the VA's telehealth infrastructure to provide clinical genetic services to Veterans in over 90 facilities throughout the country. Part of their service includes an ability to order germline genetic tests for Veterans, with an established process to facilitate testing with commercial laboratories. As applicable, study staff will use established GMS processes to carry out clinical confirmation testing with a VA-approved CLIA-certified laboratory. Biospecimen samples will be labeled with participant name, date of birth, gender, and relevant variant for clinical confirmation, and shipped to the VA Boston Healthcare System clinical laboratory for lipid testing and dissemination to the VA-approved CLIA-certified laboratory for genetic confirmation testing.

### 5.0 Study Procedures

5.0.1 Study timeline

| | **Year 1** | | | | **Year 2** | | | | **Year 3** | |
|---|---|---|---|---|---|---|---|---|---|---|
| | **Q1** | **Q2** | **Q3** | **Q4** | **Q 1** | **Q2** | **Q3** | **Q4** | **Q1** | **Q2** |



VA Central IRB
Effective Date: October 4, 2022

| | | | | | | | | | | |
|---|---|---|---|---|---|---|---|---|---|---|
| **Non-randomized pilot testing of study intervention**<br>(10 eligible MVP participants) | ■ | | | | | | | | | |
| **Patient recruitment and enrollment**<br>(20-21 patients per month x 12 months = 244) | | ■ | ■ | ■ | ■ | | | | | |
| **Return of results to patients in immediate results arm** | | ■ | ■ | ■ | ■ | | | | | |
| **Return of results to patients in delayed results arm** | | | | ■ | ■ | ■ | ■ | | | |
| **Patient data collection**<br>(6-month outcomes; EHR and survey data) | | | | ■ | ■ | ■ | ■ | ■ | | |
| **Data preparation and analysis for clinical outcomes** | | | | | | | | ■ | | |
| **Report on primary outcomes to ORD** | | | | | | | | ■ | | |
| **Data preparation and analysis for economic outcomes** | | | | | | | | | ■ | ■ |
| **Participant and provider qualitative interviews and analysis** | | | | | | | ■ | ■ | ■ | ■ |
| **Dissemination of results** | | | | | | | | | ■ | ■ |

### 5.1 Study Design

#### 5.1.1. Study overview

This project is an RCT of immediate versus delayed (after 6 months) reporting of clinically confirmed FH variants among 244 MVP participants. In an initial non-randomized pilot phase of the project, the below procedures will be piloted among 10 eligible participants, all of whom will receive their results immediately at baseline.

Figure 1 illustrates the RCT study design, which is further described in the relevant sections below. In brief, the study procedures include the following:

- Living MVP participants with an eligible FH variant are mailed a letter from the MVP Core study team, introducing this new MVP-related study and giving participants the opportunity to opt out of further contact about the study by returning a prepaid opt out postcard or by calling the MVP Call Center.
- To any participant who does not opt out within 2 weeks of this initial mailing, the MVP-ROAR-FH study team mails a letter giving more detail about the study, including all necessary informed consent information (see MVP-ROAR-FH Informed Consent Letter and MVP-ROAR-FH Informational Sheet). Participants in the non-randomized pilot of the study procedures will be mailed a letter introducing the study as well as an informational document including all elements of informed consent (see MVP-ROAR-FH Informed Consent Letter and MVP-ROAR-FH Informational Sheet (Pilot)).
- Two weeks after this mailing, the study genetic counselor calls the participant to review the informed consent information, answers any questions about the study, and documents verbal consent or decline (see MVP-ROAR-FH GC IC Phone Script). Study staff also conduct the baseline survey (see MVP-ROAR-FH Baseline Survey) on this phone call or on another call scheduled at a convenient time for the participant. Participants may complete the baseline survey in paper form by request (see MVP-ROAR-FH Survey Cover Letter). Potentially eligible participants not reached after at least three contact attempts by phone



VA Central IRB
Effective Date: October 4, 2022

will be sent a loss to follow-up letter (MVP-ROAR-FH Loss to Follow-Up Letter) via certified mail.

- The genetic counselor works with each consented participant and local laboratory staff to facilitate the collection of biospecimen(s). Two tubes of blood will be collected at baseline, one for baseline lipid panel testing and a second for potential CLIA-certified FH variant confirmation (performed by a commercial genetic laboratory). Alternatively, a saliva sample may be collected remotely from the participant for FH variant confirmation using a self-collection kit. Participants may utilize this option if they have undergone lipid testing at their local VA facility within the last 6 months and without a subsequent change in lipid-lowering medications.
- After the MVP-ROAR-FH study staff confirms baseline biospecimen collection, participants in the RCT are randomly assigned to the Immediate Results arm or the Delayed Results arm. (Participants in the non-randomized pilot will receive their results immediately at baseline).

- <u>Immediate Results arm</u>: The participant's baseline specimen for variant confirmation is shipped to the reference laboratory for clinical variant confirmation of their research results. The laboratory returns these results to the genetic counselor, who schedules a telephone or video visit with the participant, to deliver the intervention described in Section 5.1.3 below. Briefly, he/she lets the participant know the results of the FH variant confirmation and delivers standard post-test genetic counseling, including the provision of FH-related resources and other information. This genetic counseling session is neither audio- nor video-recorded. If the Veteran has living family members, he/she will also educate and provide resources for cascade genetic testing of those family members. The genetic counselor also sends the genetic and cholesterol results and physician-level materials about FH to the participant's PCP. Participants not reached after at least three contact attempts by phone will be sent a loss to follow-up letter (MVP-ROAR-FH Loss to Follow-Up Letter_Result) via certified mail.
    - <u>Delayed Results arm</u>: The participant's baseline blood specimen for variant confirmation is discarded. If collected using saliva, the participant's baseline specimen will be stored



VA Central IRB
Effective Date: October 4, 2022

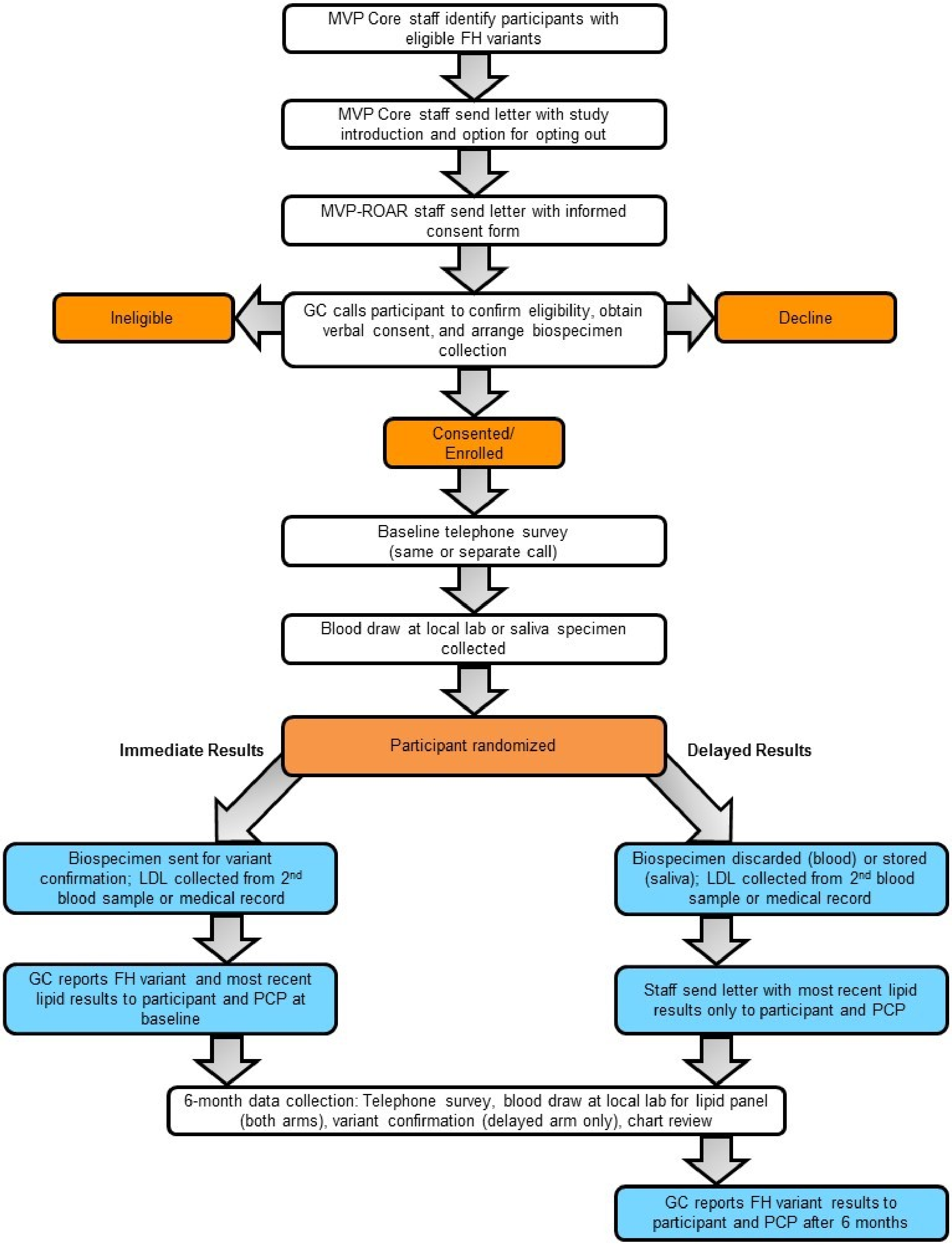


**Figure 1: MVP-ROAR study design. Abbreviations:** BL, baseline; FH, familial hypercholesterolemia; GC, genetic counselor; MVP, Million Veteran Program; PCP, primary care provider; ROAR, Return Of Actionable Results



VA Central IRB
Effective Date: October 4, 2022

in a locked and secure cabinet for six months, at which time the sample may be shipped to the laboratory for variant confirmation The study staff sends the participant a letter letting them know that they were assigned to the delayed results group and that they will be contacted in 6 months to have their research result confirmed (see MVP-ROAR-FH Baseline Letter to Delayed Results Arm). This letter also includes the participant's baseline cholesterol results. A copy of this letter is also sent to the participant's PCP (see MVP-ROAR-FH Physician Letter (Delayed Results)).

- During the 6 months after enrollment, participants will continue receiving usual care from their PCPs and other healthcare providers. As discussed below in Section 5.1.3, PCPs of patients in the Immediate Results arm may choose to change the patient's treatment or refer the patient to a specialist.
- Six months after randomization, study staff will contact each participant to conduct an end-of-study telephone survey (see MVP-ROAR-FH 6-Month Survey (Immediate Results version) and MVP-ROAR-FH 6-Month Survey (Delayed Results version)) and arrange for the participant to have end-of-study fasting lipid panel testing at their local laboratory. A second specimen, either blood or saliva, will be collected from participants in the Delayed Results arm for clinical variant confirmation testing. Participants may complete the follow-up survey in paper form by request (see MVP-ROAR-FH Survey Cover Letter).
- After their end-of-study data collection is complete, the genetic counselor contacts participants in the Delayed Results arm to deliver the same study intervention the Immediate Results arm received at baseline. Participants not reached after at least three contact attempts by phone will be sent a loss to follow-up letter (MVP-ROAR-FH Loss to Follow-Up Letter_Result) via certified mail.
- After their end-of-study data collection is complete, participants in the Immediate Results arm receive a letter with their follow-up cholesterol results (see MVP-ROAR-FH Immediate Results 6-Month Letter, MVP-ROAR-FH Physician Immediate Results 6-Month Letter, and MVP-ROAR-FH Sample Lipid Report)
- Study staff will recruit participant's VA PCPs for interviews about the barriers and benefits they perceive in genetic return-of-results. The interviews will be conducted, recorded, and transcribed via Microsoft Teams, a VA-approved system.

5.1.2. Usual care

In this project, usual care is defined as the current approach to screening and management of hypercholesterolemia across the VA healthcare system. Currently, VA locations already achieve high rates of cholesterol screening in their general patient population[43]. Using guidelines such as the 2014 VA/DoD Clinical Practice Guideline for the Management of Dyslipidemia for Cardiovascular Risk Reduction or the 2018 ACC/AHA Multisociety Guideline on the Management of Blood Cholesterol, VA clinicians use cholesterol results, along with other risk factors including blood pressure, diabetes status, and smoking status, to determine whether a Veteran's CVD risk is high enough to merit consideration of treatment with a statin medication. Without other CVD risk factors, VA guidelines do not recommend treating based on elevated LDL-C levels alone unless they are ≥190 mg/dL. Depending on a Veteran's CVD risk, a VA clinician might recommend dietary modification and/or initiate therapy with a statin, such as atorvastatin, simvastatin, or rosuvastatin. Current guidelines recommend that providers repeat cholesterol testing periodically after initiation of therapy to monitor LDL-C reduction. For patients



VA Central IRB
Effective Date: October 4, 2022

for whom maximum-dose statin therapy is not tolerated or does not achieve sufficient LDL-C reduction, providers might replace or add other medications such as ezetimibe or a PCSK9 inhibitor.

As discussed in Section 2.2, the guidelines for cholesterol management differ significantly for individuals with FH, although most of these individuals go clinically unrecognized. FH can be clinically recognized without genetic testing using varying definitions, such as the Simon Broome Register criteria, Dutch Lipid Clinic Network criteria, and Make Early Diagnosis to Prevent Early Deaths criteria[19]. However, clinical definitions of FH rely on data that are not necessarily collected systematically in busy clinical practice, such as a detailed family history of hypercholesterolemia or coronary heart disease and the presence of tendinous xanthomata or arcus cornealis on physical examination[19,20]. Recently, an expert consensus panel recommended that genetic testing for FH variants in the *LDLR*, *APOB*, and *PCSK9* genes be performed for any patient meeting a clinical definition for “probable” or “definite” FH[34], but these recommendations have not been widely adopted. Moreover, not all FH patients have an LDL-C ≥190 mg/dL at a given measurement in time. In a recent analysis of data from the Geisinger Healthcare System, only 55% of patients with an FH variant had a maximum LDL-C ≥190 mg/dL in their medical record. Nonetheless, we know that such individuals have a 3-fold higher of CHD compared to matched patients without an FH variant and would thus be recommended to be treated with lipid-lower therapy to a target LDL-C <100mg/dL. Among 325 living MVP participants with a potentially pathogenic *LDLR* variant, 177 (54%) have a most recent LDL-C > 100 mg/dL.

In this context, <u>usual care</u> in this project includes the following:

- VA clinicians, most commonly PCPs, order periodic cholesterol testing for their patients.
- Based on cholesterol results, plus other risk factors including age, sex, blood pressure, diabetes status, prior CVD, and smoking status, PCPs consider initiating treatment with lifestyle modification with or without pharmacotherapy, including statins or other agents.
- PCPs follow these patients at regular intervals, periodically repeating cholesterol testing to monitor for medication adherence and appropriate LDL-C response. PCPs may escalate therapy (*e.g.* increasing the dose of statin therapy or adding ezetimibe to statin therapy) for patients not meeting their LDL-C goals.
- There is significant variability in patients’ willingness to take statin or other medications.
- Depending on individual and regional practice patterns and availability, PCPs may refer certain patients to a preventive cardiology or lipid clinic, if cholesterol results are particularly abnormal or if there is some other concerning feature relevant to the patient’s CVD risk.
- The astute PCP or specialist may perform a detailed physical examination and family history relevant to FH and apply FH clinical criteria to determine whether the patient has a clinical diagnosis of FH.
- For patients meeting clinical criteria for FH, some providers may consider genetic testing for FH variants to make a specific molecular diagnosis of FH (*e.g.* identifying the specific *LDLR* variant causing their disease). This would typically be performed with pre- and post-test genetic counseling, to discuss test results interpretation and implications for family members.



VA Central IRB
Effective Date: October 4, 2022

5.1.3. Intervention

In contrast to usual care, participants enroll in the MVP-ROAR-FH study with the knowledge that analysis of their MVP research sample potentially included information about their heart disease risk that might be useful for them, their healthcare providers, and their family members.

Participants randomly assigned to the Immediate Results arm receive the following study intervention at baseline:

- Participants receive the results of their FH variant confirmation from the research genetic counselor (GC) via telephone or videoconferencing (see MVP-ROAR-FH GC Result Delivery Process). This genetic counseling session includes a detailed family history assessment and specific information about FH, including management guidelines, information about local specialists who treat patients with FH, recommendations for family members, and facilitation of genetic cascade testing of family members.
- A follow-up mailing or encrypted email to participants, corresponding to the variant confirmation result, reiterates the clinical implications (see MVP-ROAR-FH Patient Results Letter Positive or Negative).This mailing also includes a clinical variant report (see Clinical Variant Report), a family letter (see MVP-ROAR-FH Family Letter) that the participant may choose to share with family members, and available FH patient resources, as applicable.
- The GC also sends the participants' PCPs a clinical variant report along with physician-level information about FH treatment guidelines and a list of specialists in the area who treat patients with FH (see MVP-ROAR-FH Physician Letter (Immediate Results) Positive or Negative), as applicable. This letter provides the contact information for the GC, who is available to answer additional questions from providers.
- The clinical variant report is submitted to the participant's local VA for scanning into the medical record (See Clinical Variant Report). The genetic counselor also enters a clinical note in the local medical record summarizing the result (See Sample Post-counseling CPRS Note).
    - In the event the reference laboratory changes the classification of the result (e.g. likely pathogenic to pathogenic variant) the GC will send the participant a letter explaining the change (MVP-ROAR-FH Reclassification Letter). The updated result will also be shared with the participant' PCP and entered into the medical record.
- The essence of the study intervention is a genetic test result and supporting information about its clinical significance. Participants and providers may act on this information as they see fit. The study intervention does not include a specific treatment regimen or other protocolized management strategy.
- If the participant's VA PCP contacts the GC for additional guidance, the GC can offer to facilitate a consultation between the PCP and a cardiologist associated with the study, who can provide management recommendations to the PCP.

Participants randomly assigned to the Delayed Results arm receive a letter informing them of their randomization status and including their cholesterol results (see MVP-ROAR-FH Baseline



VA Central IRB
Effective Date: October 4, 2022

Letter to Delayed Results Arm). They and their PCPs receive the above intervention 6 months after enrollment, after completing the end-of-study data collection procedures.

5.1.4. Study population

The eligible study population includes any living MVP participant with a FH-associated genetic variant categorized as Pathogenic or Likely Pathogenic using ACMG-AMP variant interpretation criteria. Among the first >700,000 MVP participants genotyped, approximately 750 meet these criteria. We will enroll approximately 254 participants (10 for the pilot study and 244 for the randomized trial). Per standard clinical practice, the genetic counselor will facilitate cascade genetic testing among family members of participants found to have a clinically confirmed FH variant, but family members will not be considered research subjects, and no research data will be collected about them.

Upon completion of end of study data collection, study staff will invite patient-participants' VHA PCPs to participate in a brief interview about their experience.

5.1.4.1 Potentially vulnerable subjects

The MVP-ROAR-FH study population, including Veteran participants and PCP participants, may include some subjects considered vulnerable, including students, economically and/or educationally disadvantaged persons, or patients with debilitating or terminal illness. As a result, all potential participants will be informed during the consent process that participation in the study is entirely voluntary and that a decision to not participate or to withdraw from the study at any time has no bearing on the provision of medical care or the receipt of benefits to which the participant is otherwise entitled. Moreover, all participants will be provided opportunities to ask questions of the study staff as well as to consult others, including their families and/or providers, prior to participation. During the study, all participant primary care providers within VA will be provided information regarding their patients' engagement in the study. The study will not include potentially vulnerable participants who are pregnant, incarcerated, or are unable to adequately understand study procedures and provide verbal consent to participate in the study. If it is suspected by the study staff that a participant meets vulnerable subject criteria during recruitment or enrollment, such information will be reported to the study PI for further assessment, action (*i.e.* study withdrawal), and/or required reporting.

5.1.5. Risks to participants

5.1.5.1. Anticipated risks

A principal risk of genetic testing is a breach of confidentiality, in which sensitive information concerning a patient's genetic risk for disease becomes known. Moreover, the return of a genetic test result may lead to the diagnosis of a genetic condition and placement of such information into the medical record. As a result, there may be a risk that genetic information collected from participants during this study is used for the purposes of discrimination with regard to their health insurance or their job. Such recognition of genetic disease risk and/or diagnosis may affect their future insurance costs and/or coverage, such as denial of health, life, disability, or long-term care coverage. There are state, federal, and VA protections that prevent health insurance companies, group health plans, and most employers from discriminating against participants based on their genetic information.



VA Central IRB
Effective Date: October 4, 2022

It is possible that participants may be distressed by learning the results of their genetic test. It is also possible that participants may feel anxious or distressed about being randomized to the 6-month Delayed Results arm. Patient assessments including surveys and blood draws may also involve some risk. Some patients may experience distress or discomfort when answering questions about personal demographic and/or medical issues. Common risks associated with venipuncture include minor discomfort, lightheadedness, infection, or bruising at the site of the blood draw.

VHA primary care providers will be contacted to participate in semi-structured interviews to explore the facilitators and barriers to return of genetic results within MVP. Interviews pose minimal risk to VA PCPs. They may experience embarrassment during the interview if they feel they do not know the “correct” answers to questions about genetic testing. They may also perceive coercion to participate in the study as a VA employee. There is also the unlikely risk to privacy of a breach of study data.

5.1.5.2. Minimization of risks

Prior to participation, all participants will undergo a detailed informed consent process by a genetic counselor (see section 5.3 Informed Consent Procedures). The genetic counselor will explain the risks and benefits of genetic confirmation testing and the risks and benefits of participation in this randomized control trial. The informed consent document will include all required elements of consent for study participation, including an explanation of the purposes of genetic testing, a description of known risks associated with genetic testing, a description of any benefits to the patient or others (e.g., familial cascade testing) that may reasonably be expected from genetic testing, and a statement describing the extent, if any, to which confidentiality of patient medical records, data, and/or samples identifying the patient or their genetic test results will be maintained. At any time, patients may elect not to participate.

High levels of distress during patient survey assessments and blood draws are uncommon and staff will be trained to navigate such occurrences. Clinical blood samples will be collected by trained phlebotomists at patients’ local VA sites.

Genetic testing results and related clinical information will be returned to participants and their PCPs, for the purposes of medical follow-up, by a trained GC. This information will be documented in the VA medical record, accessible to VA clinicians and others providing routine and/or specialty (*e.g.* cardiology, lipidology, genetics) care.
Participants and their healthcare providers will have access to a dedicated GC throughout the entirety of the study, who will provide pre- and post-genetic test counseling and as-needed consultation. The GC will serve as a resource for both participants and their providers, and, in addition to genetic counseling, may provide education, make appropriate clinical referrals, and be available for additional support, questions, and concerns as necessary. Participants who are noted to be anxious, either by their genetic test result or randomization to the 6-month Delayed Results arm, will be identified by the GC and cases will be discussed with the principal investigator. The GC and principal investigator will use their best clinical judgment to determine the safest path forward for the participant which could include 1) engaging the participant ‘s provider 2) referral for additional clinical care (e.g., mental health or specialty care), and/or 3) withdrawal from the study (see section 5.7 Withdrawal of Subjects).

There may be other risks that are currently unknown. We will inform participants if any new information is discovered about the risks of taking part in this study and take steps to mitigate them as needed. Any adverse event or reaction experienced by participants in this study,



VA Central IRB
Effective Date: October 4, 2022

including those associated with genetic testing and/or survey or clinical assessment, will be reported by study staff to the VA Central IRB (see Section 6.0 Reporting).

For the PCP interview study, VA PCPs may experience embarrassment during the interview if they feel they do not know the “correct” answers to questions about genetic testing. This will be minimized by the use of skilled interviewers. They may also perceive coercion to participate in the study as a VA employee. This will be minimized by a clear statement in the recruitment email and at the beginning of the interview that participation or non-participation will in no way impact their VA employment or performance evaluation. There is also the unlikely risk to privacy of a breach of study data. This will be minimized through use of VA-approved methods, including: collection of data via Microsoft Teams, storage of data in VINCI behind the VA firewall accessible only to approved study staff, and removal of all personal identifiers before the presentation or publication of data. The study is performed under a Certificate of Confidentiality.

5.1.5.3. Benefits

Potential benefits to participants include the acquisition of clinical information important to their current and future medical care. Specifically, awareness of FH risk by participants and their providers may aid in the management of a yet unknown, suspected, or current FH diagnosis or for identifying risk of future disease. This information would allow participants to collaborate with their providers to develop improved treatment, surveillance, and/or prevention options related to their FH risk. Early studies also show benefits of family cascade screening as a mechanism to identify at risk family members after known cases of FH are discovered[34].

Benefits to society include an improved understanding of FH genetic testing and return of FH genetic test results. Very limited data exist regarding the benefits or harms of disclosing FH mutations to individuals and their families, especially individuals identified through population-level screening, such as MVP, rather than those who are selected for genetic testing due to their own personal or family history of high cholesterol or FH. Population screening for FH genetic variants is not currently the standard of care. This study will allow us to obtain outcomes to explore how participant and provider knowledge of FH impacts medical care compared with currently accepted standards of care. This information may inform future policy, screening, and management guidance for FH, and for the use of genetic testing and the return of results more generally.

VA PCPs participants may learn important information about the genetic testing process, but they will otherwise gain no direct benefit from participation. As a healthcare organization, VHA will benefit from applying the insights learned from this research to its care of the Veteran population.

5.1.5.4. Comparison of risks and anticipated benefits to patients and society

The risks associated with genetic testing and the return of FH genetic test results in this study are minimal and not dissimilar to what may occur in routine medical care. The potential benefits for confirming and returning FH genetic test results may lead to enhanced awareness of FH and improved treatment for patients with known FH risk. Without rigorously assessing the value of returning FH genetic test results compared to currently accepted standards, the true benefits are unknown. The risk/benefit ratio for the conduct of this study is favorable to the proposed intervention.

### 5.2. Recruitment Methods



VA Central IRB
Effective Date: October 4, 2022

5.2.1. Identification and recruitment of subjects

MVP Core study staff queries MVP databases for living participants with an eligible FH variant. The MVP Core study team mails eligible participants a letter introducing this new MVP-related study and giving participants the opportunity to opt out of further contact about the study by returning a prepaid opt out postcard or by calling the MVP Call Center. To any participant who does not opt out within 2 weeks of this initial mailing, the MVP-ROAR study team mails a letter giving more detail about the study, including all necessary informed consent information (see MVP-ROAR-FH Informed Consent Letter and MVP-ROAR-FH Informational Sheet). Participants in the non-randomized pilot of study procedures will receive a letter introducing the study and an informational document including all elements of informed consent (see MVP-ROAR-FH Informed Consent Letter and MVP-ROAR-FH Informational Sheet (Pilot)).Two weeks after this mailing, the study genetic counselor calls the participant to review the informed consent information, answers any questions about the study, and documents verbal consent or decline (see MVP-ROAR-FH GC ICF Phone Script).

Upon completion of the return of results, PCPs who provide care for a participant enrolled in the trial (n~ 250) will be recruited for interviews. Study staff will first contact the PCPs via email, sent to their VA email address, introducing the interview study and providing the option to opt out of future contact (see MVP-ROAR-FH PCP Email Invitation). A maximum of three emails will be sent. If no response is received, a maximum of three additional total attempts by phone or Microsoft Teams will be made to contact the PCP. Study staff will schedule an interview date and time with interested PCPs. Before the interview, staff will review the informed consent information, answer any questions about the interview study, and document verbal consent or decline. A total of 10-20 PCPs will be interviewed.

5.2.2. Participant incentives

Participants will be mailed a check for $50 after completing the end-of-study survey and biospecimen collection.

VA PCPs will not be provided an incentive to participate in the interview study.

#### 5.3 Informed Consent Procedures

5.3.1 Remote consent

We are requesting a waiver of HIPAA authorization and waiver of documentation of informed consent to enable remote consent by telephone or videoconferencing for eligible MVP participants and their VA PCPs nationwide.

5.3.2 Procedure

The study staff will mail eligible participants a letter including all elements necessary for informed consent (see MVP-ROAR-FH Informed Consent Letter, MVP-ROAR-FH Informational Sheet, MVP-ROAR Informational Sheet (Pilot)). The informed consent letter will describe the study, the study procedure, the risks and benefits of participation in the study, confidentiality, data security, and collection and use of health data. Upon receipt of this letter, participants may contact the study staff to ask questions about the study and to arrange a time for the study GC



VA Central IRB
Effective Date: October 4, 2022

to follow-up for the formal review of informed consent information. If a participant does not contact the study staff after a period of two weeks, the study GC will call the participant to confirm receipt of the informed consent letter and ask whether the participant would like to consent to study participation. Participants uncertain of study participation during this call will be permitted as much time as needed to review the document and to consider enrollment. If the patient is interested in study participation and demonstrates an understanding of the nature of the study and consent process, the GC will formally review the informed consent information with the participant by phone (see MVP-ROAR-FH GC IC Phone Script). The GC will log participant consent in a data file, which will include the date letters are mailed, the dates of participant phone contact, and the date phone consent is obtained. Upon verification of consent, the study staff will conduct the baseline study survey and arrange for the collection of biospecimen during this call or on another call scheduled at a convenient time for the participant. If a participant cannot be reached after at least three attempts to contact them by phone, they will be considered lost to follow-up. These individuals will be sent a letter, including a brief description of the study and study team contact information in the event they become interested or have questions (see MVP-ROAR-FH Loss to Follow-Up Letter or MVP-ROAR-FH Loss to Follow-Up Letter_General). This letter will be sent via certified mail.

For the PCP interview study, study staff will document informed consent by asking the PCP to state their full name and date of the interview and that they agree to participate in the study and have the interview recorded.

<u>5.3.3 Human subjects protection training</u>

The PI and research staff will maintain up-to-date required human subjects training certificates, including Good Clinical Practice, Privacy and HIPAA Focused Training, VA Privacy and Information Security Awareness and Rules of Behavior, and Research Compliance.

#### 5.4 Inclusion/Exclusion Criteria

A participant is eligible for enrollment in this study if he/she meets the following criteria:

- Is a living enrollee in MVP
- Is identified to have a Pathogenic or Likely Pathogenic variant in an FH-associated gene in their MVP genotype data (see Section 2.3)
- Has not previously undergone genetic testing for familial hypercholesterolemia. Study staff first ascertain this by review of the medical record and then confirm during the informed consent call by asking the participants about any prior genetic testing he/she has undergone (see MVP-ROAR-FH GC IC Phone Script).
- Is not incarcerated
- Is not pregnant

A PCP participant is eligible for enrollment in the interview study if they meet the following criteria:

- Has received genetic testing results for a Veteran enrolled in MVP-ROAR



VA Central IRB
Effective Date: October 4, 2022

- Is actively practicing at a VHA location

#### 5.5 Study Evaluations

Study data will come from the following sources and procedures:

5.5.1. MVP database

The MVP Core study staff will provide the MVP Return study staff a data file with the following information about potentially eligible participants:

- Name, date of birth, and last 4 digits of Social Security Number (SSN)
- Genotype in the *APOB*, *LDLR*, *LDLRAP1*, *PCSK9*, and *SLCO1B1* genes from their MVP genetic data
- Race and ethnicity
- VA station where participant enrolled in MVP and VA station(s) where participant currently receives health care

5.5.2. Medical record review

Study staff will review each potentially eligible participant's medical records to confirm the absence of a prior positive genetic test result for FH and to abstract the participant's list of medications.

5.5.3. Informed consent call

At the beginning of the informed consent call (see MVP-ROAR-FH GC IC Phone Script), the GC confirms the participant's name, date of birth, and last 4 SSN digits. He/she then also asks whether the participant has had prior genetic testing and, if so, what kind. Participants whose responses indicate, in the clinical judgement of the GC, a prior positive genetic test result for FH are ineligible. During this call the GC will collect or confirm participant contact information including preferred phone number, mailing address, and an email address.

5.5.4. Baseline telephone survey

The baseline survey (see MVP-ROAR-FH Baseline Survey) may occur on the same phone call as the informed consent process or on a separate call. Participants may complete the baseline survey in paper form by request (see MVP-ROAR-FH Survey Cover Letter). The survey collects the following information from the consented participant:

- Name and station of participant's VA PCP
- Name and contact information of any PCP outside of VA, if applicable



VA Central IRB
Effective Date: October 4, 2022

- Confirmation of baseline VA medication list, as assessed from medical record review
- Medication name and dose for any over-the-counter medications or medications prescribed by a provider outside VA
- Beliefs about medications: Beliefs About Medicines Questionnaire (BMQ)[44]
- Patient activation: Patient Activation Measure (PAM)[45]
- Quality of life: Veterans RAND 12-item Health Survey (VR-12)[46]
- Race and ethnicity

5.5.5. Baseline biospecimen collection

Participants present to their local laboratory for a fasting blood draw for two tests:

- Baseline lipid panel testing: This is performed by a clinical laboratory at a VA facility.
- FH variant confirmation: For participants randomized to the Immediate Results arm, this is performed by an external Clinical Laboratory Improvement Amendments (CLIA)-certified, College of American Pathologists (CAP)-accredited laboratory. This laboratory uses standard clinical techniques to sequence FH-associated genes and confirm the presence or absence of the suspected FH variant from the MVP genetic data. The laboratory sends a typical clinical report back to the study staff (see Clinical Variant Report). For participants in the Delayed Results arm, the second baseline blood specimen will be discarded.

  Participants for whom it is not feasible to visit their local VA facility for a blood draw may provide a self-collected at-home saliva sample for FH variant confirmation. To utilize this option, participants must have:
  - A documented LDL-C level in their VA health record not older than 6 months from their date of enrollment
  - Absence of a statin prescription change within the time from the most recent documented LDL value to enrollment

  For participants in the Immediate Results arm, the saliva sample will be returned to the study staff and shipped to the CLIA-certified laboratory for FH variant confirmation. For participants in the Delayed Results arm, the saliva sample will be returned to the study staff and stored in a locked and secure cabinet for six months, at which time the sample may be shipped to the laboratory for variant confirmation or discarded if the participant is able to visit their local VA facility for a blood draw.

5.5.6. End-of-study telephone survey

Six months after enrollment, the study staff calls each enrolled participant to administer the end-of-study survey (see MVP-ROAR 6-Month Survey (Immediate Results version) and MVP-ROAR-FH 6-Month Survey (Delayed Results version)). Participants may request to complete the follow-up survey by paper (see MVP-ROAR-FH Survey Cover Letter). The follow-up survey includes the following questions and instruments:



VA Central IRB
Effective Date: October 4, 2022

- Confirmation of current VA medication list, as assessed from chart review
- Medication name and dose for any current over-the-counter medications or medications prescribed by a provider outside VA
- Beliefs about medications: Beliefs About Medicines Questionnaire (BMQ)
- Health behaviors: Smoking status[47], saturated fat intake[48], physical activity[49]
- Quality of life: Repeated measurement of VR-12[46]
- Healthcare utilization, including laboratory tests, office visits to PCP and specialists, and hospitalization[50]
- Veteran time demands and transportation costs to attend CHD-related medical appointments[51]
- Whether and how many first-degree family members underwent cascade genetic testing
- Feelings about genomic testing results: End-of-study measurement of FACToR[52]
- Preferences for receiving genetic test results

<u>5.5.7. End-of-study biospecimen collection</u>

After the end-of-study survey, participants present to their local laboratory for an end-of-study fasting blood draw:

- End-of-study lipid panel testing: This is performed by VA clinical laboratory.
- FH variant confirmation: For participants randomized to the Delayed Results arm, a second blood specimen is drawn or self-collected saliva sample obtained for FH variant confirmation at an external reference laboratory, as described in Section 5.5.5 above.

<u>5.5.8. End-of-study VA database review</u>

Study staff will review each enrolled participants' medical record, Corporate Data Warehouse data, and other VA databases for the following clinical processes and outcomes related to FH:

- Lipid panel results prior to enrollment and during the study period
- Other clinical tests or procedures related to CVD, such as coronary computed tomography scan, C-reactive protein, lipoprotein(a), electrocardiogram (ECG), transthoracic echocardiogram, stress test, angiography, coronary artery bypass surgery
- All prescriptions of lipid-lowering medications prior to enrollment and during the study period
- Diagnoses related to hypercholesterolemia, FH, CHD, ischemic stroke, peripheral artery disease
- Healthcare utilization and cost data from the Health Economics Resource Center (HERC) and Managerial Cost Accounting System (MCA) datasets.



**VA Central IRB**
**Effective Date:** October 4, 2022

5.5.9. Intervention cost accounting

The costs of the intervention, including materials and personnel time, will be collected to enable the budget impact analysis described in Section 5.6.2.

5.5.10 PCP interviews

After participants receive their genetic testing results and complete the end of study survey, study staff will contact the participants' VA PCPs to complete interviews. We will invite PCPs who have received a genetic testing result from MVP-ROAR to participate. We will conduct semi-structured interviews (see interview guide) to examine the facilitators and barriers to the return of genetic testing results to Veterans in MVP. The interviews will begin with a brief introduction to the MVP-ROAR Study and explore the PCP's thoughts about genetic testing in general and experience with familial hypercholesterolemia. The interviewer will then explore potential benefits and risks of returning genetic testing results to MVP participants and the successes and limitations of the MVP-ROAR Study.

### 5.6 Data Analysis

5.6.1. Sample size determination

We will enroll a total of 254 patient-participants: 10 for the non-randomized pilot trial and 244 for the RCT.

Sample size is based on the primary outcome of change in LDL-C in each arm after 6 months. Assuming a mean LDL-C reduction of 20% in the Immediate Results arm, a mean LDL-C reduction of 0% in the Delayed Results arm, and a common standard deviation of 30%[53], 72 total participants (36 per arm) are needed to have 80% power to detect a significant between-group difference at alpha=0.05. Enrollment of twice this number (144 total) will account for an absence of therapy escalation in up to 50% of participants in the Immediate Results arm. Enrollment of 180 total participants will account for up to 20% loss to follow-up.

| **Change in LDL-C at 6 months** | | | **Total sample size required** |
|---|---|---|---|
| **Immediate Results** | **Delayed Results** | **Common SD** | |
| -20% | 0% | 30% | 72 |
| -20% | 0% | 40% | 126 |
| -20% | -5% | 30% | 126 |
| -20% | -5% | 40% | 224 |

An important secondary outcome is the proportion of participants in each arm meeting accepted LDL-C targets at 6 months (<100 mg/dL for primary prevention and <70 mg/dL for secondary prevention). In preparatory-to-research analyses, only 175/322 (46%) MVP participants with a potentially pathogenic *LDLR* variant had a most recent LDL-C <100mg/dL. To have 80% power to detect a between-arm difference of 20% of participants meeting this LDL-C target at



VA Central IRB
Effective Date: October 4, 2022

alpha=0.05, up to 194 total participants are needed (97 per arm). To account for up to 20% loss to follow-up, a total of 244 participants (122 per arm) are needed.

| Proportion of participants with LDL-C < 100mg/dL at 6 months | | Total sample size required |
|---|---|---|
| **Immediate Results** | **Delayed Results** | |
| 10% | 30% | 124 |
| 20% | 40% | 162 |
| 30% | 50% | 186 |
| 40% | 60% | 194 |

5.6.2. Data analysis plan

We will conduct intention-to-treat analyses to compare all outcomes in the Immediate and Delayed Results arms. Logistic regression will be used for dichotomous outcomes, and linear regression will be used for continuous outcomes. Poisson regression will be used to compare counts of first-degree family members undergoing cascade genetic testing between the two arms. Regression models will test the study hypotheses by including terms for randomization status. Covariates may be included if they improve model precision. Missing data will be imputed using fully conditional specification.

We will perform a budget impact analysis of the intervention alongside the randomized trial, using International Society for Pharmacoeconomics and Outcomes Research[54] and Second Panel on Cost-Effectiveness in Health and Medicine[51] recommendations. We will use administrative data and microcosting and gross costing strategies[55] to estimate the cost of the return-of-results intervention itself plus patient-level healthcare costs extracted from the VA Corporate Data Warehouse from the 6 months after enrollment. Mean Veteran health-related quality of life (VR-12) will be compared between the two study arms and used to inform cost-effectiveness analyses by mapping scores to the VR-6D to estimate health utilities.

For the PCP interview study, we will use team-based coding and qualitative analysis to identify and describe emerging themes. After the team generates a priori concepts of interests, two team members will code a small set of transcripts independently to identify additional emergent concepts to be included in the codebook, develop code definitions, and align their approaches in applying the codebook. Disagreements will be resolved via consensus, with other team members' input sought if necessary. Throughout the coding process, coders will use memos to capture insights on emergent themes in the data, including differences and similarities across the dataset. After coding is completed, coders will review data within and across codes to generate preliminary themes; the full team will be involved in refining and elaborating on themes.

All data will be stored on encrypted, password-protected VA servers and analyzed only by credentialed individuals identified as study staff on the IRB protocol.

#### 5.7 Withdrawal of Subjects



VA Central IRB
Effective Date: October 4, 2022

Study subjects will be made aware that they may withdraw from participation in this study at any time without penalty or loss of VA or other benefits to which they are entitled. It is expected, given the limited risk of the study intervention, the reporting of relevant information to participants' providers, and participants' receipt of usual care throughout their study participation, that subject withdrawal will be rare. If study staff develop serious concerns that a participant is highly distressed, anxious, depressed, or whose health and well-being may otherwise be immediately compromised as a result of his or her participation in the study, and at the discretion of the PI, the study team may inform the participant that he or she may withdraw from the study. Researchers may continue to use patient data collected prior to withdrawal. No further health data will be collected after a participant has withdrawn from the study.

Study participants may withdraw from the study by contacting the study team by phone and requesting withdrawal verbally. Once a verbal request for withdrawal is received by the study team, participants are provided with confirmation of their withdrawal from the study (see MVP-ROAR-FH Study Withdrawal Letter). If a participant withdraws before the baseline biospecimen collection, no information about the participant's MVP research result will be provided to the participant. If a participant withdraws after the baseline biospecimen collection but before the reference laboratory has analyzed his/her sample, the study staff will contact the lab to destroy the sample before analysis. If a participant withdraws after the laboratory analysis but before receiving his/her results from the GC, he/she will be given the opportunity to receive the results from the GC and/or have them sent to his/her PCP, 6 months after enrollment.

Primary care providers enrolled in the interview study may withdraw at any time by contacting the study team by email, phone, or Teams and requesting withdrawal.

### 6.0 Reporting

#### 6.1. Monitoring and quality assurance

The PI will monitor the study proceedings to ensure the study is executed according to the IRB-approved protocol, which includes ensuring the safety of participants and the validity, integrity, and protection of data associated with this study. The PI and research staff will be responsible for the day-to-day monitoring of patient safety and data quality throughout the conduct of this study. In addition to the regular monitoring of patient safety and data quality, the research team will complete any and all continuing reviews, audits, event reporting, and/or other requirements per the provisions and timelines set forth by VA Central IRB.

Any concerns regarding the ethical conduct of the study, the safety of participants, or a breach in the protection of study data made by participants, the study staff, or others will be promptly reported to the PI and escalated accordingly to VA Central IRB and/or other relevant oversight committees.

#### 6.2 Participant safety

Participant safety will be monitored by the PI and study staff throughout the conduct of study activities. All participants will have the contact information of the MVP Information Center and the MVP-ROAR-FH study team for any questions or concerns related to study participation. All participants will also have the contact information of the study GC should they have any concerns with their results or their delayed result disclosure. Per the study protocol the participant's PCP will also receive a copy of the participant's genetic test results. If a participant



VA Central IRB
Effective Date: October 4, 2022

experiences adverse effects or expresses emotional distress related to study participation at any time during their enrollment and requires medical attention based on the judgement of the PI, the study genetic counselor, and/or their PCP, they will be referred for clinical assessment and/or informed of options for study withdrawal (see Section 5.7 Withdrawal of Subjects) as appropriate. All serious such cases, including those requiring a referral to a mental health professional or other therapeutic intervention, will be reviewed and reported to VA Central IRB as an adverse event (AE) or serious adverse event (SAE) as necessary.

During the PCP interview study, if any PCP participant exhibits or expresses distress during any interview, the interviewer will offer a break to the participant and will offer the option of discontinuing the interview. If any PCP participant becomes overly agitated, distressed, or threatening the interviewer will discontinue the interview and discuss the matter with the PI.

Throughout the conduct of the study, attention will be paid to any reports of participant experiences that constitute adverse events as described in Section 6.3 Adverse events and will be reported per Section 6.4 Event reporting.

#### 6.3 Adverse events

Adverse events (AEs) related to MVP-ROAR-FH procedures do not include anticipated events related to blood draws (*e.g.*, pain, minor bleeding, bruising, fainting, or lightheadedness) and minor feelings of discomfort while answering survey questions. Pre-existing conditions or illnesses which are expected to exacerbate or worsen are also not considered adverse events and will be accounted for in the subject's medical history. An AE may be considered any other unanticipated or unintended medical occurrence or worsening of a sign or symptom (including an abnormal laboratory finding other than the return of genetic information associated with FH) or disease in a study subject, which does not necessarily have a causal relationship with the study condition, procedure(s) or study agent(s), that occurs after participant informed consent is obtained. A serious adverse event (SAE) will be defined as an AE resulting in one of the following outcomes: death during the 6 months after study enrollment, life threatening event (defined as an event that places a participant at immediate risk of death), inpatient hospitalization, and any other condition which, in the judgment of the PI, represents a significant hazard, such as an important medical event that does not result in one of the above outcomes. An event may be considered an SAE when it jeopardizes the participant or requires medical or surgical intervention to prevent one of the outcomes listed above. AEs may be observed by the study staff or volunteered by participants, their family members, their PCPs, or others. All AEs and SAEs will be assessed for relationship to the study research procedures by the study PI, to determine whether study participation was likely to have caused the AE/SAE.

#### 6.4 Event reporting

Given the minimal risk of the study intervention, the study team anticipates few AEs, SAEs, or unanticipated problems (UPs) during the course of this study. AEs, SAEs, and UPs may be observed through regular monitoring by the study staff or volunteered by participants, their family members, their PCPs, or others throughout the conduct of the study. Any concerns related to patient safety or the potential occurrence of an AE, SAE, or UP in relation to the conduct of this study will be promptly reported to the PI for review. Upon discovery, any study-related death will be immediately reported to the VA Central IRB. Study staff will report any protocol deviation that substantively affects subjects' rights or safety, UP that poses risk to participants, or SAE per VA Central IRB protocols. An annual report summarizing all non-serious adverse events and UP/protocol deviations that did not require immediate reporting will



VA Central IRB
Effective Date: October 4, 2022

be prepared and reported to the VA Central IRB at the time of continuing review. Acknowledgement of any AE, SAE, or UP/protocol deviation by the VA Central IRB will be retained by the MVP-ROAR-FH study staff.

### 7.0 Privacy and Confidentiality

#### 7.1 Use of Protected Health Information (PHI)

The MVP-ROAR-FH study team will utilize participant Protected Health Information (PHI) during the conduct of study activities, including in the identification and recruitment of eligible participants, for genetic confirmation testing, for the delivery of the study intervention, and for the collection of study data. Given the use of PHI, every effort will be made to ensure that the privacy and confidentiality of MVP-ROAR-FH participants are maintained. As with any research study, loss of privacy and/or confidentiality is a potential risk. To mitigate this risk, study personnel will take every precaution to keep participants' PHI confidential and protect each participant's privacy in all aspects in which participant data is used as part of this research study. This includes using coded information rather than PHI whenever possible, storing electronic study-related data on VA and/or VA-approved servers in accordance with appropriate information security policies, using only VA-approved methods for the transfer of data, and storing any participant-related written and/or paper documents in a secure environment (e.g., locked cabinet and in a locked office).

##### 7.1.1 Biospecimens and genetic variant data

Blood specimens will be collected at participants' local VA facilities and shipped to the VA Boston Healthcare System laboratory for FH variant confirmation testing at a VA-approved CLIA-certified laboratory. Saliva collection kits will be sent directly to participants using a common carrier delivery service and will be tracked with a unique reference number. Saliva samples will be self-collected in the participant's home and shipped to study staff for FH variant confirmation testing at a VA-approved CLIA-certified laboratory. All participant data will be retained within the VA except the following in order to complete genetic variant confirmation testing: patient name, date of birth, gender, and a test requisition form will be included with the blood sample sent to the VA-approved CLIA-certified laboratory for genetic confirmation testing, using a common carrier delivery service and chain of custody. Specimens will be shipped to the VA-approved clinical laboratory via prepaid standard biospecimen collection kits. The shipment to the VA-approved CLIA-certified laboratory will not include any personal identifiers on the external packaging. Each shipment will be distributed for overnight delivery and will be tracked with a unique reference number. The VA-approved CLIA-certified laboratory will perform variant confirmation and then retain the sample only as long as is required by internal quality assurance and other regulatory procedures. The VA-approved CLIA-certified laboratory will return the results of variant confirmation testing to the study staff via a secure, password protected, and VA-approved web-interface. The study staff will transfer the clinical variant confirmation test results to the medical record at participants' local VA facilities. Copies of the participants' results will also be provided to their designated VA PCPs via secure VA-approved methods.

#### 7.2 Staff training and data access

All study staff with access to PHI will complete required training and be instructed, in accordance with VA policy, on the requirements of Federal privacy and information laws and regulations, VA regulations and policies, and VHA policy. Only study personnel credentialed and



VA Central IRB
Effective Date: October 4, 2022

approved by the VA Central IRB and VA Research & Development committees will have access to study data stored in either physical or electronic environments. Once study team members are no longer a part of the research team, their access to data and research materials will be terminated. No unauthorized access to study servers or datasets will be permitted. All study staff will be trained on reporting data breaches within one (1) hour to the appropriate Information Security and Privacy Officers and VA Central IRB. Access to all electronic data and files (e.g., database, spreadsheet) containing identifiable patient information will be limited to approved users with a login credential. Any computer hosting such files will be password protected to prevent access by unauthorized users.

7.3 Data security

Risk of breach of confidentiality will be minimized through the appropriate management and security of participant data per VA, HIPAA, and MVP requirements. Participant PHI will be delinked from the final analytic dataset. All data will be retained within the VA. Data will be securely transmitted using VA approved methods, including FIPS 140-2 validated encryption. This will include transmission of PHI and other participant data, including genetic testing results, between VA and the commercial reference laboratory performing genetic confirmation testing. Participant data files (source and analytic) will be stored behind the VA firewall, on a drive created specifically to house the data for this research project. No PHI (including scrambled SSNs or dates) will be released to the public, nor will they be presented or published in the dissemination of study findings. All written and/or paper documents containing participant PHI will be filed in a locked cabinet and locked office at the VA Boston Healthcare System. Study data will be kept indefinitely or until the law allows their destruction in accordance with the VA Record Control Schedule. Electronic records will be destroyed, when allowed, in a manner in which they cannot be retrieved. In addition, a Certificate of Confidentiality, issued by the National Institutes of Health (NIH), will help to ensure the privacy of participant identities and data. With this certificate, researchers cannot be forced (e.g. by court subpoena) to disclose information that identify the participant in federal, state, or local civil, criminal, administrative, legal or other proceedings.

7.3.1 Participant mailing data

A copy of patient-participant mailing data only will be downloaded outside of VINCI and stored on a secure, study specific SharePoint site. The SharePoint site will be housed behind the VA firewall and viewing of mailing data will be limited to IRB approved study personnel. This will be done to enable the use of Microsoft Word mail merge software to create participant letters and address labels for efficient printing and distribution. Participant mailing data will be in the form of CSV files and may include identifying variables including: participant ID, full name, gender, mailing address, and any associated flags (i.e. temporary address). The mail merge software can be used within the secure SharePoint environment.

7.3.2 Use of encrypted emails

The MVP-ROAR-FH Study will include the option for participants and their providers to receive encrypted email correspondence associated with IRB-approved study materials and study-related appointments (*i.e.,* local facility blood draws, remote genetic counseling appointments). Per VA policy, any email distributed external to the VA network and including individually identifiable information (III), PHI, or study specific information will be encrypted using the VA-approved Microsoft Azure Rights Management Service (RMS).



VA Central IRB
Effective Date: October 4, 2022

Only after initiating contact by standard mail and telephone, and after participants have had the opportunity to opt out of contact, patient- participants will have the option to request that study staff send recruitment and other study materials (i.e. informational sheet, baseline survey, end-of-study survey, delayed arm letter, VA form 10-5345, results mailings) which may contain PHI or study-specific information via Azure RMS encrypted email. Participants may also opt to return study materials (i.e. baseline survey, end of study survey, and VA form 10-5345) by scanning them or taking a photo and returning it to study staff via Azure RMS encrypted email.

### 8.0 Communication Plan

8.1 Notification of local facility

As an MVP-associated study, MVP local site investigators will be made aware of the conduct of MVP-ROAR-FH and that MVP participants from their facilities may be recruited for enrollment into this study. In the event a SAE or unanticipated problem occurs in conjunction with the conduct of MVP-ROAR-FH, the MVP local site investigator of the affected participant will be informed and the event will be reported in accordance with VA Central IRB procedures.

8.2 Public data set

We will share de-identified individual-level trial data (participant characteristics and outcomes) through a data repository housed on a secure VA server and accessible only to outside investigators with IRB and other regulatory approvals. In the event of a data request, approval for data use will be sought from the local IRB where the repository is housed.

8.3 VA MVP Central Database

Identifiable, individual-level trial data will be shared with the MVP Program and will be stored in the VA MVP Central Database.

8.4 Dissemination of study results

This trial will be registered on ClinicalTrials.gov. Communication of study progress and results will be the responsibility of the PI and Executive Committee. Information associated with this project will be communicated to VA and other stakeholders including policymakers, healthcare providers, patients, and the scientific community. In order to broadly disseminate information about this research, the MVP-ROAR-FH study team 1) will prepare scientific presentations and manuscripts for publication, 2) present information regarding the purpose, methods, and results of this research at informal meetings throughout the VA, 3) inform the public and policymakers of new findings through press releases and related mechanisms, including those currently approved through MVP, and 4) facilitate dissemination of study-related information and findings to patients and healthcare providers through collaboration with MVP leadership, VA national leadership, and other centers.

### 9.0 References

1. Gaziano JM, Concato J, Brophy M, et al. Million Veteran Program: A mega-biobank to study genetic influences on health and disease. *J Clin Epidemiol.* 2016;70:214-223.



VA Central IRB
Effective Date: October 4, 2022

2. Allen NL, Karlson EW, Malspeis S, Lu B, Seidman CE, Lehmann LS. Biobank participants' preferences for disclosure of genetic research results: perspectives from the OurGenes, OurHealth, OurCommunity project. *Mayo Clin Proc.* 2014;89(6):738-746.
3. Kaufman D, Murphy J, Erby L, Hudson K, Scott J. Veterans' attitudes regarding a database for genomic research. *Genet Med.* 2009;11(5):329-337.
4. Bredenoord AL, Kroes HY, Cuppen E, Parker M, van Delden JJ. Disclosure of individual genetic data to research participants: the debate reconsidered. *Trends Genet.* 2011;27(2):41-47.
5. Bredenoord AL, Onland-Moret NC, Van Delden JJ. Feedback of individual genetic results to research participants: in favor of a qualified disclosure policy. *Hum Mutat.* 2011;32(8):861-867.
6. Levesque E, Joly Y, Simard J. Return of research results: general principles and international perspectives. *J Law Med Ethics.* 2011;39(4):583-592.
7. Wolf SM. Return of individual research results and incidental findings: facing the challenges of translational science. *Annual review of genomics and human genetics.* 2013;14:557-577.
8. Knoppers BM, Zawati MH, Senecal K. Return of genetic testing results in the era of whole-genome sequencing. *Nat Rev Genet.* 2015;16(9):553-559.
9. Bibbins-Domingo K, Grossman DC, Curry SJ, et al. Screening for Lipid Disorders in Children and Adolescents: US Preventive Services Task Force Recommendation Statement. *JAMA.* 2016;316(6):625-633.
10. Youngblom E, Pariani M, Knowles JW. Familial Hypercholesterolemia. In: Adam MP, Ardinger HH, Pagon RA, et al., eds. *GeneReviews((R)).* Seattle (WA): University of Washington, Seattle; 1993.
11. Sniderman AD, Tsimikas S, Fazio S. The severe hypercholesterolemia phenotype: clinical diagnosis, management, and emerging therapies. *J Am Coll Cardiol.* 2014;63(19):1935-1947.
12. Dron JS, Hegele RA. Genetics of Lipid and Lipoprotein Disorders and Traits. *Current genetic medicine reports.* 2016;4(3):130-141.
13. Benn M, Watts GF, Tybjærg-Hansen A, Nordestgaard BG. Mutations causative of familial hypercholesterolaemia: screening of 98 098 individuals from the Copenhagen General Population Study estimated a prevalence of 1 in 217. *Eur Heart J.* 2016;37(17):1384-1394.
14. Khera AV, Won HH, Peloso GM, et al. Diagnostic Yield and Clinical Utility of Sequencing Familial Hypercholesterolemia Genes in Patients With Severe Hypercholesterolemia. *J Am Coll Cardiol.* 2016;67(22):2578-2589.
15. Umans-Eckenhausen MA, Defesche JC, Sijbrands EJ, Scheerder RL, Kastelein JJ. Review of first 5 years of screening for familial hypercholesterolaemia in the Netherlands. *Lancet.* 2001;357(9251):165-168.
16. Jacobson TA, Maki KC, Orringer CE, et al. National Lipid Association Recommendations for Patient-Centered Management of Dyslipidemia: Part 2. *J Clin Lipidol.* 2015;9(6, Supplement):S1-S122.e121.
17. Knowles JW, Rader DJ, Khoury MJ. Cascade Screening for Familial Hypercholesterolemia and the Use of Genetic Testing. *JAMA.* 2017;318(4):381-382.
18. Nordestgaard BG, Chapman MJ, Humphries SE, et al. Familial hypercholesterolaemia is underdiagnosed and undertreated in the general population: guidance for clinicians to prevent coronary heart disease: consensus statement of the European Atherosclerosis Society. *Eur Heart J.* 2013;34(45):3478-3490a.
19. Nanchen D, Gencer B, Muller O, et al. Prognosis of Patients With Familial Hypercholesterolemia After Acute Coronary Syndromes. *Circulation.* 2016;134(10):698-709.





VA Central IRB
Effective Date: October 4, 2022

20. Besseling J, Hovingh GK, Huijgen R, Kastelein JJP, Hutten BA. Statins in Familial Hypercholesterolemia. *Consequences for Coronary Artery Disease and All-Cause Mortality.* 2016;68(3):252-260.
21. Gidding SS, Champagne MA, de Ferranti SD, et al. The Agenda for Familial Hypercholesterolemia: A Scientific Statement From the American Heart Association. *Circulation.* 2015;132(22):2167-2192.
22. Grundy SM, Stone NJ, Bailey AL, et al. 2018 AHA/ACC/AACVPR/AAPA/ABC/ACPM/ADA/AGS/APhA/ASPC/NLA/PCNA Guideline on the Management of Blood Cholesterol. *Circulation.*0(0):CIR.0000000000000625.
23. Versmissen J, Oosterveer DM, Yazdanpanah M, et al. Efficacy of statins in familial hypercholesterolaemia: a long term cohort study. *BMJ.* 2008;337:a2423.
24. Do R, Stitziel NO, Won HH, et al. Exome sequencing identifies rare LDLR and APOA5 alleles conferring risk for myocardial infarction. *Nature.* 2015;518(7537):102-106.
25. Neil HA, Hammond T, Huxley R, Matthews DR, Humphries SE. Extent of underdiagnosis of familial hypercholesterolaemia in routine practice: prospective registry study. *BMJ.* 2000;321(7254):148.
26. Benn M, Watts GF, Tybjaerg-Hansen A, Nordestgaard BG. Familial hypercholesterolemia in the danish general population: prevalence, coronary artery disease, and cholesterol-lowering medication. *J Clin Endocrinol Metab.* 2012;97(11):3956-3964.
27. Huijgen R, Vissers MN, Defesche JC, Lansberg PJ, Kastelein JJ, Hutten BA. Familial hypercholesterolemia: current treatment and advances in management. *Expert Rev Cardiovasc Ther.* 2008;6(4):567-581.
28. Mortality in treated heterozygous familial hypercholesterolaemia: implications for clinical management. Scientific Steering Committee on behalf of the Simon Broome Register Group. *Atherosclerosis.* 1999;142(1):105-112.
29. Abul-Husn NS, Manickam K, Jones LK, et al. Genetic identification of familial hypercholesterolemia within a single U.S. health care system. *Science.* 2016;354(6319).
30. Rodriguez F, Knowles JW, Maron DJ, Virani SS, Heidenreich PA. Frequency of Statin Use in Patients With Low-Density Lipoprotein Cholesterol >/=190 mg/dl from the Veterans Affairs Health System. *Am J Cardiol.* 2018;122(5):756-761.
31. Wonderling D, Umans-Eckenhausen MA, Marks D, Defesche JC, Kastelein JJ, Thorogood M. Cost-effectiveness analysis of the genetic screening program for familial hypercholesterolemia in The Netherlands. *Semin Vasc Med.* 2004;4(1):97-104.
32. Ademi Z, Watts GF, Pang J, et al. Cascade screening based on genetic testing is cost-effective: evidence for the implementation of models of care for familial hypercholesterolemia. *J Clin Lipidol.* 2014;8(4):390-400.
33. Alver M, Palover M, Saar A, et al. Recall by genotype and cascade screening for familial hypercholesterolemia in a population-based biobank from Estonia. *Genet Med.* 2018.
34. Sturm AC, Knowles JW, Gidding SS, et al. Clinical Genetic Testing for Familial Hypercholesterolemia: JACC Scientific Expert Panel. *J Am Coll Cardiol.* 2018;72(6):662-680.
35. Kalia SS, Adelman K, Bale SJ, et al. Recommendations for reporting of secondary findings in clinical exome and genome sequencing, 2016 update (ACMG SF v2.0): a policy statement of the American College of Medical Genetics and Genomics. *Genet Med.* 2017;19(2):249-255.
36. Richards S, Aziz N, Bale S, et al. Standards and guidelines for the interpretation of sequence variants: a joint consensus recommendation of the American College of Medical Genetics and Genomics and the Association for Molecular Pathology. *Genet Med.* 2015.



VA Central IRB
Effective Date: October 4, 2022

37. Green RC, Berg JS, Grody WW, et al. ACMG recommendations for reporting of incidental findings in clinical exome and genome sequencing. *Genet Med.* 2013;15(7):565-574.
38. Landrum MJ, Lee JM, Benson M, et al. ClinVar: public archive of interpretations of clinically relevant variants. *Nucleic Acids Res.* 2016;44(D1):D862-868.
39. Amendola LM, Jarvik GP, Leo MC, et al. Performance of ACMG-AMP Variant-Interpretation Guidelines among Nine Laboratories in the Clinical Sequencing Exploratory Research Consortium. *Am J Hum Genet.* 2016;98(6):1067-1076.
40. Rivera-Munoz EA, Milko LV, Harrison SM, et al. ClinGen Variant Curation Expert Panel experiences and standardized processes for disease and gene-level specification of the ACMG/AMP guidelines for sequence variant interpretation. *Hum Mutat.* 2018;39(11):1614-1622.
41. Clinical Domain Working Group: Cardiovascular Familial Hypercholesterolemia Variant Curation Expert Panel. https://www.clinicalgenome.org/working-groups/clinical-domain/cardiovascular-clinical-domain-working-group/familial-hypercholesterolemia-variant-curation-expert-panel/. Accessed December 2, 2018.
42. Iacocca MA, Chora JR, Carrie A, et al. ClinVar database of global familial hypercholesterolemia-associated DNA variants. *Hum Mutat.* 2018;39(11):1631-1640.
43. Virani SS, Woodard LD, Wang D, et al. Correlates of repeat lipid testing in patients with coronary heart disease. *JAMA Intern Med.* 2013;173(15):1439-1444.
44. Horne R, Weinman J, Hankins M. The Beliefs about Medicines Questionnaire: the development and evaluation of a new method for assessing the cognitive representation of medication. *Psychol Health.* 1999;14(1):1-24.
45. Fowles JB, Terry P, Xi M, Hibbard J, Bloom CT, Harvey L. Measuring self-management of patients’ and employees’ health: Further validation of the Patient Activation Measure (PAM) based on its relation to employee characteristics. *Patient Educ Couns.* 2009;77(1):116-122.
46. Selim AJ, Rogers W, Fleishman JA, et al. Updated U.S. population standard for the Veterans RAND 12-item Health Survey (VR-12). *Qual Life Res.* 2009;18(1):43-52.
47. Behavioral Risk Factor Surveillance System. Centers for Disease Control and Prevention. http://www.cdc.gov/brfss/annual_data/pdf-ques/2009brfss.pdf. 2009.
48. Riordan F, McGann R, Kingston C, et al. A systematic review of methods to assess intake of saturated fat (SF) among healthy European adults and children: a DEDIPAC (Determinants of Diet and Physical Activity) study. *BMC Nutrition.* 2018;4(1):21.
49. Topolski TD, LoGerfo J, Patrick DL, Williams B, Walwick J, Patrick MB. The Rapid Assessment of Physical Activity (RAPA) among older adults. *Prev Chronic Dis.* 2006;3(4):A118.
50. Vassy JL, Christensen KD, Schonman EF, et al. The impact of whole-genome sequencing on the primary care and outcomes of healthy adult patients: A pilot randomized trial. *Ann Intern Med.* 2017;167(3):159-169.
51. Sanders GD, Neumann PJ, Basu A, et al. Recommendations for Conduct, Methodological Practices, and Reporting of Cost-effectiveness Analyses: Second Panel on Cost-Effectiveness in Health and Medicine. *JAMA.* 2016;316(10):1093-1103.
52. Li M, Bennette CS, Amendola LM, et al. The Feelings About genomiC Testing Results (FACToR) Questionnaire: Development and Preliminary Validation. *Journal of genetic counseling.* 2019;28(2):477-490.
53. Lewis SJ, Olufade T, Anzalone DA, Malangone-Monaco E, Evans KA, Johnston S. LDL cholesterol levels after switch from atorvastatin to rosuvastatin. *Curr Med Res Opin.* 2018;34(10):1717-1723.



**VA Central IRB**
**Effective Date:** October 4, 2022

54. Ramsey SD, Willke RJ, Glick H, et al. Cost-effectiveness analysis alongside clinical trials II-An ISPOR Good Research Practices Task Force report. *Value Health.* 2015;18(2):161-172.
55. Barnett PG, Hong JS, Carey E, Grunwald GK, Joynt Maddox K, Maddox TM. Comparison of Accessibility, Cost, and Quality of Elective Coronary Revascularization Between Veterans Affairs and Community Care Hospitals. *JAMA cardiology.* 2018;3(2):133-141.



VA Central IRB
Effective Date: October 4, 2022
