## Supplemental Figure 1 for "Introducing return of results in the Million Veteran Program: Design and pilot results of the MVP-ROAR Familial Hypercholesterolemia Study"

VistA CPRS in use by: Danowski,Morgan (vista.boston.med.va.gov)

File Edit View Action Options Tools Help

**MOUSEA,MICKEY (OUTPATIENT)** 000-00-1300 Apr 11,1962 (61) | **BAC Aug 31,23 10:13** Current Provider Not Selected | No PACT assigned at any VA location

**COVID-19** **Not Tested**

Last 6 Signed Notes (Total: 6)

- All signed notes
  - Aug 31,23 RESEARC
  - Mar 07,23 CCC: SCHE
  - Feb 05,21 TELEPHOI
  - Dec 08,20 MH Psych
  - Aug 07,20 MH CIWA
  - Nov 18,19 MH Psych

Templates

Encounter

New Note

Visit: 08/31/23 RESEARCH/TELEPHONE, JP ADMINISTRATIVE CLINIC, morgan danowski (Aug 31,23@10:13)

GENETIC TEST RESULT NOTE
Telephone encounter with genetic counselor, Morgan Danowski, MS, CGC

RESULTS: Genetic analysis (next generation sequencing and deletion/duplication analysis) for mutations in four genes associated with familial hypercholesterolemia (LDLR, APOB, LDLRAP1, PSCK9) was POSITIVE for a heterozygous pathogenic variant in the LDLR gene (c.1784G>A). This result explains the Veteran's history of early onset hypercholesterolemia with elevated direct LDL levels.

The Veteran was contacted with this information on 8/01/2023 and had the opportunity to ask questions.

INTERPRETATION/DISCUSSION: Familial hypercholesterolemia (FH) is a common hereditary condition with an estimated prevalence of about 1/250. Mutations in the LDLR gene are associated with autosomal dominant FH. Penetrance of the condition is estimated to be >90%. Those with an FH mutation are at an increased risk of developing high cholesterol, particularly very elevated LDL-C levels. This can lead to plaque deposition increasing the risk for cardiovascular disease -especially coronary artery disease (CAD)- and death. Untreated women are at a 30% risk of having a coronary event by age 60. Men are at a 50% risk of having a coronary event by age 50. Some individuals have secondary physical findings such as xanthomas and corneal arcus.

60-80% of FH cases are attributed to mutations in the LDLR gene. Most cases of FH are inherited; it is likely one of the Veteran's parents carried the same mutation, however this was not determined by the current test. If the variant was inherited from a parent, then siblings have a 50% chance of also having the familial variant. There is a 50% chance children also have the familial LDLR variant.

MEDICAL MANAGEMENT:
On 3/23/2023 his LDL level was 150 mg/dL.

Literature and professional organization guidelines recommend the following in general for individuals with FH. The personal health care team should modify to meet this Veteran's specific needs.
- Lifestyle modification should focus on regular physical activity and weight control. A healthy diet with reduced saturated fat intake and increased (10-20 g/day) intake of soluble fiber is recommended. Limit alcohol consumption. Affected individuals should NOT use tobacco products.
- If not currently prescribed, consider initiation of statins to reduce LDL-C levels if >100mg/dL.
- If at a high risk for CAD or stroke, use of low dose aspirin (75-81 mg/day) can be considered.
- Blood pressure should be monitored and treated if >140/90mm Hg.
- Diabetes should be screened for and managed appropriately, if diagnosed.
- If statins are not working, consideration of other drug therapies and/or LDL apheresis should be considered. These may include cholesterol absorption inhibitors, Mipomersen (to decrease APOB production), MTP inhibitors, PCSK9 inhibitors, bile acid sequestrants and stanol esters (to decrease cholesterol absorption and increase LDLR activity).

PLAN:
- Local providers will continue to monitor the Veteran's LDL-C levels and monitor for any cardiac complications.
- Primary care: please discuss continuing recommended lifestyle modifications with the Veteran.
- Primary care: consider referral to a lipid specialist if LDL-C concentrations cannot be reduced after treatment with statins and lifestyle modifications.
- No further germline genetic testing is recommended for the Veteran at this time.
- The Veteran will be mailed a copy of the results and a letter summarizing them.
- Mr. Mouse was encouraged to share this information with family members as first-degree relatives are all genetic testing candidates. Testing of second-degree relatives may also be recommended. Invitae offers free testing to close relatives of those with an identified pathogenic mutation if tested prior to 09/07/2023. Blood relatives are encouraged to have regular cholesterol level screening and to discuss this result with their primary care physicians.

Duration of Call: 15 minutes

/es/ morgan danowski
HEALTH SCIENCE SPECIALIST
Signed: 08/31/2023 10:36

Cover Sheet | Problems | Meds | Orders | Notes | Consults | Surgery | D/C Summ | Labs | Reports
